# Donanemab outperformed Aducanumab and Lecanemab on cognitive, but not on biomarker and safety outcomes: systematic review, frequentist and Bayesian network meta-analyses

**DOI:** 10.1101/2024.03.31.24305134

**Authors:** Danko Jeremic, Juan D. Navarro-López, Lydia Jiménez-Díaz

## Abstract

**INTRODUCTION:** Questions remain regarding safety and clinical relevance of anti-amyloid antibodies in Alzheimer’s disease (AD), with no scientific basis for choosing between different therapies.

**METHODS:** Systematic review, frequentist and Bayesian network meta-analyses of phase III randomized placebo-controlled trials were performed to comparatively evaluate cognitive, functional and biomarker efficacy and safety of anti-amyloid antibodies in sporadic AD. Treatments were ranked with P- and SUCRA scores, with rank robustness measured by Cohen’s *kappa*, and uncertainty in ranking probabilities estimated with Shannon’s normalized entropy.

**RESULTS:** Based on data from 16,971 patients (16 studies), we found Donanemab the best-ranked antibody on cognitive measures. Lecanemab was the most effective at reducing amyloid burden. Caution is needed concerning brain edema and microbleeding, with clinically important risks for Donanemab, Aducanumab and Lecanemab.

**DISCUSSION:** Risk/benefit profile of anti-amyloid antibodies remains unfavorable. Patients in Donanemab study were stratified by *tau* load, with greater effects observed in low/medium *tau* population.

**Highlights:**

- No single therapy ranked the best among all outcomes.
- Donanemab was the most effective antibody at reducing cognitive decline across all primary outcomes, while Lecanemab ranked the highest on amyloid PET removal.
- Consistently greater cognitive, functional and biomarker effects of Donanemab were observed in patients with low/medium *tau* load, suggesting more promising effects in earlier AD stages.
- All antibodies, except Solanezumab, were significantly less tolerable than Placebo.
- The risk of cerebral edema and microbleeding may outweigh the benefits, independently of APOE status.

## 1. BACKGROUND

Although known for a little more than a century, Alzheimer’s disease (AD) became a real-world problem in the last few decades as the most common form of dementia and significant burden for our societies [1]. Until recently, symptomatic treatments were the only therapies available for AD patients, and that situation remains in the majority of countries in the world. The recent approvals of Aducanumab (Aduhelm^®^, U.S) and Lecanemab (Leqembi^®^, U.S and Japan) have initiated the new era in AD therapeutics, as these monoclonal antibodies have shown some potential to modify disease progression by binding to amyloid-β (Aβ) peptide, toxic peptides thought to be essential in the AD pathophysiology [2]. Both Aducanumab (epitope: 3–7) and Lecanemab (epitope: 1–16) have high selectivity for N-terminus of full-length Aβ found in soluble oligomers and fibrils, however, Lecanemab showed tenfold stronger binding to small and large protofibrils than to fibrils [3, 4].

The regulatory approvals of Aduhelm^®^ and Leqembi^®^ were both promising and controversial, and they came after more than a decade of investigating anti-Aβ vaccines and antibodies in AD patients [5, 6]. Many other antibodies, such as Solanezumab (specific to Aβ monomers) and Bapineuzumab (non-specific) were discontinued since they failed to show beneficial effects in clinical trials. Another anti-Aβ antibody, Donanemab, have gained significant attention recently and it is currently under review for standard approval in the EU and the U.S. Instead of binding the N-terminus of full-length Aβ, Donanemab targets the N-truncated pyroglutamate Aβ (at position 3), present only in Aβ plaques. It is reported to rapidly induce plaque removal and reduce cognitive decline in patients with early symptomatic AD [7, 8].

Despite clear advantages of disease-modifying therapies, substantial doubt remains about both the clinical relevance and safety of anti-amyloid antibodies, and they still have to demonstrate clinically meaningful effect in real life [9, 10]. We have previously conducted the systematic review and web application meta-analysis of anti-Aβ antibodies in phase III randomized placebo-controlled clinical trials (RCTs), showing that Aducanumab and Lecanemab produced the most promising biomarker results (reducing both Aβ and *tau* load) and improved cognitive outcomes by small effect size when compared to control group. However, these cognitive effects were achieved at the great expense of increasing adverse events, most importantly amyloid-related imaging abnormalities (ARIA) in the form of cerebral edema (ARIA-E), hemorrhages and localized superficial siderosis (ARIA-H) [11]. Our study had some limitations, as we performed only the conventional (frequentist) meta-analysis method with DerSimonian and Laird random-effect model. This did not allow us to estimate the relative effects and compare the performance of these treatments. Also, we did not analyze amyloid PET data from original studies, and we failed to include the effects of Donanemab, as phase III trial results on this drug were not available at the time. Therefore, our aim in this study was to update and expand our previous study by conducting random-effect frequentist and Bayesian network meta-analyses (NMAs) of phase III RCTs in order to compare the effects of anti-Aβ antibodies and rank their performance across cognitive, functional, biomarker and safety outcomes.

## 2. METHODS

### 2.1. Search strategy

We followed the Preferred Reporting Items for Systematic Reviews and Meta-analyses (PRISMA) – NMA guideline of health care interventions [12]. The comprehensive search for published phase III trials was done on March 11, 2024, using the following keywords: “Alzheimer’s”, “sporadic”, “mild cognitive impairment”, “phase 3”, “monoclonal antibody”, “passive immunotherapy”, “Aducanumab”, “BIIB037”, “Lecanemab”, “BAN-2401”, “Solanezumab”, “LY2062430”, “Bapineuzumab” and “AAB-001”, “Donanemab” and “LY3002813”. The search was performed on Google Scholar, PubMed, and clinical trial databases, including International Clinical Trials Registry Platform (ICTRP) of the World Health Organization, the U.S. Clinical trial Registry ClinicalTrials.gov, The EU Clinical Trial Registry and Australian New Zealand Clinical Trials Registry (ANZCTR).

### 2.2. Quality assessment

Modified Jadad scale was used to assess the methodological quality of clinical trial reports. The Jadad scale, also known as the Oxford quality scoring system has been tested for reliability in different settings [13, 14] and includes the evaluation of study randomization, blindness of patients and investigators, blindness in outcome assessment, report of withdrawals, and dropouts [15]. In addition, we evaluated reporting of the methodology used to assess adverse events (most notably ARIA), whether ARIA is reported depending on the APOE genotype, and whether the methods of the statistical analysis were described in the study. The studies were excluded if they (1) had modified Jadad score < 3; (2) were not phase III RCTs; (3) did not include patients with mild cognitive impairment (MCI) or sporadic AD; (4) tested less than 50 participants for cognitive and safety measures.

### 2.3. Risk of bias

The potential bias was classified as “low risk”, “with some concerns”, or “high risk, by using Revised Cochrane risk-of-bias tool (RoB 2). The domains encompassed in RoB 2 analysis cover all types of bias that are currently understood to affect the results of RCTs: (1) bias arising from the randomization process; (2) bias due to deviations from intended interventions; (3) bias due to missing outcome data; (4) bias in measurement of the outcome; and (5) bias in selection of the reported result [16]. Risk-of-bias assessments were visualized using RoB 2 web app [17]. Finally, publication bias was statistically evaluated for efficacy outcomes with more than ten studies [18] by using Egger’s regression test [19], funnel plots, and the Duval & Tweedie’s Trim-and-Fill procedure [20].

### 2.4. Grading the evidence and the impact of COVID-19 pandemic

To evaluate the quality of underlying evidence and the strength of recommendations in this study, we followed Grades of Recommendation, Assessment, Development, and Evaluation (GRADE) approach that focuses on six items: within-study bias, reporting bias, indirectness, imprecision, heterogeneity, and incoherence. In addition, the impact of COVID-19 pandemic on each study was evaluated by summarizing the number (%) of patients affected by COVID-19 disease (Drug vs. Placebo) and how the pandemic affected each stage of the trial, including adverse events and dropouts, missing doses, delays and staffing difficulties, and whether the impact of COVID-19 was analyzed in the main and sensitivity analyses.

### 2.5. Outcomes

Primary outcomes included efficacy measures reported by all or the majority of the original studies. These were mean changes from baseline in cognitive function evaluated by AD Assessment Scale-Cognitive Subscale (ADAS-Cog), Clinical Dementia Rating scale-Sum of Boxes (CDR-SB), and Mini Mental State Examination (MMSE).

Secondary outcomes were biomarker and safety measures reported by all or the majority of studies. Biomarker outcomes included changes from baseline in amyloid burden on PET (centiloid scale), cerebrospinal fluid (CSF) levels of Aβ_1−42_ and p-tau (Thr_181_), and plasma *p-tau*. Safety measures were Serious adverse events, tolerability (treatment discontinuations due to adverse events), total events of ARIA-E (cerebral edema), cerebral microhemorrhages, total events of ARIA-H (including cerebral microhemorrhages, macrohemorrhages and superficial siderosis), headaches, fatigue, dizziness, syncope, nausea, falls, cardiac disorders, back pain, arthralgia, nasopharyngitis, upper respiratory and urinary infections.

Tertiary outcomes included efficacy and safety measures reported by few studies only: functional scale AD Cooperative Study-Activities of Daily Living in MCI (ADCS-ADL-MCI), superficial siderosis (ARIA-H), infusion-related reactions and total events of ARIA-E and ARIA-H in APOE-ε4 non-carriers and carriers (total and depending on the ε4 allele dose).

The results of NMAs were summarized by League Tables with Standardized Mean Differences (SMDs) calculated for efficacy (continuous) outcomes, and Relative Risks (natural logarithm) for safety outcomes.

### 2.6. Data analysis and reporting

The aims of this study were to: (1) update our previous study by performing conventional (frequentist) random-effect meta-analysis with DerSimonian and Laird method and (2) conduct frequentist and Bayesian NMA to assess comparative efficacy and safety of anti-amyloid antibodies. The results obtained from different methods were then compared for each outcome. In order to properly manage the differences among multiple subgroups within the primary studies with respect to differences in drug doses, biomarker findings and other patient characteristics, we followed the recommendation to treat each subgroup as a separate study [21].

#### 2.6.1. Frequentist NMA

Frequentist random-effects NMA was conducted using the *netmeta* package [22] in R (version 4.2.2). The values of I^2^ statistic and their statistical significance were evaluated for overall heterogeneity or inconsistency in the network, which was then split into within-design (i.e., heterogeneity) and between-design (i.e., inconsistency) and the statistical significance of Q-values was assessed. For each outcome, the results are presented in the form of league tables and treatment rankings calculated as P-scores by using *netrank* function from *netmeta* package [22–24]. P-score of 1 is the best possible score and 0 is the worst. Further information on this ranking can be found in the Method section of the Supplement.

#### 2.6.2. Bayesian NMA

Bayesian random-effect NMA was performed by using *rjags* and *gemtc* packages in R [25] and JAGS 4.3.0 software [26] that allow automating NMA in the Bayesian hierarchical model framework with uninformative priors and Markov chain Monte Carlo (MCMC) estimation. The number of tuning iterations was set at 10 and the number of simulation iterations at 100,000. Convergence of the MCMC model was assessed through Gelman-Rubin plots and the potential scale reduction factor. Trace and density plots were then applied to determine the best parameters of the number of burn-in iterations, the number actual simulation iterations, and thinning parameter. For each outcome, the results are presented in the form of league tables and rankings are calculated in the form of SUCRA score (surface under the cumulative ranking curve).

This score can be interpreted as the estimated proportion of treatments worse than the treatment of interest (0% ≤ SUCRA ≤ 100%). Values of SUCRA close to 1 (100%) indicate the higher likelihood that specific treatment is the best. To calculate the SUCRA scores in R, we used the *sucra* function from *dmetar* package in R [23]. Further information on SUCRA score can be found in the Supplement.

#### 2.6.3. Influence and Sensitivity Analyses

Influence analyses were done to detect outliers and influential studies in the network. These included heterogeneity assessments using Baujat plots, Influence diagnostics, and Leave-One-Out meta-analysis. Then, graphic display of study heterogeneity (GOSH) plot analysis was used to examine the results of an equal-effects model in the combinatorial meta-analyses with all possible subsets of studies [23, 27]. GOSH plot diagnostics employs three unsupervised machine learning algorithms: (1) k-means clustering based on vector quantization [28], (2) density-based spatial clustering of applications with noise (DBSCAN) [29], and (3) probabilistic Gaussian mixture model (GMM) [30]. Sensitivity analyses were then performed by rerunning frequentist and Bayesian NMAs upon excluding influential observations.

#### 2.6.4. Rank Robustness

To quantify the influence a study had on the SUCRA based treatment ranks, the robustness of ranks was evaluated within the sensitivity analyses, upon excluding all influential studies and during Leave-One-Out meta-analysis. The influence of study elimination on the ranks in the Bayesian NMA was estimated by using quadratic weighted Cohen’s kappa [31], as recently proposed by Daly and colleagues [32]. This allowed us to measure the agreement between the ranks produced with the complete dataset and the ranks produced from sub-datasets generated upon removing influential studies and each study at a time. The magnitude of Cohen’s kappa is usually categorised into levels of agreement for interpretation (eg, poor (<0%), slight (0%–20%), fair (21%40%), moderate (41%–60%), substantial (61%–80%) and almost perfect (81%– 100%) agreement). However, it should be added that a kappa is not an index of accuracy but of agreement beyond chance [33], and that the uncertainty of rankings and other study characteristics that are context-dependent should be incorporated into the interpretation of the results, as a kappa value of 90%, for example, is not always indicative of almost perfect rank agreement [32]. The calculations were performed by using cohen.kappa function available in *psych* package in R [34]. In addition, the robustness of each rank was examined by determining how many (%) studies did not change any treatment rank (%), as well as how many (%) studies displaced treatment ranks.

#### 2.6.5. Rank Uncertainty

The uncertainty of the SUCRA ranks in Bayesian NMA is visualized by rankograms and tables that show the distribution of the ranking probabilities [35–37], as suggested by the PRISMA–NMA guideline and GRADE working group. Furthermore, we measured Shannon’s information entropy, as recommended recently by Wu et al (2021) [38]. The authors suggested very intuitive way to estimate uncertainty associated with ranking probabilities in NMA by applying Shannon’s information entropy formula to obtain a normalised entropy score. Their work showed that the normalised entropy score gives a more accurate assessment of ranking uncertainty and does not depend on the number of analysed treatments. This approach can be used to compare the uncertainty of treatment rankings within a NMA and also between different NMAs, which is crucial for interpretation of results [38].

For a NMA, the most precise scenario would be that we are absolutely certain in the ranking of treatments in our network. Therefore, each treatment would have 100% probability of being in one ranking position and 0% probability for the other positions (peaked distribution). Under this scenario, the entropy is zero bit, and Shannon’s normalized entropy equals zero. On the other hand, the normalized entropy reaches 1 in the least precise scenario when the ranking probabilities are the same for each rank (flat distribution). In this work, we regarded as “low” those values of normalized entropy that were equal or lower than 0.25 and “moderate” those between 0.25 and 0.5. Larger values were considered as conveying “high” or “substantial” uncertainty or normalized entropy associated with ranking probabilities. Further information on normalized entropy score, and the code for calculation are located in the Supplement.

#### 2.6.6. Clinical relevance

Finally, the clinical relevance of the statistically significant results (compared to Placebo) from the frequentist main NMA and sensitivity analyses was assessed by calculating the “Number Needed to Treat” (NNT) or “Number Needed to Harm” (NNH) from the treatment’s pooled effect size (Hedges g’) by Area Under the Curve (AUC) method [39]. Here, AUC is defined as the probability that a patient receiving the treatment has an outcome preferable to one receiving Placebo. Lower NNT and higher NNH values are associated with a more favorable benefit/risk profile. The best NNT is 1, indicating that for every patient treated one got better, while NNT > 1, means that the fewer people will benefit the treatment [23]. Similarly, NNH of 1 means that for every patient treated one got harmed, while NNH > 1 suggests that fewer people will be harmed.

## 2. RESULTS

The results obtained from the conventional (frequentist) meta-analysis with DerSimonian-Laird estimator were very similar to the Frequentist and Bayesian NMA results. Due to word limitation and large consistency in the findings, this manuscript will only refer to the results obtained from the NMAs. Conventional meta-analysis will be reproducible online in AlzMeta.app (https://alzmetaapp.shinyapps.io/alzmeta/), upon publication of this work.

The meta-analysis included 16 studies (11 ClinicalTrials.gov registries), considering different subgroups within certain RCTs as separate studies, according to our methodology [21]. Bapineuzumab was tested in six studies [40, 41], Solanezumab in three studies [42, 43] Aducanumab in four studies ([44]), Donanemab in two studies (low/medium-tau and high-tau populations from TRAILBLAZER-ALZ 2 [8], and Lecanemab in one study [45]. The details concerning the quality of reports from primary studies (modified Jadad scale) can be found in the Supplement (Supplementary Tables A1 and A2), as well as the number of records identified, included and excluded, and the overall reasons for exclusions (PRISMA Flow Diagram, Supplement), along with PRISMA Checklist (Supplement). Specific reasons for excluding each primary study are listed in Supplementary Tables A3 and A4 (Supplement).

In total, 16,971 patients were analyzed in the primary studies, 8,447 (mean age 71,43 years) received Placebo, while 8,524 (mean age 71,87) were passively immunized against Aβ. All treatments lasted either 76 or 78 weeks across multiple centers, with the average number of trial sites 212.27 (median 198). Acetylcholinesterase inhibitors and memantine were allowed during the trials, alone or in combination. Additional baseline participant characteristics and the inclusion criteria for primary studies are documented in the Supplement (Table A5 and A6).

Statistical power was 100% assuming moderate between-studies heterogeneity and any possible effect size (small, medium, large) (Fig. S1, Supplement). Regarding risk of bias, all studies were considered as either “low risk” or raising “some concerns” (Fig. S2a,b and Table A7; Supplement). For each outcome, certainty of evidence is rated low or very low within each comparison, according to GRADE approach (Table A8; Supplement), with low number of studies being the most important reason for downgrading the certainty.

### 2.1. Primary outcomes

Frequentist NMA results showed that Donanemab, Lecanemab and Aducanumab were significantly more effective than Placebo on ADAS-Cog (cognitive scale, Table 1A). Bapineuzumab and Solanezumab had no favorable effects, with frequentist (but not Bayesian) NMA results suggesting that Placebo was superior to Solanezumab on this scale. In frequentist framework, Donanemab was significantly more effective than Bapineuzumab and Solanezumab and had a double effect size compared to that achieved with Aducanumab and Lecanemab. Bayesian NMA results revealed strong evidence that Aducanumab and Donanemab improved ADAS-Cog and the lack of convincing support for the effects of Lecanemab. Furthermore, in Bayesian framework, we found strong indication that Donanemab outperformed Bapineuzumab, but not Solanezumab, which was clear from the credible intervals including zero (Table 1B).

**Table 1.**
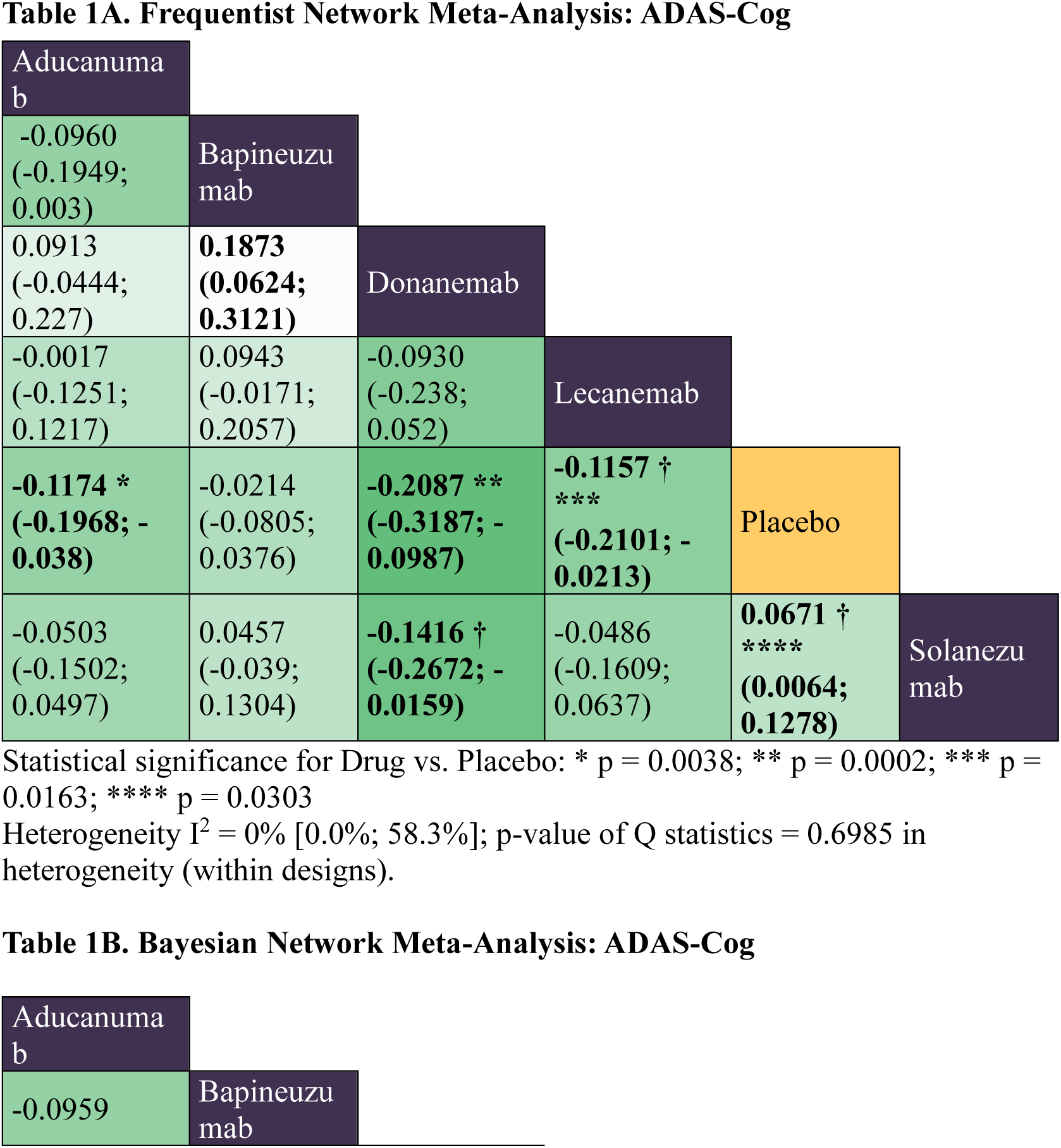
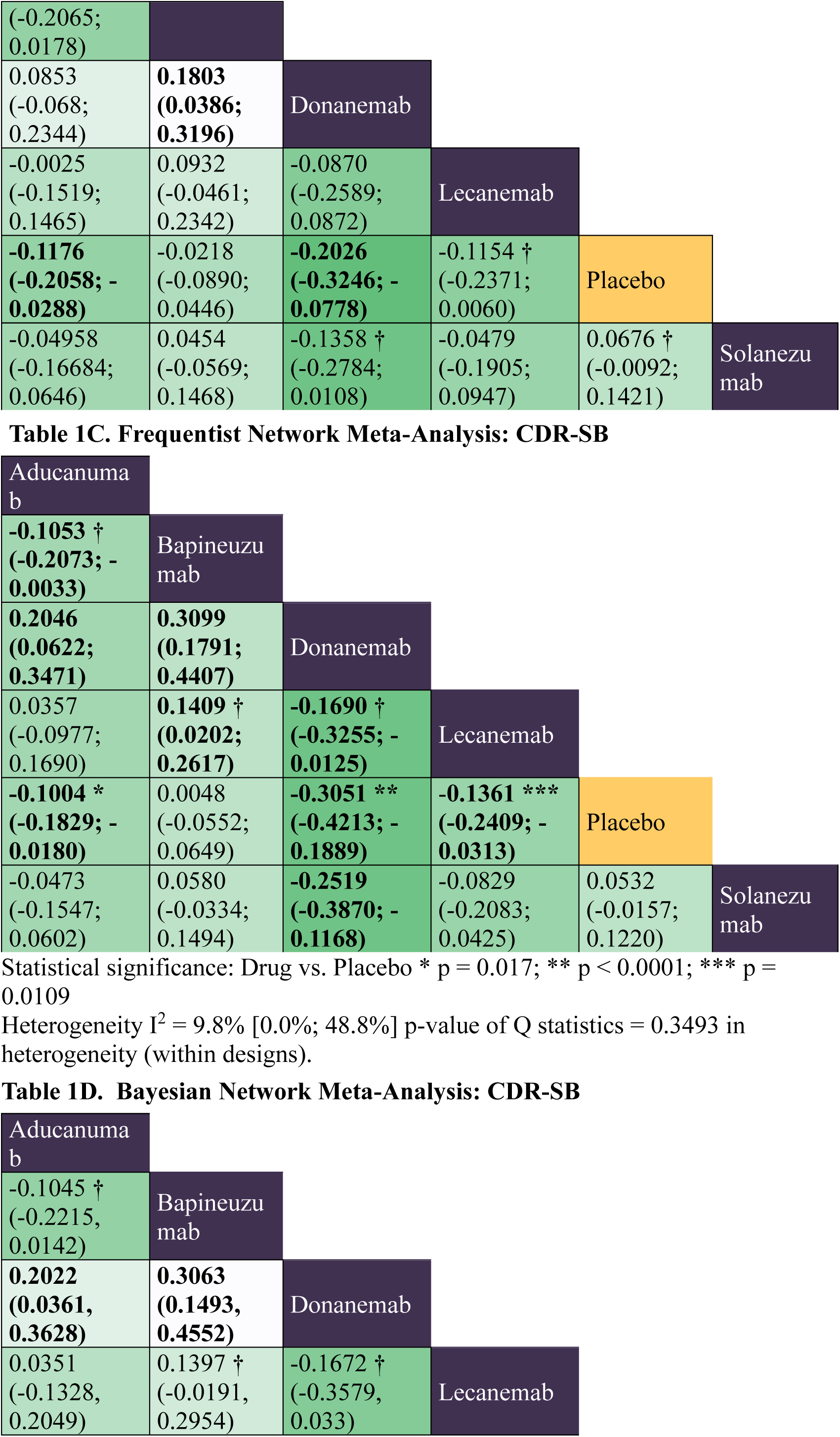
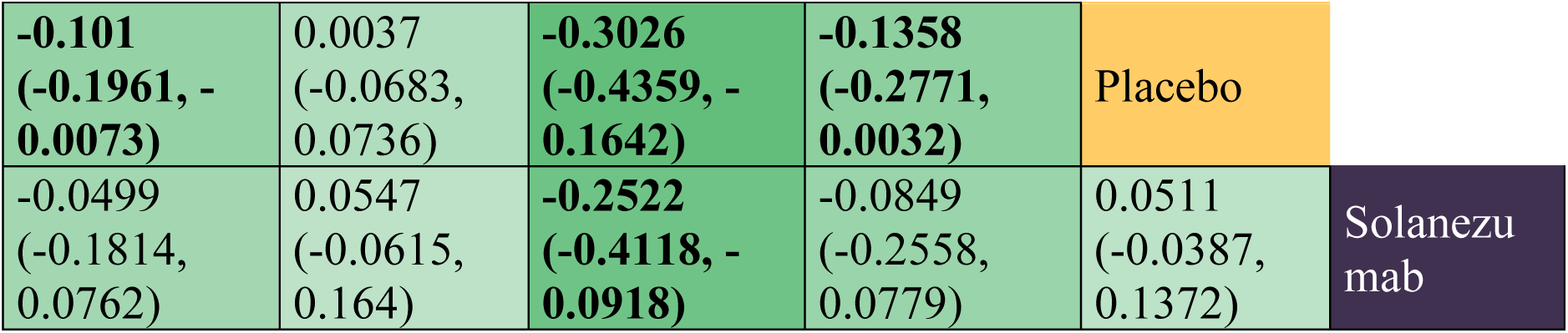
League tables representing the results of random-effect model Frequentist and Bayesian network meta-analysis (NMA). The values in the tables represent Standardized Mean Differences (SMDs) for cognitive outcome Alzheimer’s Disease Assessment Scale-Cognitive sub-scale (ADAS-Cog, **A**, **B**) and for global (cognitive and functional) outcome Clinical Dementia Rating Sum of Boxes (CDR-SB, **C**, **D**). The number below 0 suggests that the drug in the column is superior to the drug in the row. Darker green colors correspond to relatively greater effects, and bolded numbers denote statistical significance (**A, C**) or strong evidence (**B**, **D**). The numbers in parentheses represent confidence intervals (CIs) in Frequentist NMA (**A**, **C**) or credible intervals (CrIs) in Bayesian framework (**B**, **D**). Dagger symbol (**†**) denotes that the interpretation differs considerably between frequentist and Bayesian NMA results, indicating the uncertainty and lack of strong evidence for a non-zero effect.

Even better results were obtained on cognitive and functional (global) scale CDR-SB (Table 1C), where Donanemab, Lecanemab and Aducanumab were significantly more effective than Placebo. Donanemab was more effective than other interventions with nearly three times greater effect size than that of the approved antibodies. Aducanumab and Lecanemab were significantly more effective than Bapineuzumab, however, with the confidence intervals very close to zero. Bayesian NMA produced very similar results as frequentist NMA on CDR-SB, with the exception of three indirect comparisons (Donanemab vs. Lecanemab; Lecanemab vs Bapineuzumab and Aducanumab vs Bapineuzumab), where credible intervals included zero, suggesting the lack of strong evidence for the non-zero effect in these comparisons (Table 1D).

Frequentist NMA results for MMSE (cognitive) score revealed weaker effects of all drugs in the network, with Donanemab and Solanezumab significantly more effective than Placebo (MMSE results for Lecanemab were not reported). These results were fully validated by Bayesian NMA (Table S1; Supplement).

Heterogeneity was not significant for any primary outcome, and inconsistency was not assessed because the network did not have a closed loop. We found no evidence of publication bias, with the Egger’s test p-values 0.661, 0.8462, and 0.8929 for ADAS-Cog, CDR-SB, and MMSE, respectively. Funnel plots were symmetric for all three outcomes (Fig. S3a-c in the Supplement), and the imputation of potentially “missing studies” did not change results.

Across all primary outcomes, Donanemab was the best-ranked antibody according to both frequentist P-scores [24] and SUCRA scores [35] in Bayesian framework. On CDR-SB, Donanemab obtained the maximum scores (near 1) and very low Shannon’s normalized (information) entropy score [38]. This suggests high certainty in the ranking probabilities for Donanemab on CDR-SB. In contrast, on ADAS-Cog and MMSE, SUCRA scores for Donanemab were 0.93 and 0.89 respectively, with moderate uncertainty in the ranking probabilities (Fig. 1C and Table P1 in the Supplement).

**Figure 1.**
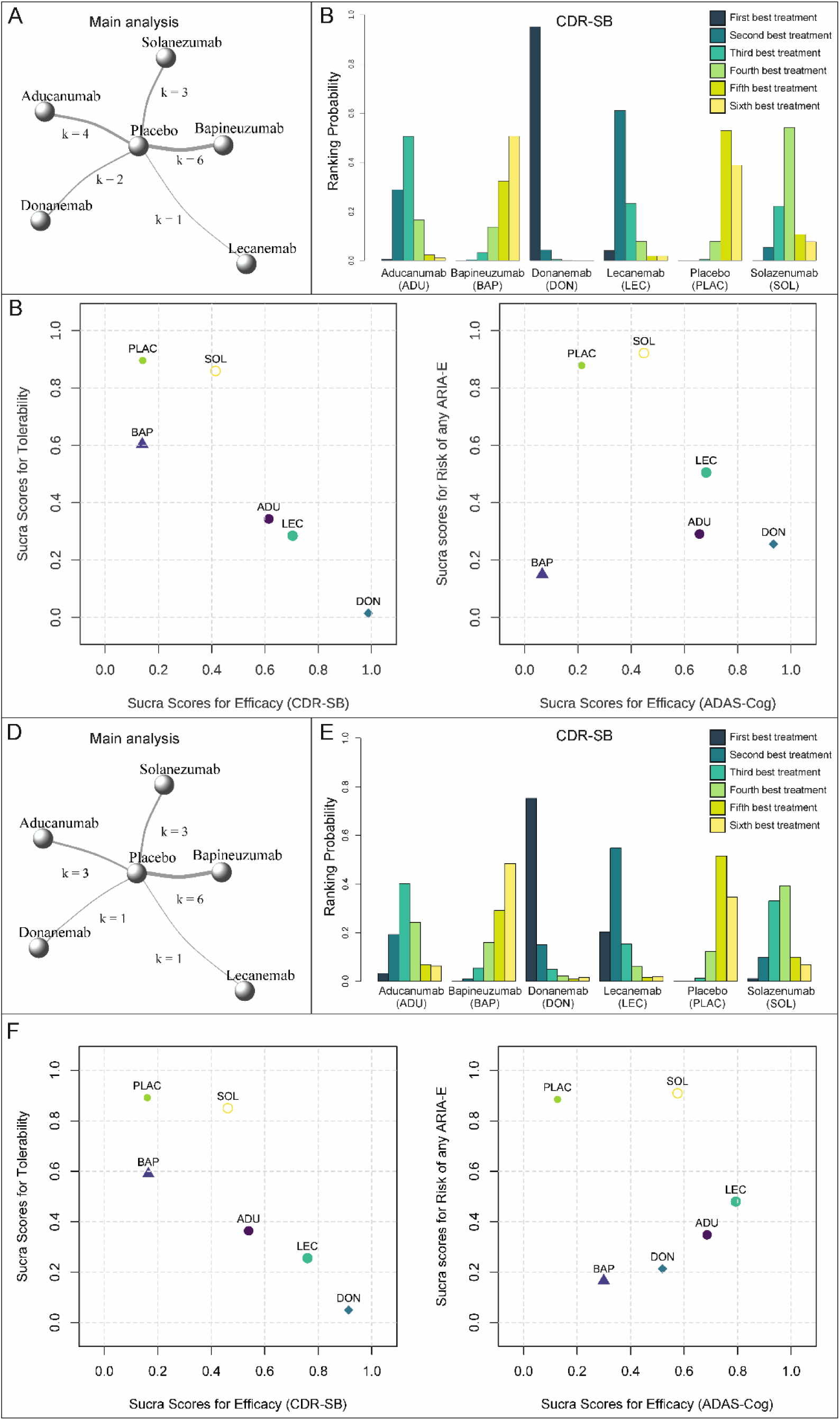
The results of Bayesian NMA for efficacy and safety of anti-amyloid antibodies in Alzheimer’s disease in phase III randomized placebo-controlled clinical trials. **A**, **B** and **C** correspond to the main analysis with all studies included, and **D**, **E** and **F** refer to the sensitivity analysis with two influential studies left out: Donanemab study in low/medium-*tau* population and high-dose EMERGE study of Aducanumab. **A, D.** Network models with nodes (circles) representing antibodies, and the width of the edges or links (k) indicating the number of studies included for each comparison. **B, E.** Rankogram for global (cognitive and functional) outcome CDR-SB. The Y axis represents the probability for each treatment being the best option, second best option, third best option etc. (legend). **C, F** Scatterplots showing jointly the ranking results for efficacy and safety of drugs in the network. SUCRA scores were calculated for the main efficacy outcomes (X-axes: CDR-SB and ADAS-Cog) and safety measures (Y-axes: tolerability and risk of any ARIA-E). Abbreviations: ARIA-E, Amyloid-Related Imaging Abnormalities in form of cerebral Edema; CDR-SB, Clinical Dementia Rating Sum of Boxes; SUCRA, Surface Under Cumulative RAnking curve.

Donanemab produced clinically the most relevant effects on all primary outcomes in combined low/medium population and high-*tau* population, with NNT = 9 on ADAS-Cog, 6 on CDR-SB and 13 on MMSE. When Donanemab was compared to Aducanumab and Lecanemab, the most striking difference was observed on CDR-SB, with NNT values almost two times higher for Lecanemab (13) and almost three times higher for Aducanumab (18). On ADAS-Cog, Lecanemab and Aducanumab also had high NNT (15). Finally, Solanezumab had high NNT (20) on MMSE, suggesting low clinical relevance of cognitive effects of this drug.

### 2.2. Secondary outcomes

#### 2.2.1. Biomarkers of Aβ and *tau*

Both Frequentist and Bayesian NMA results showed that Lecanemab, Donanemab, and Aducanumab significantly improved clearance of Αβ deposits from brain when compared to Placebo (Table S2.1; Supplement). Lecanemab demonstrated the greatest amyloid-clearance activity on PET and obtained P- and SUCRA scores very close to 1, with high certainty (low information entropy) associated with the ranking probabilities (Table P1 in the Supplement). However, despite large effect-sizes, these data should be interpreted with caution, as substantial heterogeneity was found between the studies and the number of analyzed trials/patients was small. While frequentist NMA results indicated significant differences between Lecanemab and other antibodies, Bayesian NMA results revealed a lack of strong evidence that Lecanemab outperformed Aducanumab and Donanemab on this outcome.

Frequentist NMA results revealed that Aducanumab and Lecanemab were significantly superior to Placebo on CSF biomarkers of Aβ_1-42_ and p-181-*tau* (data for Donanemab were not reported). Aducanumab significantly outperformed Solanezumab on CSF Aβ (Table S2.1) and Bapineuzumab on CSF p-*tau* (Table S2.2.). Bayesian NMA validated the relative effect of Aducanumab on both CSF Aβ and p-*tau*, providing strong evidence in favor of positive effects of Aducanumab on these outcomes when compared to Placebo, but not when compared to other drugs. Furthermore, Bayesian NMA results revealed an absence of strong evidence that Lecanemab was superior to Placebo on CSF Aβ and p-*tau*. Aducanumab was ranked the best treatment on both CSF biomarkers of Aβ and p-*tau*, with SUCRA scores 0.86 and 0.89, respectively, and moderate uncertainty in the ranking probabilities (Table P1; Supplement).

As in case with amyloid PET data, these findings should be interpreted carefully, since CSF biomarker analyses were available only for limited number of patients in few studies, and considerable heterogeneity was found for CSF Aβ (I^2^ = 68.9% [20.0%; 87.9%]; p-value of Q statistics = 0.012), indicating that the true effect size may vary across studies. Heterogeneity was not significant for CSF p*-tau* (Table S2.2; Supplement), while the analysis of plasma *tau* biomarkers had to be dropped because of extreme heterogeneity (I^2^ = 99% [98.5%; 99.3%], p < 0.0001) and differences in measured biomarker outcome.

All biomarker effects were clinically relevant for Aducanumab, Lecanemab and Donanemab, with NNT values of 1 for the effects on amyloid burden (PET data), indicating that the favorable effect on amyloid removal was accomplished for each treated patient, with each of these antibodies. NNTs for CSF outcomes were also very small (clinically relevant) for Aducanumab and Lecanemab, with NNT = 2 on CSF Aβ_1-42_ for both antibodies. As for CSF-p*-tau*, the NNT of Aducanumab (3) was somewhat lower than that of Lecanemab (4), reflecting slightly greater effect of Aducanumab on p-*tau*.

#### 2.2.2. Safety outcomes

Anti-amyloid antibodies did not increase the risk of serious adverse events (Table S3.1; Supplement). Nevertheless, both the frequentist and Bayesian NMA results provide substantial evidence that high-clearance antibodies (Donanemab, Lecanemab and Aducanumab) were less tolerable than Placebo and Solanezumab since these drugs greatly increased the risk of treatment discontinuation due to adverse events (Table 2). Donanemab was the least tolerable antibody in the network, significantly worse than Bapineuzumab and Aducanumab. Frequentist NMA results revealed considerable risks of discontinuation for Bapineuzumab when compared to Placebo, however, this was not corroborated in Bayesian analysis.

**Table 2.**
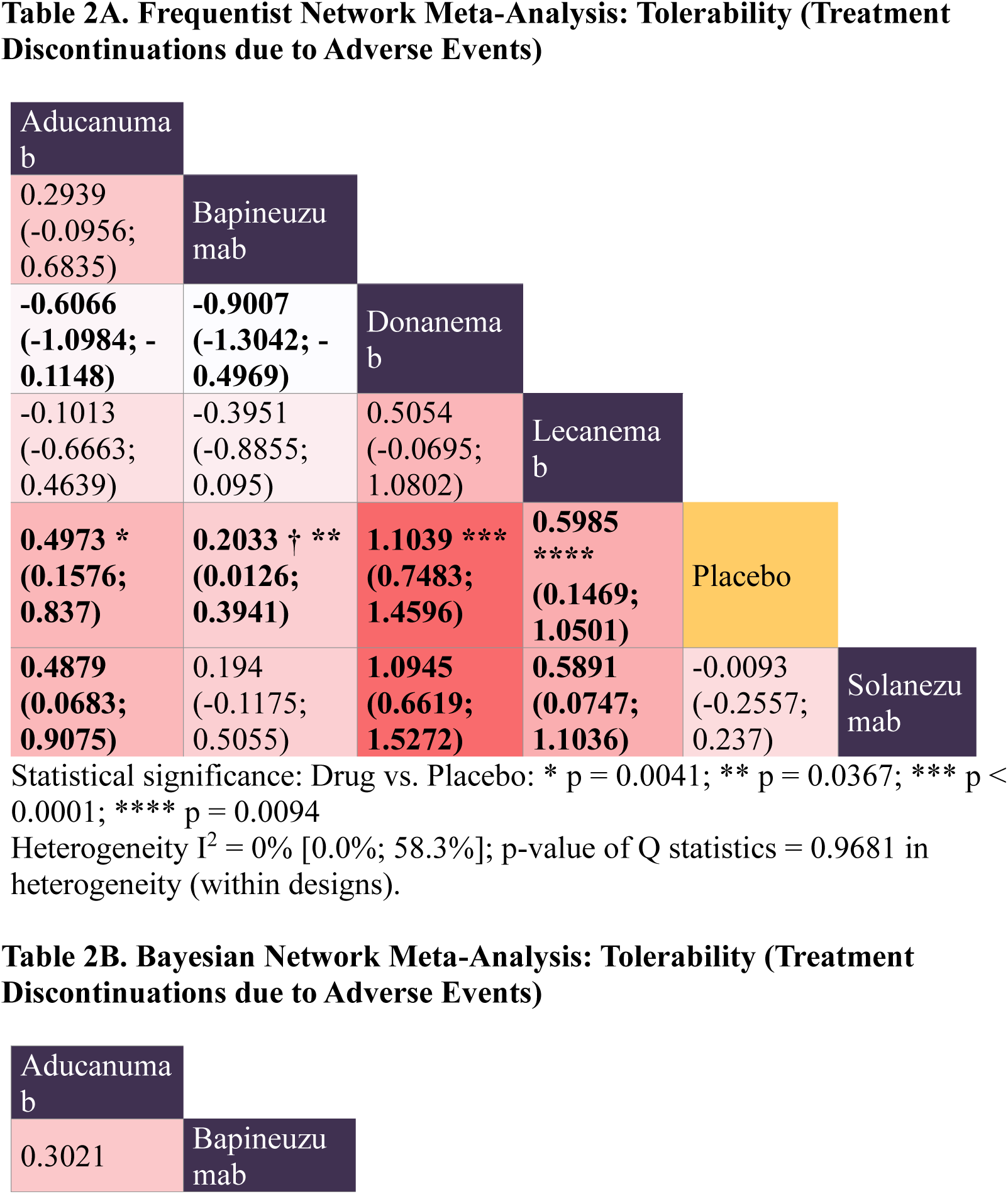
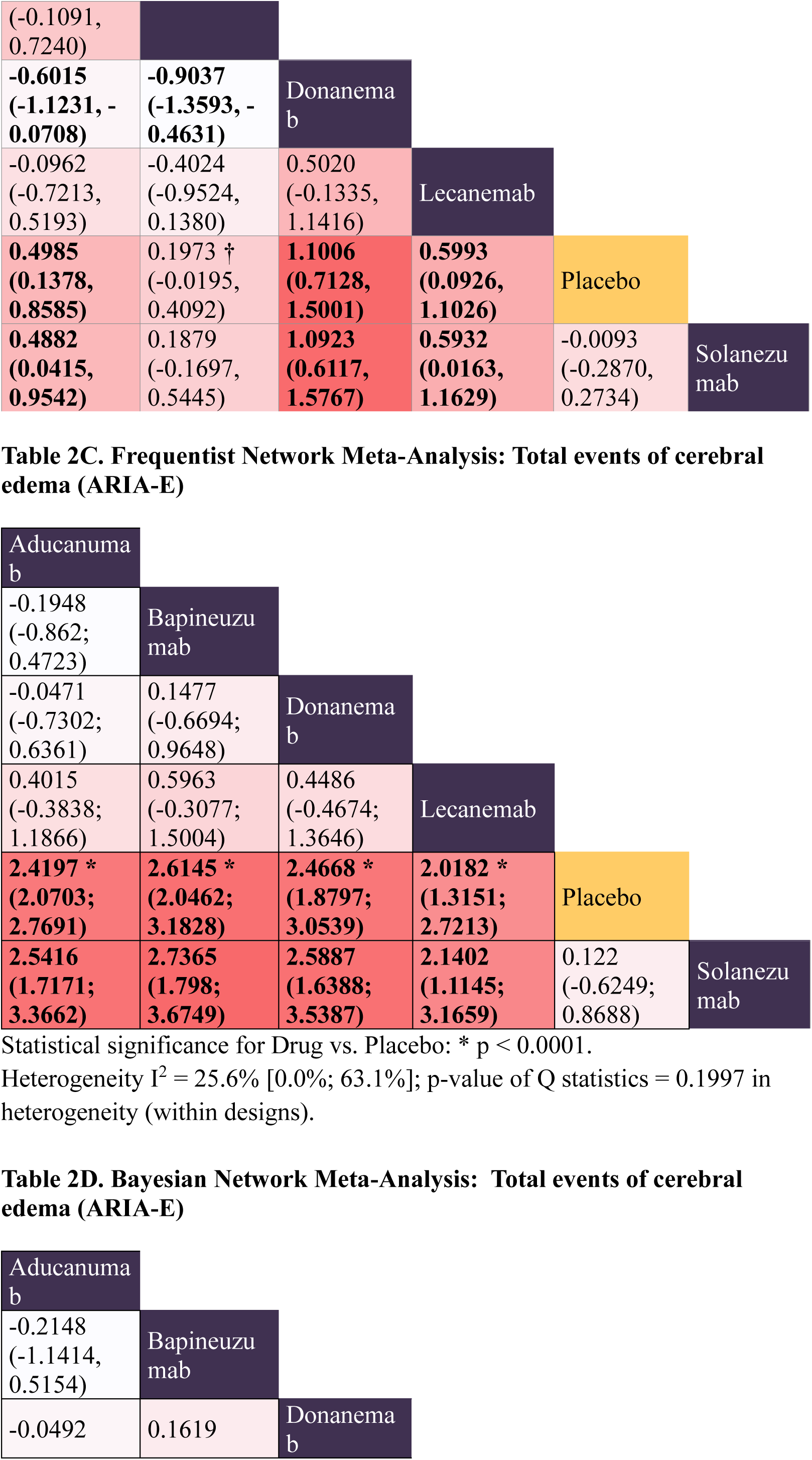
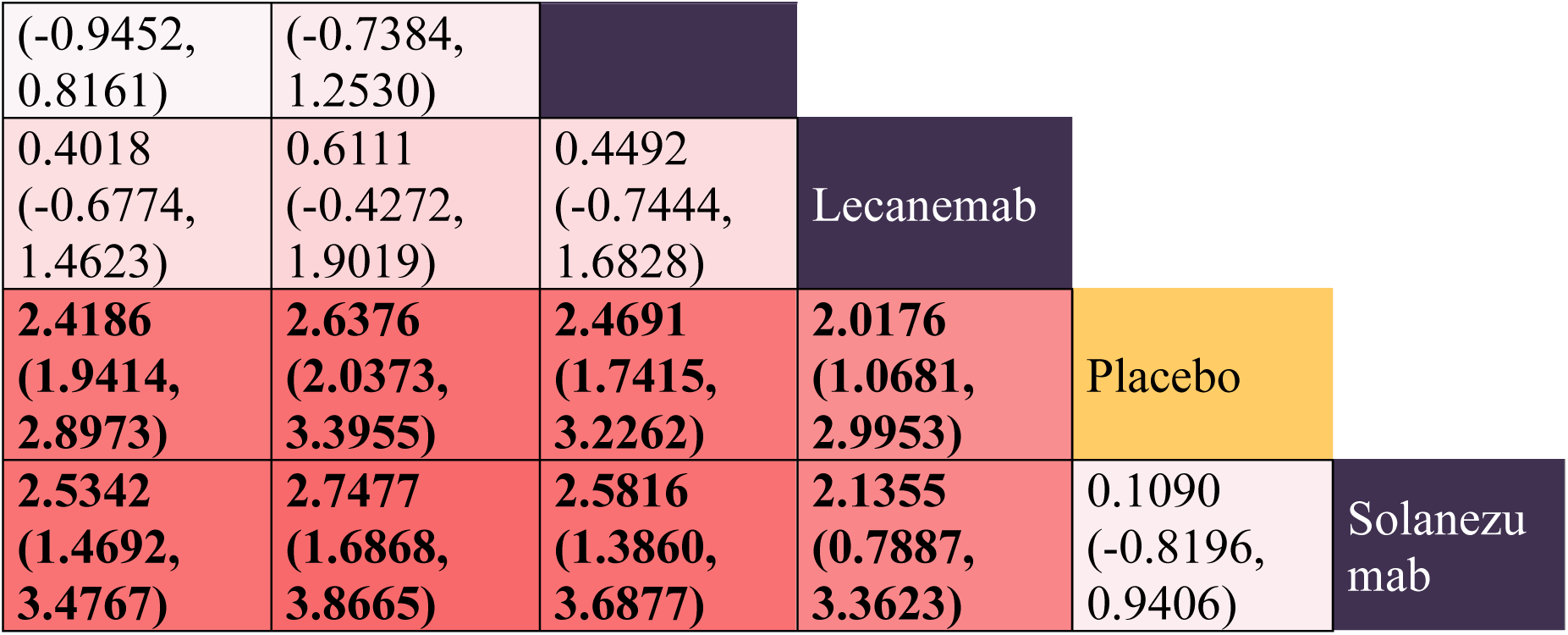
League tables representing the results of random-effect model Frequentist NMA and Bayesian NMA. The values in the tables represent Relative Risks (natural logarithm) for tolerability (treatment discontinuations due to adverse events, **A**, **B**) and for amyloid-related imaging abnormalities in form of cerebral edema (ARIA-E, **C**, **D**). For each comparison, the number below 0 suggests that the drug in the column is safer than the drug in the row. Darker red colors correspond to relatively higher risk. Bolded numbers indicate statistical significance for Frequentist NMA (**A**, **C**), or strong evidence in Bayesian NMA (**B**, **D**). The numbers in parentheses represent confidence intervals (CIs) for Frequentist NMA (**A**, **C**), or credible intervals (CrIs) in Bayesian framework (**B**, **D**). Dagger symbol (**†**) denotes when the interpretation differs significantly between frequentist and Bayesian NMA results, indicating the uncertainty and lack of strong evidence for a non-zero risk.

Frequentist and Bayesian NMA results provide compelling evidence that Solanezumab was the most tolerable and the safest antibody in terms of ARIAs (Fig. 1C, Table 2). Other antibodies substantially increased the total risk of ARIA-E and the risk of cerebral microhemorrhages (ARIA-H), when compared to Placebo and Solanezumab. Furthermore, Aducanumab, Donanemab and Lecanemab significantly increased the total risk of ARIA-H (including all the events of cerebral superficial siderosis, micro- and macrohemorrhages), when compared to Solanezumab and Placebo (Tables S3.2 and S3.3 in the Supplement)

Frequentist NMA on additional safety outcomes showed that Aducanumab significantly increased the risk of headaches (Table S3.4) and falls (Table S3.5; Supplement) when compared to Placebo, however, with confidence intervals very close to zero and substantial heterogeneity between the studies (see 3.6). Further Bayesian NMA revealed lack of conclusive support for these comparisons. Similarly, in Bayesian framework, there was an absence of strong evidence that Donanemab increased the risk of nausea in high-*tau* population, when compared to Placebo, whereas frequentist NMA would suggest significant difference in risk (Table S3.6; Supplement).

Further analyses showed that anti-Aβ did not raise the risks of fatigue, dizziness, arthralgia, back pain, cardiac disorders, diarrhea, nor urinary infections more than Placebo (Table S3.7-S3.16; Supplement). In addition, we found no improved risk of syncope, nasopharyngitis and upper respiratory infections, however, these events were not reported for Donanemab and Lecanemab, antibodies tested in clinical trials affected by COVID-19 pandemic (see 2.4 section). Heterogeneity was not significant for secondary safety outcomes with the exception of risks of headaches and cardiac disorders.

The risks of treatment discontinuation due to adverse events, total ARIA-E, cerebral microhemorrhages, and total ARIA-H were all clinically relevant, with NNH values of 1, which correspond to the situation in which for each patient treated, one exhibited treatment-emergent adverse reaction.

### 2.3. Tertiary outcomes

Frequentist NMA of ADCS-MCI-ADL showed that Aducanumab (NNT = 12) and Lecanemab (NNT = 10) significantly slowed functional decline by small effect size, when compared to Placebo. Bayesian NMA results validated these results for Aducanumab, but not for Lecanemab (Table S4.1). The heterogeneity was not significant; however, these results should be interpreted with care since they are based on four studies with Aducanumab and one study testing Lecanemab. Furthermore, credible intervals for the relative effects of Aducanumab (vs. Placebo) were very close to zero, indicating uncertainty in the evidence (also, see 2.4).

Both frequentist and Bayesian NMA results gave strong support for increased risk of superficial siderosis (ARIA-H) in patients treated with Donanemab and Aducanumab, when compared to those in control group, while the results for Lecanemab were not conclusive. Whereas frequentist NMA results suggested significantly greater risk (p = 0.001) of superficial siderosis in patients treated with Lecanemab (compared to Placebo), Bayesian NMA gave an estimate with overlapping credible intervals, suggesting the lack of strong evidence for this comparison (Table S4.2). Even so, the risk of superficial siderosis was clinically relevant for Donanemab, Aducanumab and Lecanemab, with NNH values of 1 (the least favorable).

The risk of infusion-related reactions was clinically meaningful (NNH = 1) for Donanemab and Lecanemab (not reported for other drugs), significantly higher (p < 0.0001) when compared to Placebo. Both the Frequentist and Bayesian NMA results provided strong evidence that Donanemab increased the risks of infusion-related reactions, with almost two times greater risk than that observed in Lecanemab study (Table S4.3 in the Supplement).

Having in mind the limited information regarding ARIA-E and ARIA-H depending on the APOE status, frequentist and Bayesian NMAs strongly indicate that all antibodies, except Solanezumab, significantly increased the risk of ARIA-E in both APOE-ε4 carriers and non-carriers. The results were not that convincing for ARIA-H, where smaller risks were observed and frequentist NMA results suggested significant increase in risk for Donanemab and Lecanemab in both carriers and non-carriers, which was not validated in Bayesian framework. Due to inconclusive findings on ARIAs depending on APOE status, we were unable to determine whether the risk of ARIAs increases substantially with ε4 allele dose (homozygotes vs. heterozygotes). Overall, the risk of ARIAs was clinically relevant (NNH = 1) independently of APOE status, and the risk generally increased with each ε4 allele (Tables S4.4-S4.7 in the Supplement). Heterogeneity between the studies could not be assessed due to small numbers of studies.

### 2.4. Sensitivity analyses

Donanemab consistently showed greater cognitive effects in patients with low/medium *tau* load than in high-*tau* population (assessed by PET imaging), and removing this study had a big impact on the results, similarly to the greater effect observed for high-dose Aducanumab in controversial EMERGE study, as found in our previous meta-analysis [46] (Tables S5 and S7 in the Supplement). For this reason, removing these two studies from the NMAs yielded much lower relative effects of these drugs on cognitive measures (when compared to Placebo), with p-values of 0.0502 and 0.5334 on ADAS-Cog and 0.1419 and 0.0249 on CDR-SB for Aducanumab and Donanemab, respectively. Likewise, excluding study in low/medium-*tau* population greatly reduced relative effects of Donanemab on MMSE (p = 0.3430), further emphasizing favorable effects of this drug in patients with low/medium *tau*, but not high *tau* burden. Bayesian NMA validated these findings, strongly indicating that CDR-SB score was improved by Donanemab (even in high *tau* population), but not by Aducanumab, after excluding controversial EMERGE study. There was lack of strong evidence for improving both ADAS-Cog and MMSE scores neither by Aducanumab nor by Donanemab after removing their influential studies. Furthermore, frequentist NMA results of amyloid PET data revealed that both Donanemab and Aducanumab significantly removed amyloid brain burden (Table S7.4), when compared to Placebo. Conversely, the results from Bayesian NMA provided strong evidence supporting the efficacy of Aducanumab, but not Donanemab. In other words, comparatively higher effect of Donanemab was obtained in low/medium-*tau* population, and removing this measurement increased the uncertainty of the evidence, shifting the credible intervals for the relative efficacy of Donanemab from (−6.4251; −2.0495) towards (−9.3948; 1.7020). Finally, frequentist NMA results of ADCS-ADL-MCI showed that removing high-dose Aducanumab EMERGE trial did not impact the relative effect of this drug when compared to Placebo (Table S7.7). Bayesian NMA weakly favored Aducanumab over Placebo, however, with credible intervals (−0.014; 0.245) including zero. In summary, significant dose-dependent efficacy of Aducanumab was evident only in EMERGE study, and the relative effects of Donanemab in the main study were largely due to its beneficial effects on AD patients with low/medium *tau* load.

Removing the influential studies of Donanemab and Aducanumab reduced ranking scores of these drugs in the network (Figure 1F). On MMSE score, for example, Solanezumab was the best-ranked drug with SUCRA score 0.81, followed by Donanemab in high-*tau* population (0.71). Lecanemab obtained the best score (0.79) on ADAS-Cog, followed by Aducanumab (0.69), and Solanezumab (0.58). Donanemab in high-*tau* population was the best-ranked treatment only on CDR-SB score (0.91) and fourth-ranked on ADAS-Cog (0.3). Additionally, Lecanemab was better ranked treatment than Aducanumab on CSF-Aβ_1-42_ and ADCS-ADL-MCI, upon excluding the influential effects from high-dose Aducanumab in EMERGE study. For efficacy outcomes, the most robust SUCRA scores were obtained for CDR-SB, with 92% of agreement with the main analysis (median of observed weighted Cohen’s kappa values) and 8 (50%) studies that did not change any treatment rank. The robustness of SUCRA scores further improved upon excluding influential Aducanumab and Donanemab studies, however, the uncertainty in ranking probabilities also increased. The least robust SUCRA scores were found for amyloid burden on PET, which was a direct consequence of small number of studies. Across all efficacy outcomes, the treatments moved only one or very rarely two ranks up or down upon excluding each study at a time. Further details on ranking probabilities and their sensitivity analysis can be found in the Supplement (Tables P1-P4; Supplement).

Clinical relevance of the cognitive and functional effects of Donanemab and Aducanumab decreased remarkably upon removing their influential studies. while the biomarker effects on Aβ and *p-tau* remained clinically relevant with small NNTs. For Donanemab, the NNT was 28 for ADAS-Cog and 18 for MMSE, while NNT for CDR-SB did not change drastically (NNT = 8), when compared to the main analysis (NNT = 6). In other words, the beneficial effects of Donanemab on cognitive and functional CDR-SB score in patients with high*-tau* load were still clinically relevant, even if the effects were greater in those with low/medium *tau*. Aducanumab had higher (less favorable) NNTs in sensitivity analysis for ADAS-Cog (21) and CDR-SB (19).

The results of sensitivity analyses on safety outcomes were largely consistent with the main analysis. SUCRA scores for treatment discontinuation, risks of ARIA-E and ARIA-H were robust to study exclusions, with median weighted Cohen’s kappa values above 89%, indicating almost perfect agreement in study ranks between main and sensitivity analysis (Tables P1-P4 in the Supplement). Additionally, there was small statistically significant increase in the risk of headaches for all high-clearance antibodies: Aducanumab (p = 0.0001), Donanemab (p = 0.0085), and Lecanemab (p = 0.0323), when compared to Placebo (Table S2.9), after excluding two low-dose Bapineuzumab studies [41]. These two studies were the source of high heterogeneity in the main analysis, which was detected by Q statistics (p < 0.0001) and Gaussian Mixture Model in GOSH plot analysis (Table S8.1 and Figure S11 in the Supplement). Bayesian NMA validated frequentist NMA results for Aducanumab and Donanemab, suggesting that these antibodies slightly increased the risk of headaches when compared to Placebo and Solanezumab. On the other hand, there was no conclusive evidence that Lecanemab elevated risk of headaches, when compared to Placebo. Additionally, the statistical significance for the risk of falls in Aducanumab studies dropped (p = 0.8785) once we removed ENGAGE studies (Table S8.2 in the Supplement), suggesting that these studies contributed to the observed high risk of falls in the main analysis.

### 2.5. The Impact of COVID-19 pandemic

Lecanemab and Donanemab RCTs included in this study were affected by global outbreak of coronavirus – an infectious disease caused by the severe acute respiratory syndrome. COVID-19 was the most commonly reported adverse event across treatment groups in Donanemab studies, ocurring in ∼17% of patients in all arms. The impact of pandemic was less severe on Lecanemab study, with the incidence of COVID-19 around 7% in experimental and control groups. The pandemic also caused delays in study visits and assessments in both trials and other difficulties summarized in the Supplement (Table S6).

## 3. DISCUSSION

This study reveals a complex pattern of evidence supporting the efficacy of Aducanumab, Lecanemab and Donanemab across multiple cognitive, functional and biomarker outcomes. At the same time, the risks of cerebral edema and microbleeding were substantial, suggesting greater harm than benefit. The similarities and the differences between frequentist and Bayesian NMA results presented here strongly emphasize the need for further studies with larger sample sizes and the need to study these antibodies in earlier stages of AD and MCI, and in younger patients with lower Aβ and *tau* load.

The inconsistencies between ENGAGE and EMERGE trials revealed here and in our previous study [46] have been discussed in literature ever since these studies were published, as they caused a lot of controversy within the FDA and in the broader community [47]. The Aducanumab provider, in partial collaboration with the FDA, provided two possible explanations for these discrepancies: (1) fewer numbers of patients in the ENGAGE trial receiving higher doses of drug, and (2) ENGAGE trials had more outliers, and removing these outliers in sensitivity analysis gave more compatible dose-dependent beneficial effects [48, 49]. Despite these inconsistencies, Aducanumab received accelerated approval in the U.S. Afterwards, the FDA granted breakthrough status to Lecanemab and Donanemab, in order to speed up their approval [50], which finally resulted in full FDA approval of Lecanemab in the U.S. and more recently in Japan. Furthermore, Lecanemab is currently undergoing regulatory review in the European Union (EU), United Kingdom, South Korea, and Canada, while Donanemab is reviewed in the EU and the U.S [7]. Recent exploratory *post hoc* modeling based on phase III RCT of Donanemab suggested that Aβ levels in immunized patients would remain below the positivity threshold for almost 4 years without the treatment. The authors also revealed that greater plaque removal by Donanemab was associated with slower progression of *tau* PET and slower clinical decline, but only in APOE-ε4 carriers [51]. In our study, all high-clearance antibodies (Aducanumab, Donanemab and Lecanemab) produced substantial Aβ and *tau* biomarker effects.

Donanemab was the best-ranked treatment across all cognitive measures, while Lecanemab was the most efficient at reducing amyloid burden on PET. Donanemab consistently showed better cognitive, functional and biomarker effects in patients with low/medium *tau* load, compared to those with high *tau*. Donanemab produced clinically relevant cognitive effects in low/medium-*tau* population on all three primary outcomes, suggesting greater benefit of Donanemab in earlier AD stages.

Low tolerance and high risk of ARIAs looms over high-clearance anti-Aβ antibodies. Our results showed that Donanemab was significantly less safe and less tolerable than Placebo and Solanezumab, with similar risks of treatment discontinuation and ARIAs as in Aducanumab and Lecanemab studies. Both frequentist and Bayesian NMA results from this study imply that the risk of ARIAs associated with the treatment may outweigh the benefits, independently of APOE status. In summary, our results demonstrate modest cognitive and substantial biomarker effects of Donanemab, Lecanemab and Aducanumab, yet with significant trade-off in the form of dose-dependent cerebral vasogenic edema (ARIA-E), bleeding and hemosiderin deposits (ARIA-H), which may or may not be symptomatic or serious adverse event. These risks require careful dose titration and close patient monitoring on Magnetic Resonance Imaging (MRI), which increases the costs of already expensive treatments [52]. In our view, additional and comparative studies of these drugs with vascular biomarkers are needed to evaluate their risk/benefit profile, as well as more anticipatory policies from regulatory bodies if we were to avoid new controversies as those seen with the FDA approval of Aducanumab. Moreover, personalized approach and safer multi-target therapies in combination with novel biomarkers is expected to effectively mitigate the AD progression [53].

Different mechanisms of actions of these drugs outlined in the Introduction certainly contribute to the differences in their effects and heterogeneity in our networks. However, there is no doubt that Aducanumab, Lecanemab and Donanemab achieved great biomarker results and modest cognitive effects in 18 months. Therefore, comparative and longitudinal studies are required to fully disclose the impact of anti-Aβ immunotherapy over longer periods of time. Future trials would need to compare the effects of anti-Aβ antibodies in multiple stages of AD and in different populations, as inter-individual differences and comorbidities may be crucial to prevent ARIAs and select patients that could benefit from anti-Aβ immunotherapy. The inflammation and vascular comorbidities are of special concern, as blood-brain barrier disruption and Aβ deposition are central to the development of both AD (with Aβ deposited in brain parenchyma) and cerebral amyloid angiopathy (with Aβ deposition on blood vessel walls). These two pathologies often co-occur and APOE-ε4 status is known to worsen both of them. Risk factors for ARIA are dose-dependent and include receiving anti-Aβ immunotherapy, anticoagulant and thrombolytic medication, carrying APOE-e4 allele, and history of brain microhemorrhages and stroke [54, 55].

The important limitations of this study are (1) COVID-19 impacted Donanemab and Lecanemab trials, (2) the patients in Donanemab study were stratified by their level of the brain protein *tau*; (3) the effects of antibodies on plasma *tau* biomarkers could not be evaluated due to excessive between-study heterogeneity and different measures used in primary studies; (4) potential bias might be introduced due to unmasking, since ARIAs were the most frequent treatment-emergent adverse events of all drugs except Solanezumab, and (5) direct comparison between the antibodies has not been done yet, and our NMA only included studies where treatments were compared with Placebo. NMA is a statistical method to compare more than two interventions simultaneously by combining direct and indirect evidence. When head-to-head trials are not available, as often happens in many research fields, indirect comparisons can be made via a common control arm (Placebo in our case) [56]. Therefore, this study should be viewed as a way to summarize the results from previous phase III RCTs and provide comparative assessment of these drugs based on existing data, until further evidence comes along. The same should be emphasized for the efficacy and safety rankings and their uncertainty measures, as clinical decision making should never be based solely on one or two measures or studies.

## 4. RESEARCH IN CONTEXT

1. Systematic review and meta-analysis: Frequentist and Bayesian network meta-analyses (NMAs) were performed to compare the efficacy and safety of anti-amyloid antibodies in phase III randomized placebo-controlled trials in sporadic AD.
2. Interpretation: Aducanumab, Lecanemab and Donanemab achieved substantial biomarker results and small cognitive effects, and were less tolerable than Placebo and Solanezumab, significantly increasing the risk of ARIAs. Donanemab was the most efficient antibody on the cognitive measures, but it was also the least tolerable, significantly less tolerable than Placebo, Solanezumab, Bapineuzumab and Aducanumab.
3. Future directions: Studying these drugs in earlier stages and in younger patients with lower Aβ and *tau* burden might yield better outcome than that observed in phase III trials. Since ARIAs are the major adverse events limiting the broad use of anti-amyloid antibodies, prevention of vascular damage, better biomarkers and safer, personalized treatment options are needed to improve AD treatment.

## Data Availability

All data produced in the present study are available upon reasonable request to the authors

## ACKNOWLEDGEMENTS

This work was supported by grants PID2020–115823-GB100 funded by MCIN/AEI/10.13039/501100011033, and SBPLY/21/180501/000150 funded by JCCM/ERDF - A way of making Europe, and 2022-GRIN-34354 grant by UCLM/ERDF intramural funding to LJ-D and JDN-L.

## AUTHORS’ CONTRIBUTIONS

JDNL and LJD were responsible for the initial conceptualization, funding acquisition, supervision, and project administration; DJ was responsible for Data curation, Software, Formal analysis, visualization and writing the original draft; SD, JDNL and LJD did the writing – review and editing. All authors read and approved the final manuscript.

## REFERENCES

[1] Holtzman DM, Morris JC, Goate AM. Alzheimer’s disease: the challenge of the second century. Sci Transl Med. 2011;3:77sr1.

[2] Wessels AM, Dennehy EB, Dowsett SA, Dickson SP, Hendrix SB. Meaningful Clinical Changes in Alzheimer Disease Measured With the iADRS and Illustrated Using the Donanemab TRAILBLAZER-ALZ Study Findings. Neurol Clin Pract. 2023;13:e200127.

[3] Söderberg L, Johannesson M, Nygren P, Laudon H, Eriksson F, Osswald G, et al. Lecanemab, Aducanumab, and Gantenerumab - Binding Profiles to Different Forms of Amyloid-Beta Might Explain Efficacy and Side Effects in Clinical Trials for Alzheimer’s Disease. Neurotherapeutics. 2023;20:195–206.

[4] Plotkin SS, Cashman NR. Passive immunotherapies targeting Aβ and tau in Alzheimer’s disease. Neurobiol Dis. 2020;144:105010.

[5] Prins ND, Scheltens P. Treating Alzheimer’s disease with monoclonal antibodies: current status and outlook for the future. Alzheimers Res Ther. 2013;5:56.

[6] Rabinovici GD, La Joie R. Amyloid-Targeting Monoclonal Antibodies for Alzheimer Disease. Jama. 2023;330:507–9.

[7] Cummings J, Osse AML, Cammann D, Powell J, Chen J. Anti-Amyloid Monoclonal Antibodies for the Treatment of Alzheimer’s Disease. BioDrugs. 2024;38:5–22.

[8] Sims JR, Zimmer JA, Evans CD, Lu M, Ardayfio P, Sparks J, et al. Donanemab in Early Symptomatic Alzheimer Disease: The TRAILBLAZER-ALZ 2 Randomized Clinical Trial. Jama. 2023;330:512–27.

[9] Digma LA, Winer JR, Greicius MD. Substantial Doubt Remains about the Efficacy of Anti-Amyloid Antibodies. J Alzheimers Dis. 2024;97:567–72.

[10] Lacorte E, Ancidoni A, Zaccaria V, Remoli G, Tariciotti L, Bellomo G, et al. Safety and Efficacy of Monoclonal Antibodies for Alzheimer’s Disease: A Systematic Review and Meta-Analysis of Published and Unpublished Clinical Trials. J Alzheimers Dis. 2022;87:101–29.

[11] Jeremic D, Navarro-López JD, Jiménez-Díaz L. Efficacy and safety of anti-amyloid-β monoclonal antibodies in current Alzheimer’s disease phase III clinical trials: A systematic review and interactive web app-based meta-analysis. Ageing Res Rev. 2023;90:102012.

[12] Hutton B, Salanti G, Caldwell DM, Chaimani A, Schmid CH, Cameron C, et al. The PRISMA extension statement for reporting of systematic reviews incorporating network meta-analyses of health care interventions: checklist and explanations. Ann Intern Med. 2015;162:777–84.

[13] Clark HD, Wells GA, Huët C, McAlister FA, Salmi LR, Fergusson D, et al. Assessing the quality of randomized trials: reliability of the Jadad scale. Control Clin Trials. 1999;20:448–52.

[14] Olivo SA, Macedo LG, Gadotti IC, Fuentes J, Stanton T, Magee DJ. Scales to assess the quality of randomized controlled trials: a systematic review. Phys Ther. 2008;88:156–75.

[15] Jadad AR, Moore RA, Carroll D, Jenkinson C, Reynolds DJ, Gavaghan DJ, et al. Assessing the quality of reports of randomized clinical trials: is blinding necessary? Control Clin Trials. 1996;17:1–12.

[16] Higgins JP, Savović J, Page MJ, Elbers RG, Sterne JA. Assessing risk of bias in a randomized trial. Cochrane handbook for systematic reviews of interventions. 2019:205–28.

[17] Sterne JAC, Savović J, Page MJ, Elbers RG, Blencowe NS, Boutron I, et al. RoB 2: a revised tool for assessing risk of bias in randomised trials. Bmj. 2019;366:l4898.

[18] Sterne JA, Sutton AJ, Ioannidis JP, Terrin N, Jones DR, Lau J, et al. Recommendations for examining and interpreting funnel plot asymmetry in meta-analyses of randomised controlled trials. Bmj. 2011;343:d4002.

[19] Egger M, Davey Smith G, Schneider M, Minder C. Bias in meta-analysis detected by a simple, graphical test. Bmj. 1997;315:629–34.

[20] Duval S, Tweedie R. Trim and fill: A simple funnel-plot-based method of testing and adjusting for publication bias in meta-analysis. Biometrics. 2000;56:455–63.

[21] Borenstein M, Hedges LV, Higgins JP, Rothstein HR. Introduction to meta-analysis: John Wiley & Sons; 2021.

[22] Balduzzi S, Rücker G, Nikolakopoulou A, Papakonstantinou T, Salanti G, Efthimiou O, et al. netmeta: An R package for network meta-analysis using frequentist methods. Journal of Statistical Software. 2023;106:1–40.

[23] Harrer M, Cuijpers P, Furukawa T, Ebert D. Doing meta-analysis with R: A hands-on guide: Chapman and Hall/CRC; 2021.

[24] Rücker G, Schwarzer G. Ranking treatments in frequentist network meta-analysis works without resampling methods. BMC Med Res Methodol. 2015;15:58.

[25] van Valkenhoef G, Lu G, de Brock B, Hillege H, Ades AE, Welton NJ. Automating network meta-analysis. Res Synth Methods. 2012;3:285–99.

[26] Plummer M. JAGS: A program for analysis of Bayesian graphical models using Gibbs sampling. Proceedings of the 3rd international workshop on distributed statistical computing: Vienna, Austria; 2003. p. 1–10.

[27] Olkin I, Dahabreh IJ, Trikalinos TA. GOSH - a graphical display of study heterogeneity. Res Synth Methods. 2012;3:214–23.

[28] Hartigan JA, Wong MA. Algorithm AS 136: A k-means clustering algorithm. Journal of the royal statistical society series c (applied statistics). 1979;28:100–8.

[29] Schubert E, Sander J, Ester M, Kriegel HP, Xu X. DBSCAN revisited, revisited: why and how you should (still) use DBSCAN. ACM Transactions on Database Systems (TODS). 2017;42:1–21.

[30] Fraley C, Raftery AE. Model-based clustering, discriminant analysis, and density estimation. Journal of the American statistical Association. 2002;97:611–31.

[31] Cohen J. Weighted kappa: nominal scale agreement with provision for scaled disagreement or partial credit. Psychol Bull. 1968;70:213–20.

[32] Daly CH, Neupane B, Beyene J, Thabane L, Straus SE, Hamid JS. Empirical evaluation of SUCRA-based treatment ranks in network meta-analysis: quantifying robustness using Cohen’s kappa. BMJ Open. 2019;9:e024625.

[33] Foody GM. Explaining the unsuitability of the kappa coefficient in the assessment and comparison of the accuracy of thematic maps obtained by image classification. Remote sensing of environment. 2020;239:111630.

[34] Revelle W. An introduction to the psych package: Part I: data entry and data description. Northwestern University. 2019.

[35] Salanti G. Indirect and mixed-treatment comparison, network, or multiple-treatments meta-analysis: many names, many benefits, many concerns for the next generation evidence synthesis tool. Research synthesis methods. 2012;3:80–97.

[36] Cipriani G, Ulivi M, Danti S, Lucetti C, Nuti A. Sexual disinhibition and dementia. Psychogeriatrics. 2016;16:145–53.

[37] Cipriani A, Barbui C, Salanti G, Rendell J, Brown R, Stockton S, et al. Comparative efficacy and acceptability of antimanic drugs in acute mania: a multiple-treatments meta-analysis. Lancet. 2011;378:1306–15.

[38] Wu Y-C, Shih M-C, Tu Y-K. Using normalized entropy to measure uncertainty of rankings for network meta-analyses. Medical Decision Making. 2021;41:706–13.

[39] Kraemer HC, Kupfer DJ. Size of treatment effects and their importance to clinical research and practice. Biol Psychiatry. 2006;59:990–6.

[40] Salloway S, Sperling R, Fox NC, Blennow K, Klunk W, Raskind M, et al. Two phase 3 trials of bapineuzumab in mild-to-moderate Alzheimer’s disease. N Engl J Med. 2014;370:322–33.

[41] Vandenberghe R, Rinne JO, Boada M, Katayama S, Scheltens P, Vellas B, et al. Bapineuzumab for mild to moderate Alzheimer’s disease in two global, randomized, phase 3 trials. Alzheimers Res Ther. 2016;8:18.

[42] Doody RS, Thomas RG, Farlow M, Iwatsubo T, Vellas B, Joffe S, et al. Phase 3 trials of solanezumab for mild-to-moderate Alzheimer’s disease. N Engl J Med. 2014;370:311–21.

[43] Honig LS, Vellas B, Woodward M, Boada M, Bullock R, Borrie M, et al. Trial of Solanezumab for Mild Dementia Due to Alzheimer’s Disease. N Engl J Med. 2018;378:321–30.

[44] Budd Haeberlein S, Aisen PS, Barkhof F, Chalkias S, Chen T, Cohen S, et al. Two Randomized Phase 3 Studies of Aducanumab in Early Alzheimer’s Disease. J Prev Alzheimers Dis. 2022;9:197–210.

[45] van Dyck CH, Swanson CJ, Aisen P, Bateman RJ, Chen C, Gee M, et al. Lecanemab in Early Alzheimer’s Disease. N Engl J Med. 2023;388:9–21.

[46] Jeremic D, Navarro-López JD, Jiménez-Díaz L. Efficacy and Safety of Anti-Amyloid-β Monoclonal Antibodies in Current Alzheimer’s Disease Phase III Clinical Trials: A Systematic Review and Interactive Web App-based Meta-Analysis. Ageing Research Reviews. 2023:102012.

[47] Largent EA, Peterson A, Karlawish J, Lynch HF. Aspiring to Reasonableness in Accelerated Approval: Anticipating and Avoiding the Next Aducanumab. Drugs Aging. 2022;39:389–400.

[48] Beshir SA, Aadithsoorya AM, Parveen A, Goh SSL, Hussain N, Menon VB. Aducanumab Therapy to Treat Alzheimer’s Disease: A Narrative Review. Int J Alzheimers Dis. 2022;2022:9343514.

[49] Kuller LH, Lopez OL. ENGAGE and EMERGE: Truth and consequences? Alzheimers Dement. 2021;17:692–5.

[50] Tampi RR, Forester BP, Agronin M. Aducanumab: evidence from clinical trial data and controversies. Drugs Context. 2021;10.

[51] Shcherbinin S, Evans CD, Lu M, Andersen SW, Pontecorvo MJ, Willis BA, et al. Association of Amyloid Reduction After Donanemab Treatment With Tau Pathology and Clinical Outcomes: The TRAILBLAZER-ALZ Randomized Clinical Trial. JAMA Neurol. 2022;79:1015–24.

[52] Ross EL, Weinberg MS, Arnold SE. Cost-effectiveness of Aducanumab and Donanemab for Early Alzheimer Disease in the US. JAMA Neurol. 2022;79:478–87.

[53] Toups K, Hathaway A, Gordon D, Chung H, Raji C, Boyd A, et al. Precision Medicine Approach to Alzheimer’s Disease: Successful Pilot Project. J Alzheimers Dis. 2022;88:1411–21.

[54] Cogswell PM, Barakos JA, Barkhof F, Benzinger TS, Jack CR, Jr., Poussaint TY, et al. Amyloid-Related Imaging Abnormalities with Emerging Alzheimer Disease Therapeutics: Detection and Reporting Recommendations for Clinical Practice. AJNR Am J Neuroradiol. 2022;43:E19–e35.

[55] Doran SJ, Sawyer RP. Risk factors in developing amyloid related imaging abnormalities (ARIA) and clinical implications. Front Neurosci. 2024;18:1326784.

[56] Higgins JP. Cochrane handbook for systematic reviews of interventions version 6.2. Cochrane Database Syst Rev. 2021.

