## Supplementary material for "Donanemab outperformed Aducanumab and Lecanemab on cognitive, but not on biomarker and safety outcomes: systematic review, frequentist and Bayesian network meta-analyses": 01.PRISMA Flow Diagram.pdf

**PRISMA 2020 flow diagram for updated systematic reviews which included searches of databases and registers only**

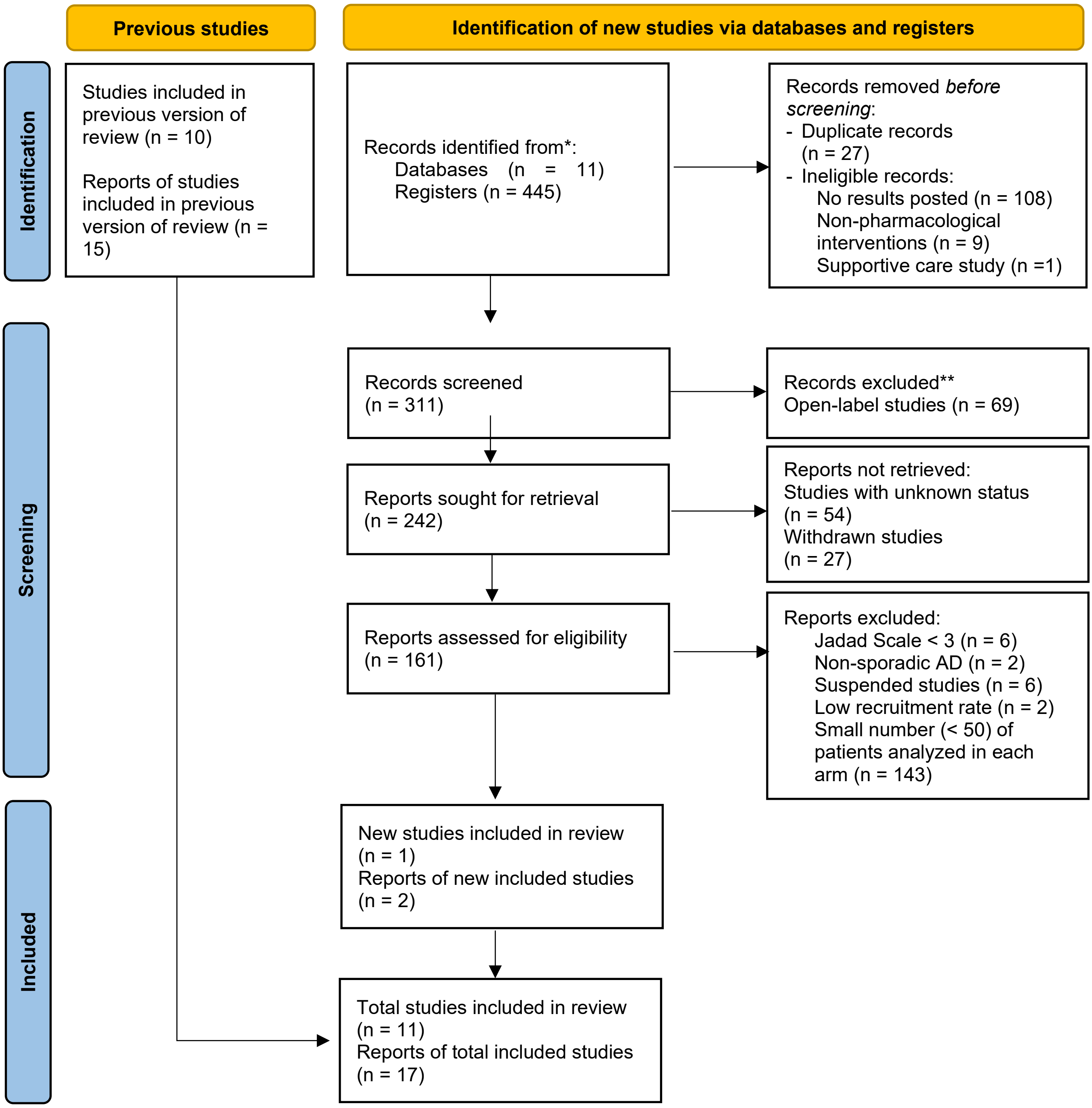


*Consider, if feasible to do so, reporting the number of records identified from each database or register searched (rather than the total number across all databases/registers).

**If automation tools were used, indicate how many records were excluded by a human and how many were excluded by automation tools.

*From:* Page MJ, McKenzie JE, Bossuyt PM, Boutron I, Hoffmann TC, Mulrow CD, et al. The PRISMA 2020 statement: an updated guideline for reporting systematic reviews. BMJ 2021;372:n71. doi: 10.1136/bmj.n71

For more information, visit: http://www.prisma-statement.org/
