## Supplementary material for "Donanemab outperformed Aducanumab and Lecanemab on cognitive, but not on biomarker and safety outcomes: systematic review, frequentist and Bayesian network meta-analyses": 02.Supplementary Methods and Results (Tables S1-S8).pdf

**1. METHODS ... 3**

1.1. SUCRA Score in Bayesian NMA ... 3

1.2. P-Score in Frequentist NMA ... 3

1.3. Rank Uncertainty ... 3

**2. RESULTS ... 5**

**TABLE S1. PRIMARY OUTCOMES: Efficacy on cognitive outcomes ... 5**

The Mini Mental Examination (MMSE) ... 5

**TABLE S2. SECONDARY OUTCOMES: Biomarkers of Amyloid and *Tau* ... 6**

TABLE S2.1. Amyloid Burden on PET (centiloid scale) ... 6

TABLE S2.2. Cerebrospinal fluid (CSF) A$\beta_{1-42}$ ... 7

TABLE S2.3. CSF p-*tau* ... 7

**TABLE S3. SECONDARY OUTCOMES: Safety Measures ... 8**

TABLE S3.1. Serious adverse events ... 8

TABLE S3.2. Total events of ARIA-H ... 9

TABLE S3.3. ARIA-H: Cerebral Microhemorrhages ... 9

TABLE S3.4. Headaches ... 10

TABLE S3.5. Falls ... 10

TABLE S3.6. Nausea ... 11

TABLE S3.7. Fatigue ... 11

TABLE S3.8. Dizziness ... 12

TABLE S3.9. Syncope ... 12

TABLE S3.10. Arthralgia ... 13

TABLE S3.11. Back Pain ... 13

TABLE S3.12. Diarrhea ... 14

TABLE S3.13. Cardiac disorders ... 14

TABLE S4.14. Nasopharyngitis ... 15

TABLE S3.15. Upper respiratory infections ... 15

TABLE S3.16. Urinary infections ... 16

**TABLE S4. TERTIARY OUTCOMES ... 17**

TABLE S4.1. Alzheimer's Disease Cooperative Study - Activities of Daily Living Scale for use in Mild Cognitive Impairment (ADCS-ADL-MCI) ... 17

TABLE S4.2. ARIA-H: Superficial Siderosis ... 18

TABLE S4.3. Infusion-related reactions ... 18

TABLE S4.4. Total events of ARIA-E (cerebral edema) in APOE-ε4 carriers ... 19

TABLE S4.5. Total events of ARIA-E (cerebral edema) in APOE-ε4 non-carriers ... 20

TABLE S4.6. Total events of ARIA-H in APOE-ε4 carriers ... 21

### 1. METHODS

Ranking of treatments is proposed to facilitate the interpretation of comparative effectiveness and to support clinical decision making, which is often determined by the mean rank, SUCRA, P-score and other scores. Here we briefly describe recently developed measures of rankings used in this study as well as the measures of their uncertainties.

#### 1.1. SUCRA Score in Bayesian NMA

In a Bayesian framework and for every Markov Chain Monte Carlo (MCMC) cycle, each treatment is ranked based on the estimated treatment effects. Introduced by Salanti et al (Salanti, 2012), SUCRA score is calculated based on the Bayesian posterior distributions:

$$\text{SUCRA}_J = \frac{\sum_{b=1}^{a-1} cum.prob_{jb}}{a-1},$$

where $j$ is some treatment, $a$ are all competing treatments, $b$ are the $b$ = 1, 2, …, $a$ −1 best treatments, and *cum.prob* represents the cumulative probability of a treatment being among the $b$ best treatments. $\text{SUCRA}_J$ can be interpreted as the average proportion of treatments worse than $j$ and can range from 0 to 100%. The closer SUCRA is to 100%, the higher the likelihood that treatment $j$ is the best. To calculate the SUCRA scores in R, we used the *sucra* function from *dmetar* package in R (Harrer *et al.*, 2021).

#### 1.2. P-Score in Frequentist NMA

Rücker and Schwarzer (2015) proposed ranking treatments without resampling methods in frequentist NMA by using P-scores, analogue to SUCRA scores. P-scores can be interpreted as the mean extent of certainty that one treatment is better than another treatment, averaged over all competing treatments. P-score of 1 is the best possible score and 0 is the worst. The authors pointed out that neither SUCRA nor P-score offer a major advantage compared to looking at credible or confidence intervals. In this study, P-score was calculated by using *netrank* function available in *netmeta* package in R (Rücker & Schwarzer, 2015).

#### 1.3. Rank Uncertainty

To make an informed clinical decision, estimates of treatment effects derived from the results of NMAs should be interpreted with due consideration of their uncertainty, since vulgar interpretation based on simple scores can be misleading (Beis & Papasotiriou, 2023). However, the interpretation of these measures is not always straightforward, and it is not clear whether reporting uncertainty is useful in all cases (Veroniki *et al.*, 2018). The uncertainty of the ranking scores is becoming increasingly discussed in literature and currently there is no consensus on the utility of these scores and the ways to interpret and compare their uncertainties between NMAs. The Preferred Reporting Items for Systematic Reviews and Meta-Analyses (PRISMA)–NMA guideline and Grading of Recommendation, Assessment, Development, and Evaluation (GRADE) working group suggested that the uncertainty of ranking for each treatment should be reported, either by rankograms plots that show the distribution of the ranking probabilities, the credible/confidence intervals of the mean rank, or the interquartile range (IQR) of median rank. Salanti et al. (2012) introduced a rank statistic and extended the consideration to the probabilities that a treatment out of $n$ treatments in a NMA is the best, the second, the third and so on until the least effective treatment (Salanti, 2012). They also introduced several graphical presentations of ranking, such as rankograms, bar graphs and scatterplots (Cipriani *et al.*, 2011; Cipriani *et al.*, 2016).

In this work, the uncertainty of rankings is visualized by rankograms and tables showing the distribution of the ranking probabilities (Cipriani *et al.*, 2011; Salanti, 2012; Cipriani *et al.*, 2016). We also measured Shannon's information entropy, as suggested recently by Wu et al (2021) (Wu *et al.*, 2021). The authors proposed very intuitive way to estimate uncertainty associated with ranking probabilities in NMA by applying Shannon's information entropy formula to obtain a normalised entropy score. They showed that the normalised entropy score gives a more accurate assessment of ranking uncertainty and does not depend on the number of analysed treatments. This approach can be used to compare the uncertainty of treatment rankings within a NMA and also between different NMAs, which is crucial for interpretation of results (Wu *et al.*, 2021).

Initially introduced by Claude Shannon in 1948 (Shannon, 1948), the concept of information entropy is directly analogous to the entropy in statistical thermodynamics. The Shannon entropy H(x) for a discrete random variable x with a probability mass function P ($x_i$) is calculated in the following way:

$$\mathrm{H(x)} = -\sum_{i=1}^{n} P(x=i) log_b P(x=i),$$

where $n$ is the number of possible outcomes of the random variable X, P ($x = i$) is the probability of the $i$-th outcome, $b$ is the base of logarithm, and the unit of entropy depends on the choice of $b$. With the base-2 logarithms, the entropy is measured in bit (binary digit), which is the smallest unit of data in a computer. Normalized entropy thus rescales the entropy to a range from 0 to 1 ($\mathrm{H(x)} \in [0,1]$). For a NMA, the most precise scenario would be that we are absolutely certain in the ranking of treatments in our network. Therefore, each treatment would have 100% probability of being in one ranking position and 0% probability for the other positions (peaked distribution). Under this scenario, the entropy is zero bit, and normalized entropy equals zero. On the other hand, the normalized entropy reaches 1 in the least precise scenario when the ranking probabilities are the same for each rank (flat distribution). In this work, we regarded as "low" those values of normalized entropy that were equal or lower than 0.25 and "moderate" those between 0.25 and 0.5. Larger values were considered as conveying "high" or "substantial" uncertainty or normalized entropy associated with ranking probabilities.

Below is the entropy_arm_function for calculating normalized entropy in R that is used in this study (originally published by (Wu *et al.*, 2021). The inputs for this function are the number of treatments (k) and ranking probabilities matrix (a), with rows representing the ranks (1st, 2nd, 3rd…) and columns representing interventions / drugs in the network.

```
entropy_arm_function <- function(a, k) {
  e = -a * log(a, 2) #
  e[!is.finite(e)] <- 0
  e_colsum = colSums(e)
  e_max = k * (-1/k) * log2(1/k)
  e_prop = e_colsum/e_max
  return(e_prop)
}
```

#### 2. RESULTS

##### TABLE S1. PRIMARY OUTCOMES: Efficacy on cognitive outcomes

League tables representing the results of network meta-analysis (NMA) (random-effect model). The values in the tables below represent relative effects (Standardized Mean Difference) for primary (efficacy) outcomes. Bolded numbers indicate statistical significance ($p < 0.05$) for Frequentist NMA, or strong evidence in Bayesian NMA. The numbers in parentheses represent confidence intervals (CIs) for Frequentist NMA, or credible intervals (CrIs) in Bayesian framework. Dagger symbol (†) denotes when the interpretation differs significantly between frequentist and Bayesian NMA results, indicating the uncertainty and lack of strong evidence for a non-zero effect.

##### The Mini Mental Examination (MMSE)

For each comparison, the number above 0 suggests that the drug in the column is superior to the drug in the row, and darker green colors correspond to greater relative effect.

###### Frequentist Network Meta-Analysis Results

| | | | | |
|---|---|---|---|---|
| Aducanumab | | | | |
| 0.028 (-0.0826; 0.1386) | Bapineuzumab | | | |
| -0.0972 (-0.2326; 0.0383) | -0.1251 (-0.2597; 0.0094) | Donanemab | | |
| 0.0442 (-0.0348; 0.1233) | 0.0162 (-0.0611; 0.0936) | **0.1414 * (0.0314; 0.2514)** | Placebo | |
| -0.0434 (-0.1447; 0.0580) | -0.0714 (-0.1714; 0.0287) | 0.0538 (-0.0732; 0.1808) | **-0.0876 ** (-0.1511; -0.0242)** | Solanezumab |

Statistical significance for Drug vs. Placebo: * p = 0.0118; ** p = 0.0068

$I^2$ = 0% [0.0%; 64.8%]; p-value of Q statistics = 0.6881 in heterogeneity (within designs).

###### Bayesian Network Meta-Analysis Results

| | | | | |
|---|---|---|---|---|
| Aducanumab | | | | |
| 0.0272 (-0.1049; 0.159) | Bapineuzumab | | | |
| -0.0936 (-0.2516; 0.0648) | -0.1212 (-0.2783, 0.0401) | Donanemab | | |
| 0.0449 (-0.047, 0.1363) | 0.0174 (-0.0748, 0.1118) | **0.1391 (0.0091, 0.2685)** | Placebo | |
| -0.0408 (-0.1636; 0.0820) | -0.0683 (-0.1922, 0.0579) | 0.0524 (-0.1007, 0.2069) | **-0.0861 (-0.1676, -0.0028)** | Solanezumab |

#### TABLE S2. SECONDARY OUTCOMES: Biomarkers of Amyloid and *Tau*

League tables representing the results of network meta-analysis (NMA) (random-effect model). The values in the tables below represent relative effects (Standardized Mean Difference) for efficacy (biomarker) outcomes. Bolded numbers indicate statistical significance ($p < 0.05$) for Frequentist NMA, or strong evidence in Bayesian NMA. The numbers in parentheses represent confidence intervals (CIs) for Frequentist NMA, or credible intervals (CrIs) in Bayesian framework. Dagger symbol (†) denotes when the interpretation differs significantly between frequentist and Bayesian NMA results, indicating the uncertainty and lack of strong evidence for a non-zero effect.

##### TABLE S2.1. Amyloid Burden on PET (centiloid scale)

For each comparison, the number below 0 suggests that the drug in the column is superior to the drug in the row, and darker green colors correspond to greater relative amyloid removal on PET.

###### Frequentist Network Meta-Analysis Results

| | | | |
|---|---|---|---|
| Aducanumab | | | |
| -0.3105 (-1.6022; 0.9812) | Donanemab | | |
| **3.2694 † (1.5316; 4.9872)** | **3.5699 † (1.7036; 5.4363)** | Lecanemab | |
| **-4.5403 * (-5.3053; -3.7752)** | **-4.2298 * (-5.2705; -3.1890)** | **-7.7997 * (-9.3489; -6.2505)** | Placebo |

Statistical significance: Drug vs. Placebo: * $p < 0.0001$
Heterogeneity $I^2 = 92.5\%$ [85.4%; 96.1%]; p-value of Q statistics < 0.0001 in heterogeneity (within designs). Design-specific decomposition of within-designs: Placebo vs Aducanumab $p < 0.0001$ and Placebo vs Donanemab $p = 0.0005$.

###### Bayesian Network Meta-Analysis Results

| | | | |
|---|---|---|---|
| Aducanumab | | | |
| -0.3233 (-2.9757; 2.2818) | Donanemab | | |
| 3.2397 † (-0.2632; 6.6676) | 3.5576 † (-0.1665; 7.3472) | Lecanemab | |
| **-4.5484 (-6.1205; -3.007)** | **-4.2300 (-6.3827, -2.036)** | **-7.7946 (-10.8581, -4.6608)** | Placebo |

#### TABLE S2.2. Cerebrospinal fluid (CSF) Aβ1-42

For each comparison, the number above 0 suggests that the drug in the column is superior to the drug in the row, and darker green colors correspond to more beneficial relative effect.

##### Frequentist Network Meta-Analysis Results

| | | | |
|---|---|---|---|
| Aducanumab | | | |
| 0.2363 (-0.9143; 1.3869) | Lecanemab | | |
| **1.3419 * (0.7555; 1.9283)** | **1.1056 † ** (0.1157; 2.0956)** | Placebo | |
| **1.3161 † (0.3541; 2.2780)** | 1.0798 (-0.1699; 2.3294) | -0.0258 (-0.7885; 0.7368) | Solanezumab |

Statistical significance for Drug vs. Placebo: * p < 0.0001; ** p = 0.0286
$I^2$ = 68.9% [20.0%; 87.9%]; p-value of Q statistics = 0.012 in heterogeneity (within designs). Design-specific decomposition of within-designs: Placebo vs Aducanumab p = 0.0056 and Placebo vs Solanezumab p = 0.6049.

##### Bayesian Network Meta-Analysis Results

| | | | |
|---|---|---|---|
| Aducanumab | | | |
| 0.23274 (-1.9202; 2.4216) | Lecanemab | | |
| **1.3402 (0.3633; 2.348)** | 1.1054 † (-0.8064; 3.026) | Placebo | |
| 1.3167 † (-0.360; 3.0239) | 1.0804 (-1.3070; 3.4616) | -0.0253 (-1.4156; 1.3743) | Solanezumab |

#### TABLE S2.3. CSF p-*tau*

For each comparison, the number below 0 suggests that the drug in the column is superior to the drug in the row, and darker green colors correspond to greater relative effect at reducing p-*tau*.

##### Frequentist Network Meta-Analysis Results

| | | | |
|---|---|---|---|
| Aducanumab | | | |
| **-0.472 † (-0.8500; -0.0941)** | Bapineuzumab | | |
| -0.1723 (-0.6199; 0.2754) | 0.2998 (-0.0748; 0.6743) | Lecanemab | |
| **-0.664 * (-0.9826; -0.3455)** | -0.192 ** (-0.3954; 0.0114) | **-0.4918 † *** (-0.8063;-0.1773)** | Placebo |

Statistical significance for Drug vs. Placebo: * p < 0.0001; ** p = 0.0643; *** p = 0.0022
$I^2$ = 9.2% [0.0%; 77.0%]; p-value of Q statistics = 0.3571 in heterogeneity (within designs).

##### Bayesian Network Meta-Analysis Results

| | | | |
|---|---|---|---|
| Aducanumab | | | |
| -0.46794 † (-1.013; 0.0839) | Bapineuzumab | | |
| -0.1643 (-0.9052; 0.59203) | 0.3010 (-0.4289; 1.0305) | Lecanemab | |
| **-0.65803 (-1.0599, -0.2308)** | -0.1917 (-0.5619; 0.1825) | -0.4938 † (-1.1218; 0.13487) | Placebo |

#### TABLE S3. SECONDARY OUTCOMES: Safety Measures

League tables representing the results of network meta-analysis (NMA) (random-effect model). The values in the tables below represent Relative Risks (natural logarithm) for safety outcomes. Bolded numbers indicate statistical significance ($p < 0.05$) for Frequentist NMA, or strong evidence in Bayesian NMA. The numbers in parentheses represent confidence intervals (CIs) for Frequentist NMA, or credible intervals (CrIs) in Bayesian framework. Dagger symbol (†) denotes when the interpretation differs significantly between frequentist and Bayesian NMA results, indicating the uncertainty and lack of strong evidence for a non-zero effect.

For each comparison, the number below 0 suggests that the drug in the column is safer than the drug in the row, and darker red colors correspond to relatively higher risk.

##### TABLE S3.1. Serious adverse events

###### Frequentist Network Meta-Analysis Results

| | | | | | |
|---|---|---|---|---|---|
| Aducanumab | | | | | |
| -0.0146 (-0.2709; 0.2418) | Bapineuzumab | | | | |
| -0.1162 (-0.3936; 0.1612) | -0.1017 (-0.4056; 0.2023) | Donanemab | | | |
| -0.2375 (-0.5494; 0.0745) | -0.2229 (-0.5586; 0.1128) | -0.1213 (-0.4734; 0.2308) | Lecanemab | | |
| -0.0175 (-0.1759; 0.1411) | -0.0028 (-0.2043; 0.1986) | 0.0988 (-0.1289; 0.3264) | 0.22 (-0.0486; 0.4886) | Placebo | |
| 0.1138 (-0.1527; 0.3803) | 0.1284 (-0.1657; 0.4225) | 0.23 (-0.0826; 0.5426) | **0.3513 † (0.0077; 0.6948)** | 0.1312 (-0.0831; 0.3454) | Solanezumab |

Heterogeneity $I^2$ = 10.9% [0.0%; 74.0%]; p-value of Q statistics = 0.3464 in heterogeneity (within designs).

###### Bayesian Network Meta-Analysis Results

| | | | | | |
|---|---|---|---|---|---|
| Aducanumab | | | | | |
| -0.0024 (-0.2980, 0.3281) | Bapineuzumab | | | | |
| -0.1227 (-0.4737, 0.2119) | -0.1211 (-0.5195, 0.2319) | Donanemab | | | |
| -0.2361 (-0.6409, 0.1659) | -0.2337 (-0.6800, 0.1773) | -0.1109 (-0.5570, 0.3416) | Lecanemab | | |
| -0.0173 (-0.2112, 0.1799) | -0.0147 (-0.2768, 0.2157) | 0.1060 (-0.1674, 0.3948) | 0.2189 (-0.1340, 0.5713) | Placebo | |
| 0.1147 (-0.2518, 0.4876) | 0.1194 (-0.3064, 0.4973) | 0.2388 (-0.1753, 0.6703) | 0.3507 † (-0.1189, 0.8293) | 0.1322 (-0.1858, 0.4459) | Solanezumab |

#### TABLE S3.2. Total events of ARIA-H

Including all incidents of cerebral microhemorrhages, macrohemorrhages and superficial siderosis.

##### Frequentist Network Meta-Analysis Results

| | | | | |
|---|---|---|---|---|
| Aducanumab | | | | |
| **0.3764 † (0.1051; 0.6477)** | Donanemab | | | |
| **0.5679 † (0.2412; 0.8947)** | 0.1915 (-0.1644; 0.5475) | Lecanemab | | |
| **1.2158 * (1.0521; 1.3795)** | **0.8394 * (0.6231; 1.0557)** | **0.6478 * (0.3651; 0.9306)** | Placebo | |
| **1.3488 (0.9247; 1.7728)** | **0.9724 (0.5254; 1.4194)** | **0.7808 (0.2982; 1.2635)** | 0.133 (-0.2583; 0.5242) | Solanezumab |

Statistical significance for Drug vs. Placebo: * $p < 0.0001$

Heterogeneity $I^2$ = 15.3% [0.0%; 82.4%]; p-value of Q statistics = 0.3168 in heterogeneity (within designs).

##### Bayesian Network Meta-Analysis Results

| | | | | |
|---|---|---|---|---|
| Aducanumab | | | | |
| 0.3712 † (-0.1116, 0.8364) | Donanemab | | | |
| 0.5672 † (-0.0344, 1.1517) | 0.1959 (-0.4633, 0.8530) | Lecanemab | | |
| **1.2149 (0.9373, 1.4898)** | **0.8444 (0.4627, 1.2382)** | **0.6477 (0.1300, 1.1860)** | Placebo | |
| **1.3479 (0.6965, 2.0116)** | **0.9772 (0.2844, 1.6912)** | **0.7796 (0.0032, 1.5773)** | 0.1319 (-0.4560, 0.7280) | Solanezumab |

#### TABLE S3.3. ARIA-H: Cerebral Microhemorrhages

##### Frequentist Network Meta-Analysis Results

| | | | | | |
|---|---|---|---|---|---|
| Aducanumab | | | | | |
| -0.2422 (-0.5692; 0.0846) | Bapineuzumab | | | | |
| -0.2422 (-0.0503; 0.5036) | **0.4688 † (0.1273; 0.8103)** | Donanemab | | | |
| **0.3763 † (0.0418; 0.7108)** | **0.6185 (0.229; 1.0081)** | 0.1497 (-0.1991; 0.4985) | Lecanemab | | |
| **0.992 * (0.809; 1.1749)** | **1.2342 * (0.9633; 1.5051)** | **0.7654 * (0.5574; 0.9733)** | **0.6157 * (0.3356; 0.8957)** | Placebo | |
| **0.979 (0.6341; 1.3238)** | **1.2212 (0.8226; 1.6197)** | **0.7524 (0.3936; 1.1111)** | **0.6026 (0.1978; 1.0074)** | -0.013 (-0.3053; 0.2793) | Solanezumab |

Statistical significance for Drug vs. Placebo: * $p < 0.0001$

Heterogeneity $I^2$ = 15.3% [0.0%; 82.4%]; p-value of Q statistics = 0.3168 in heterogeneity (within designs).

##### Bayesian Network Meta-Analysis Results

| | | | | | |
|---|---|---|---|---|---|
| Aducanumab | | | | | |
| -0.2278 (-0.6516, 0.2280) | Bapineuzumab | | | | |
| 0.2144 (-0.2034, 0.6034) | 0.4420 † (-0.0722, 0.8926) | Donanemab | | | |
| 0.3779 † (-0.1196, 0.8711) | **0.6047 (0.0267, 1.1440)** | 0.1633 (-0.3621, 0.7170) | Lecanemab | | |
| **0.9932 (0.7467, 1.2376)** | **1.2209 (0.8455, 1.5609)** | **0.7784 (0.4729, 1.1125)** | **0.6148 (0.1840, 1.0559)** | Placebo | |
| **0.9731 (0.5172, 1.4113)** | **1.1976 (0.6579, 1.7012)** | **0.7567 (0.2728, 1.2566)** | **0.5945 (0.0172, 1.1599)** | -0.0205 (-0.4013, 0.3493) | Solanezumab |

#### TABLE S3.4. Headaches

##### Frequentist Network Meta-Analysis Results

| | | | | | |
|---|---|---|---|---|---|
| Aducanumab | | | | | |
| **0.4081 † (0.0629; 0.7532)** | Bapineuzumab | | | | |
| -0.1037 (-0.5775; 0.3701) | **-0.5118 † (-0.98; -0.0436)** | Donanemab | | | |
| -0.059 (-0.6296; 0.5116) | -0.4671 (-1.0328; 0.0988) | 0.0447 (-0.6077; 0.6971) | Lecanemab | | |
| **0.2546 † * (0.0051; 0.5041)** | -0.1535 (-0.3919; 0.085) | 0.3583 ** (-0.0446; 0.7612) | 0.3136 (-0.1995; 0.8268) | Placebo | |
| 0.2877 (-0.1557; 0.7312) | -0.1204 (-0.5577; 0.317) | 0.3914 (-0.1533; 0.9362) | 0.3467 (-0.284; 0.9774) | 0.0331 (-0.3335; 0.3998) | Solanezumab |

Statistical significance for Drug vs. Placebo: * p = 0.0455; ** p = 0.0813

Heterogeneity $I^2$ = 63.2% [29.4%; 80.8%]; p-value of Q statistics = 0.0025 in heterogeneity (within designs). Design-specific decomposition of within-designs: Placebo vs Bapineuzumab p < 0.0001, and non-significant (p > 0.05) for other comparisons.

##### Bayesian Network Meta-Analysis Results

| | | | | | |
|---|---|---|---|---|---|
| Aducanumab | | | | | |
| 0.4091 † (-0.0204; 0.8158) | Bapineuzumab | | | | |
| -0.1008 (-0.6670; 0.4568) | -0.5099 † (-1.0587; 0.0466) | Donanemab | | | |
| -0.0610 (-0.7642; 0.6251) | -0.4705 (-1.1424; 0.2265) | 0.0431 (-0.7405; 0.8196) | Lecanemab | | |
| 0.2538 † (-0.0523; 0.5573) | -0.1540 (-0.4215; 0.1378) | 0.3562 (-0.1147; 0.8334) | 0.3154 (-0.3048; 0.9352) | Placebo | |
| 0.2878 (-0.2559; 0.8246) | -0.1221 (-0.6337; 0.4236) | 0.3892 (-0.2523; 1.0395) | 0.3483 (-0.4049; 1.1166) | 0.0341 (-0.4141; 0.4761) | Solanezumab |

#### TABLE S3.5. Falls

##### Frequentist Network Meta-Analysis Results

| | | | | | |
|---|---|---|---|---|---|
| Aducanumab | | | | | |
| 0.2148 (-0.0052; 0.4348) | Bapineuzumab | | | | |
| 0.1192 (-0.1703; 0.4087) | -0.0956 (-0.3854; 0.1942) | Donanemab | | | |
| 0.1014 (-0.2173; 0.42) | -0.1134 (-0.4323; 0.2055) | -0.0178 (-0.388; 0.3525) | Lecanemab | | |
| **0.1786 † * (0.0232; 0.3338)** | -0.0362 (-0.1921; 0.1196) | 0.0594 (-0.1849; 0.3037) | 0.0771 (-0.2011; 0.3554) | Placebo | |
| 0.2227 (-0.0003; 0.4456) | 0.0079 (-0.2155; 0.2313) | 0.1035 (-0.1886; 0.3955) | 0.1212 (-0.1997; 0.4422) | 0.0441 (-0.1159; 0.2042) | Solanezumab |

Statistical significance for Drug vs. Placebo: * p = 0.0242

Heterogeneity $I^2$ = 0% [0.0%; 60.2%]; p-value of Q statistics = 0.7272 in heterogeneity (within designs).

##### Bayesian Network Meta-Analysis Results

| | | | | | |
|---|---|---|---|---|---|
| Aducanumab | | | | | |
| 0.2180 (-0.0357, 0.4745) | Bapineuzumab | | | | |
| 0.1225 (-0.2141, 0.4650) | -0.0947 (-0.4285, 0.2444) | Donanemab | | | |
| 0.1017 (-0.2885, 0.4990) | -0.1155 (-0.5014, 0.2712) | -0.0217 (-0.4700, 0.4286) | Lecanemab | | |
| 0.1804 † (-0.0043, 0.3667) | -0.0378 (-0.2182, 0.1397) | 0.0570 (-0.2321, 0.3406) | 0.0780 (-0.2702, 0.4230) | Placebo | |
| 0.2279 (-0.0469, 0.5170) | 0.0114 (-0.2666, 0.2962) | 0.1070 (-0.2544, 0.4662) | 0.1254 (-0.2787, 0.5386) | 0.0486 (-0.1590, 0.2711) | Solanezumab |

#### TABLE S3.6. Nausea

##### Frequentist Network Meta-Analysis Results

| | | | | |
|---|---|---|---|---|
| Aducanumab | | | | |
| -0.2062 (-0.7302; 0.3178) | Bapineuzumab | | | |
| **-1.018 † (-1.9126; -0.1234)** | -0.8119 (-1.7958; 0.1716) | Donanemab | | |
| -0.087 (-0.319; 0.1449) | 0.1191 (-0.3507; 0.589) | **0.9311 † * (0.067; 1.7952)** | Placebo | |
| -0.1652 (-0.5237; 0.1933) | 0.0409 (-0.5027; 0.5846) | 0.8529 (-0.0534; 1.7593) | -0.0782 (-0.3515; 0.1952) | Solanezumab |

Statistical significance for Drug vs. Placebo: * p = 0.0347
Heterogeneity $I^2$ = 0% [0.0%; 79.2%]; p-value of Q statistics = 0.5479 in heterogeneity (within designs).

##### Bayesian Network Meta-Analysis Results

| | | | | |
|---|---|---|---|---|
| Aducanumab | | | | |
| -0.2037 (-0.9856; 0.5817) | Bapineuzumab | | | |
| -1.0143 † (-2.0644; 0.0545) | -0.8076 (-2.0356; 0.4118) | Donanemab | | |
| -0.0835 (-0.4238; 0.2702) | 0.119 (-0.5837; 0.8235) | 0.9287 † (-0.0829; 1.9279) | Placebo | |
| -0.1603 (-0.7216; 0.4168) | 0.0434 (-0.7806; 0.8877) | 0.8507 (-0.2489; 1.9423) | -0.0782 (-0.3515; 0.1952) | Solanezumab |

#### TABLE S3.7. Fatigue

##### Frequentist Network Meta-Analysis Results

| | | | |
|---|---|---|---|
| Aducanumab | | | |
| -0.1603 (-0.6298; 0.3092) | Donanemab | | |
| -0.2064 (-0.4357; 0.0229) | -0.0461 (-0.4559; 0.3635) | Placebo | |
| -0.2709 (-0.6463; 0.1043) | -0.1107 (-0.6167; 0.3953) | -0.0645 (-0.3615; 0.2325) | Solanezumab |

Heterogeneity $I^2$ = 0% [0.0%; 74.6%]; p-value of Q statistics = 0.7870 in heterogeneity (within designs).

##### Bayesian Network Meta-Analysis Results

| | | | |
|---|---|---|---|
| Aducanumab | | | |
| -0.1618 (-0.7005; 0.3689) | Donanemab | | |
| -0.2070 (-0.4773; 0.0656) | -0.0457 (-0.5043; 0.4141) | Placebo | |
| -0.2676 (-0.7166; 0.1895) | -0.1043 (-0.6887; 0.4815) | -0.0610 (-0.4207; 0.2986) | Solanezumab |

#### TABLE S3.8. Dizziness

##### Frequentist Network Meta-Analysis Results

| | | | | | |
|---|---|---|---|---|---|
| Aducanumab | | | | | |
| 0.0635 (-0.3133; 0.4403) | Bapineuzumab | | | | |
| -0.1233 (-0.6129; 0.3662) | -0.1868 (-0.7148; 0.3412) | Donanemab | | | |
| -0.0539 (-0.5722; 0.4642) | -0.1174 (-0.6722; 0.4372) | 0.0693 (-0.5672; 0.706) | Lecanemab | | |
| 0.0081 (-0.2187; 0.2349) | -0.0554 (-0.3564; 0.2455) | 0.1314 (-0.3025; 0.5653) | 0.062 (-0.4038; 0.528) | Placebo | |
| -0.0396 (-0.4204; 0.3413) | -0.103 (-0.5322; 0.3261) | 0.0838 (-0.4471; 0.6147) | 0.0144 (-0.543; 0.5718) | -0.0476 (-0.3535; 0.2583) | Solanezumab |

Heterogeneity $I^2$ = 24.6% [0.0%; 64.5%]; p-value of Q statistics = 0.2251 in heterogeneity (within designs).

##### Bayesian Network Meta-Analysis Results

| | | | | | |
|---|---|---|---|---|---|
| Aducanumab | | | | | |
| 0.0665 (-0.3725; 0.5073) | Bapineuzumab | | | | |
| -0.1335 (-0.7290; 0.4263) | -0.1966 (-0.8256; 0.3939) | Donanemab | | | |
| -0.0554 (-0.6926; 0.5830) | -0.1231 (-0.7896; 0.5477) | 0.0743 (-0.6619; 0.8781) | Lecanemab | | |
| 0.0081 (-0.2713; 0.2845) | -0.0577 (-0.3997; 0.2854) | 0.1390 (-0.3480; 0.6647) | 0.0654 (-0.5072; 0.6341) | Placebo | |
| -0.0364 (-0.5147; 0.4370) | -0.1021 (-0.6251; 0.4081) | 0.0918 (-0.5176; 0.7467) | 0.0179 (-0.6675; 0.6983) | -0.0462 (-0.4340; 0.3345) | Solanezumab |

#### TABLE S3.9. Syncope

##### Frequentist Network Meta-Analysis Results

| | | | |
|---|---|---|---|
| Aducanumab | | | |
| 0.2576 (-0.673; 1.1881) | Bapineuzumab | | |
| 0.3837 (-0.4368; 1.2042) | 0.1261 (-0.3129; 0.565) | Placebo | |
| 0.5483 (-0.4017; 1.4982) | 0.2907 (-0.3588; 0.9401) | 0.1646 (-0.3142; 0.6432) | Solanezumab |

Heterogeneity $I^2$ = 0% [0.0%; 62.4%]; p-value of Q statistics = 0.8886 in heterogeneity (within designs).

##### Bayesian Network Meta-Analysis Results

| | | | |
|---|---|---|---|
| Aducanumab | | | |
| 0.2535 (-0.7977; 1.2961) | Bapineuzumab | | |
| 0.3789 (-0.5082; 1.2656) | 0.1271 (-0.4230; 0.6719) | Placebo | |
| 0.5783 (-0.5287; 1.7015) | 0.3252 (-0.5386; 1.2228) | 0.1986 (-0.4663; 0.9053) | Solanezumab |

#### TABLE S3.10. Arthralgia

##### Frequentist Network Meta-Analysis Results

| | | | | | |
|---|---|---|---|---|---|
| Aducanumab | | | | | |
| -0.3679 (-0.917; 0.1813) | Bapineuzumab | | | | |
| -0.1942 (-0.8347; 0.4464) | 0.1737 (-0.5135; 0.8609) | Donanemab | | | |
| 0.0918 (-0.5897; 0.7733) | 0.4596 (-0.2659; 1.1852) | 0.2859 (-0.511; 1.0829) | Lecanemab | | |
| -0.0661 (-0.4123; 0.28) | 0.3017 (-0.1247; 0.728) | 0.128 (-0.411; 0.667) | -0.1579 (-0.7451; 0.4291) | Placebo | |
| 0.0906 (-0.4785; 0.6597) | 0.4584 (-0.1628; 1.0796) | 0.2847 (-0.4186; 0.988) | -0.0012 (-0.7421; 0.7396) | 0.1567 (-0.2952; 0.6086) | Solanezumab |

Heterogeneity $I^2$ = 37.7% [0.0%; 70.3%]; p-value of Q statistics = 0.1072 in heterogeneity (within designs).

##### Bayesian Network Meta-Analysis Results

| | | | | | |
|---|---|---|---|---|---|
| Aducanumab | | | | | |
| -0.3604 (-1.0137; 0.2829) | Bapineuzumab | | | | |
| -0.2012 (-0.9457; 0.5826) | 0.1565 (-0.6142; 0.9947) | Donanemab | | | |
| 0.0964 (-0.8006; 0.9756) | 0.4545 (-0.4712; 1.3688) | 0.2982 (-0.7635; 1.2745) | Lecanemab | | |
| -0.0632 (-0.4931; 0.3527) | 0.2979 (-0.1957; 0.7757) | 0.1389 (-0.5411; 0.7460) | -0.1563 (-0.9448; 0.6249) | Placebo | |
| 0.0965 (-0.6416; 0.7921) | 0.4535 (-0.3079; 1.1959) | 0.2973 (-0.6170; 1.1096) | 0.0002 (-0.9867; 0.9666) | 0.1562 (-0.4231; 0.7234) | Solanezumab |

#### TABLE S3.11. Back Pain

##### Frequentist Network Meta-Analysis Results

| | | | | |
|---|---|---|---|---|
| Aducanumab | | | | |
| -0.4405 (-1.0856; 0.2047) | Bapineuzumab | | | |
| -0.1851 (-0.6677; 0.2974) | 0.2554 (-0.4739; 0.9848) | Lecanemab | | |
| -0.0431 (-0.2852; 0.1989) | 0.3974 (-0.2006; 0.9955) | 0.142 (-0.2755; 0.5594) | Placebo | |
| -0.1624 (-0.5367; 0.212) | 0.2782 (-0.3846; 0.9409) | 0.0227 (-0.483; 0.5285) | -0.1192 (-0.4048; 0.1663) | Solanezumab |

Heterogeneity $I^2$ = 21.4% [0.0%; 67.0%]; p-value of Q statistics = 0.2781 in heterogeneity (within designs).

##### Bayesian Network Meta-Analysis Results

| | | | | |
|---|---|---|---|---|
| Aducanumab | | | | |
| -0.4465 (-1.2372; 0.3482) | Bapineuzumab | | | |
| -0.1898 (-0.8579; 0.4643) | 0.2569 (-0.6727; 1.1793) | Lecanemab | | |
| -0.0473 (-0.3655; 0.2617) | 0.3986 (-0.3252; 1.1176) | 0.1430 (-0.4401; 0.7283) | Placebo | |
| -0.1657 (-0.6872; 0.3469) | 0.2799 (-0.5462; 1.1057) | 0.0245 (-0.6876; 0.7344) | -0.1184 (-0.5224; 0.2939) | Solanezumab |

#### TABLE S3.12. Diarrhea

##### Frequentist Network Meta-Analysis Results

| | | | | | |
|---|---|---|---|---|---|
| Aducanumab | | | | | |
| 0.1525 (-0.1495; 0.4547) | Bapineuzumab | | | | |
| 0.2994 (-0.1486; 0.7474) | 0.1469 (-0.3064; 0.6) | Donanemab | | | |
| 0.3641 (-0.0613; 0.7895) | 0.2115 (-0.2194; 0.6424) | 0.0647 (-0.4785; 0.6079) | Lecanemab | | |
| 0.1737 (-0.0344; 0.3818) | 0.0212 (-0.198; 0.2402) | -0.1257 (-0.5224; 0.271) | -0.1903 (-0.5614; 0.1807) | Placebo | |
| 0.1821 (-0.1197; 0.4839) | 0.0295 (-0.28; 0.339) | -0.1173 (-0.5704; 0.3356) | -0.182 (-0.6127; 0.2487) | 0.0084 (-0.2102; 0.227) | Solanezumab |

Heterogeneity $I^2$ = 0% [0.0%; 60.2%]; p-value of Q statistics = 0.7053 in heterogeneity (within designs).

##### Bayesian Network Meta-Analysis Results

| | | | | | |
|---|---|---|---|---|---|
| Aducanumab | | | | | |
| 0.1629 (-0.1779; 0.5289) | Bapineuzumab | | | | |
| 0.2989 (-0.2122; 0.8053) | 0.1303 (-0.3819; 0.6339) | Donanemab | | | |
| 0.3661 (-0.1571; 0.8927) | 0.2016 (-0.3295; 0.7207) | 0.0695 (-0.5662; 0.7102) | Lecanemab | | |
| 0.1756 (-0.0711; 0.4260) | 0.0113 (-0.2449; 0.2565) | -0.1218 (-0.5665; 0.3212) | -0.1899 (-0.6508; 0.2699) | Placebo | |
| 0.1802 (-0.1982; 0.5651) | 0.0161 (-0.3764; 0.3928) | -0.1170 (-0.6550; 0.4116) | -0.1854 (-0.7349; 0.3539) | 0.0036 (-0.2872; 0.2960) | Solanezumab |

#### TABLE S3.13. Cardiac disorders

##### Frequentist Network Meta-Analysis Results

| | | | |
|---|---|---|---|
| Aducanumab | | | |
| -0.4614 (-1.3645; 0.4418) | Bapineuzumab | | |
| -0.2221 (-0.8045; 0.3603) | 0.2393 (-0.4511; 0.9295) | Placebo | |
| -0.1226 (-1.0142; 0.7688) | 0.3387 (-0.6266; 1.3041) | 0.0995 (-0.5754; 0.7744) | Solanezumab |

Heterogeneity $I^2$ = 54.5% [0.0%; 79.4%]; p-value of Q statistics = 0.0315 in heterogeneity (within designs). Design-specific decomposition of within-designs: Placebo vs Solanezumab: p = 0.0106 and non-significant for other comparisons (p > 0.05).

##### Bayesian Network Meta-Analysis Results

| | | | |
|---|---|---|---|
| Aducanumab | | | |
| -0.4478 (-1.4294; 0.4915) | Bapineuzumab | | |
| -0.2156 (-0.8577; 0.3993) | 0.2329 (-0.4850; 0.9580) | Placebo | |
| -0.1403 (-1.0933; 0.8640) | 0.3065 (-0.6989; 1.3951) | 0.0696 (-0.6539; 0.8785) | Solanezumab |

#### TABLE S4.14. Nasopharyngitis

##### Frequentist Network Meta-Analysis Results

| | | | |
|---|---|---|---|
| Aducanumab | | | |
| 0.0268 (-0.2468; 0.3004) | Bapineuzumab | | |
| -0.0655 (-0.2209; 0.0898) | -0.0923 (-0.3175; 0.1328) | Placebo | |
| -0.09 (-0.3613; 0.1812) | -0.1169 (-0.4332; 0.1996) | -0.0245 (-0.2468; 0.1978) | Solanezumab |

Heterogeneity $I^2 = 6.7\%$ [0.0%; 69.8%]; p-value of Q statistics = 0.3785 in heterogeneity (within designs).

##### Bayesian Network Meta-Analysis Results

| | | | |
|---|---|---|---|
| Aducanumab | | | |
| 0.0268 (-0.2468; 0.3004) | Bapineuzumab | | |
| -0.064 (-0.2694; 0.1416) | -0.0956 (-0.3639; 0.1590) | Placebo | |
| -0.0886 (-0.4438; 0.2834) | -0.1188 (-0.5198; 0.2738) | -0.0234 (-0.3181; 0.2771) | Solanezumab |

#### TABLE S3.15. Upper respiratory infections

##### Frequentist Network Meta-Analysis Results

| | | | |
|---|---|---|---|
| Aducanumab | | | |
| -0.1401 (-0.4419; 0.1618) | Bapineuzumab | | |
| -0.0234 (-0.2219; 0.175) | 0.1166 (-0.1108; 0.344) | Placebo | |
| 0.0988 (-0.2255; 0.4232) | 0.2389 (-0.104; 0.5818) | 0.1222 (-0.1343; 0.3788) | Solanezumab |

Heterogeneity $I^2 = 0\%$ [0.0%; 70.8%]; p-value of Q statistics = 0.4300 in heterogeneity (within designs).

##### Bayesian Network Meta-Analysis Results

| | | | |
|---|---|---|---|
| Aducanumab | | | |
| -0.1420 (-0.5392, 0.2623) | Bapineuzumab | | |
| -0.0244 (-0.2860, 0.2405) | 0.1183 (-0.1859, 0.4201) | Placebo | |
| 0.1049 (-0.3284, 0.5512) | 0.2462 (-0.2180, 0.7207) | 0.1293 (-0.2211, 0.4860) | Solanezumab |

#### TABLE S3.16. Urinary infections

##### Frequentist Network Meta-Analysis Results

| | | | | | |
|---|---|---|---|---|---|
| Aducanumab | | | | | |
| -0.1569 (-0.4856; 0.1719) | Bapineuzumab | | | | |
| 0.1179 (-0.3501; 0.5859) | 0.2748 (-0.1809; 0.7305) | Donanemab | | | |
| -0.0608 (-0.4824; 0.3607) | 0.096 (-0.3117; 0.5039) | -0.1788 (-0.7052; 0.3478) | Lecanemab | | |
| -0.1119 (-0.3564; 0.1324) | 0.0449 (-0.1749; 0.2648) | -0.2299 (-0.629; 0.1692) | -0.0511 (-0.3945; 0.2923) | Placebo | |
| 0.0898 (-0.2631; 0.4426) | 0.2467 (-0.0896; 0.5831) | -0.0281 (-0.5015; 0.4453) | 0.1507 (-0.2768; 0.5781) | 0.2018 (-0.0528; 0.4563) | Solanezumab |

Heterogeneity $I^2$ = 12.3% [0.0%; 52.7%]; p-value of Q statistics = 0.3270 in heterogeneity (within designs).

##### Bayesian Network Meta-Analysis Results

| | | | | | |
|---|---|---|---|---|---|
| Aducanumab | | | | | |
| -0.1655 (-0.5778, 0.2202) | Bapineuzumab | | | | |
| 0.1216 (-0.4308, 0.6678) | 0.2843 (-0.2357, 0.8356) | Donanemab | | | |
| -0.0614 (-0.6160, 0.5013) | 0.1016 (-0.4248, 0.6663) | -0.1800 (-0.8475, 0.4777) | Lecanemab | | |
| -0.1104 (-0.4071, 0.1813) | 0.0528 (-0.2009, 0.3349) | -0.2306 (-0.6980, 0.2296) | -0.0504 (-0.5299, 0.4185) | Placebo | |
| 0.0920 (-0.3644, 0.5477) | 0.2559 (-0.1674, 0.7108) | -0.0276 (-0.6129, 0.5409) | 0.1512 (-0.4374, 0.7397) | 0.2027 (-0.1431, 0.5516) | Solanezumab |

#### TABLE S4. TERTIARY OUTCOMES

##### TABLE S4.1. Alzheimer's Disease Cooperative Study - Activities of Daily Living Scale for use in Mild Cognitive Impairment (ADCS-ADL-MCI)

League tables representing the results of network meta-analysis (NMA) (random-effect model). The values in the tables below represent relative effects (Standardized Mean Difference) for tertiary efficacy (functional) outcome ADCS-ADL-MCI. Bolded numbers indicate statistical significance ($p < 0.05$) for Frequentist NMA, or strong evidence in Bayesian NMA. The numbers in parentheses represent confidence intervals (CIs) for Frequentist NMA, or credible intervals (CrIs) in Bayesian framework. Dagger symbol (†) denotes when the interpretation differs significantly between frequentist and Bayesian NMA results, indicating the uncertainty and lack of strong evidence for a non-zero effect.

For each comparison, the number above 0 suggests that the drug in the column is superior to the drug in the row, and darker green colors correspond to greater relative effect.

###### Frequentist Network Meta-Analysis Results

| | | |
|---|---|---|
| Aducanumab | | |
| -0.0206 (-0.1475; 0.1063) | Lecanemab | |
| **0.1537 * (0.0741; 0.2333)** | **0.1743 † ** (0.0755; 0.2731)** | Placebo |

Statistical significance for Drug vs. Placebo: * p = 0.0002; ** p = 0.0005
Heterogeneity: $I^2 = 0\%$ [0.0%; 84.7%]; p-value of Q statistics = 0.4041 in heterogeneity (within designs).

###### Bayesian Network Meta-Analysis Results

| | | |
|---|---|---|
| Aducanumab | | |
| -0.0204 (-0.3005; 0.2648) | Lecanemab | |
| **0.1545 (0.0203, 0.2912)** | 0.1745 † (-0.0737; 0.4215) | Placebo |

#### TABLE S4.2. ARIA-H: Superficial Siderosis

League tables representing the results of network meta-analysis (NMA) (random-effect model). The values in the tables below represent Relative Risks (natural logarithm) for safety outcomes. Bolded numbers indicate statistical significance ($p < 0.05$) for Frequentist NMA, or strong evidence in Bayesian NMA. The numbers in parentheses represent confidence intervals (CIs) for Frequentist NMA, or credible intervals (CrIs) in Bayesian framework. Dagger symbol (†) denotes when the interpretation differs significantly between frequentist and Bayesian NMA results, indicating the uncertainty and lack of strong evidence for a non-zero effect.

For each comparison, the number below 0 suggests that the drug in the column is safer than the drug in the row, and darker red colors correspond to relatively higher risk.

##### Frequentist Network Meta-Analysis Results

| | | | |
|---|---|---|---|
| Aducanumab | | | |
| 0.008 (-0.5148; 0.5309) | Donanemab | | |
| **0.798 † (0.1956; 1.4005)** | **0.79 † (0.1235; 1.4565)** | Lecanemab | |
| **1.6644 * (1.3544; 1.9744)** | **1.6564 * (1.2353; 2.0776)** | **0.8664 † ** (0.3498; 1.383)** | Placebo |

Statistical significance for Drug vs. Placebo: * $p < 0.0001$; ** $p = 0.0010$

Heterogeneity: $I^2 = 3.9\%$ [0.0%; 80.0%]; p-value of Q statistics = 0.3843 in heterogeneity (within designs).

##### Bayesian Network Meta-Analysis Results

| | | | |
|---|---|---|---|
| Aducanumab | | | |
| -0.0090 (-0.9657, 0.8824) | Donanemab | | |
| 0.8016 † (-0.3199, 1.9751) | 0.8101 † (-0.3994, 2.1391) | Lecanemab | |
| **1.6663 (1.1478, 2.2141)** | **1.6766 (0.9512, 2.4798)** | 0.8672 † (-0.1800, 1.8663) | Placebo |

#### TABLE S4.3. Infusion-related reactions

##### Frequentist Network Meta-Analysis Results

| | | |
|---|---|---|
| Donanemab | | |
| **1.5771 † (0.5382; 2.6159)** | Lecanemab | |
| **2.8544 * (1.8477; 3.861)** | **1.2773 † * (1.0207; 1.5339)** | Placebo |

Statistical significance for Drug vs. Placebo: * $p < 0.0001$

Heterogeneity: $I^2 = 0\%$ [0.0%; 60.2%]; p-value of Q statistics = 0.3825 in heterogeneity (within designs).

##### Bayesian Network Meta-Analysis Results

| | | |
|---|---|---|
| Donanemab | | |
| 1.581 † (-2.8347; 5.8902) | Lecanemab | |
| **2.8567 (0.2347; 5.4776)** | 1.2788 † (-2.2187; 4.8008) | Placebo |

#### TABLE S4.4. Total events of ARIA-E (cerebral edema) in APOE-ε4 carriers

##### Frequentist Network Meta-Analysis Results

| | | | | |
|---|---|---|---|---|
| Aducanumab | | | | |
| -1.4351 (-3.4327; 0.5614) | Bapineuzumab | | | |
| 0.3258 (-0.3157; 0.9674) | 1.7611 (-0.2769; 3.7992) | Donanemab | | |
| **0.8612 † (0.2106; 1.5119)** | **2.2965 † (0.2555; 4.3375)** | 0.5354 (-0.2331; 1.3038) | Lecanemab | |
| **2.7925 * (2.4429; 3.1422)** | **4.2278 * (2.262; 6.1936)** | **2.4667 * (1.9287; 3.0046)** | **1.9313 * (1.3826; 2.48)** | Placebo |

Statistical significance for Drug vs. Placebo: * p < 0.0001

Heterogeneity $I^2$ = 0% [0.0%; 84.7%]; p-value of Q statistics = 0.6529 in heterogeneity (within designs).

| HOMOZYGOTES | | | HETEROZYGOTES | | |
|---|---|---|---|---|---|
| Donanemab | | | Donanemab | | |
| 0.311 (-0.9449; 1.5668) | Lecanemab | | 0.7331 (-0.2324; 1.6986) | Lecanemab | |
| **2.4718 † * (1.5878; 3.3557)** | **2.1608 † * (1.2688; 3.0528)** | Placebo | **2.485 † * (1.8161; 3.154)** | **1.7519 † * (1.0558; 2.4481)** | Placebo |

Statistical significance for Drug vs. Placebo: * p < 0.0001

##### Bayesian Network Meta-Analysis Results

| | | | | |
|---|---|---|---|---|
| Aducanumab | | | | |
| -1.4292 (-3.8130, 0.9670) | Bapineuzumab | | | |
| 0.3304 (-1.1360, 1.8003) | 1.7512 (-0.8506, 4.3838) | Donanemab | | |
| 0.8715 † (-0.6021, 2.3868) | 2.2945 † (-0.3280, 4.9222) | 0.5437 (-1.3111, 2.3945) | Lecanemab | |
| **2.7957 (2.1271, 3.4753)** | **4.2223 (1.9215, 6.5246)** | **2.4649 (1.1641, 3.7608)** | **1.9247 (0.6004, 3.2173)** | Placebo |

| HOMOZYGOTES | | | HETEROZYGOTES | | |
|---|---|---|---|---|---|
| Donanemab | | | Donanemab | | |
| 0.2961 (-4.209, 4.8414) | Lecanemab | | 0.7356 (-3.6955, 5.2277) | Lecanemab | |
| 2.46838 † (-0.7311, 5.6601) | 2.1749 † (-1.0334, 5.3421) | Placebo | 2.48038 † (-0.6836, 5.6226) | 1.7427 † (-1.4145, 4.9626) | Placebo |

#### TABLE S4.5. Total events of ARIA-E (cerebral edema) in APOE-ε4 non-carriers

##### Frequentist Network Meta-Analysis Results

| | | | | |
|---|---|---|---|---|
| Aducanumab | | | | |
| **-1.9166 (-3.3932; -0.4399)** | Bapineuzumab | | | |
| -1.3951 (-2.8647; 0.0736) | 0.5217 (-1.4775; 2.5208) | Donanemab | | |
| -1.1555 (-3.2139; 0.9038) | 0.7612 (-1.7048; 3.2271) | 0.2395 (-2.2219; 2.7005) | Lecanemab | |
| **1.5808 * (1.1673; 1.9943)** | **3.4976 * (2.0798; 4.9154)** | **2.9759 * (1.5666; 4.3853)** | **2.7364 ** (0.7189; 4.7539)** | Placebo |

Statistical significance for Drug vs. Placebo: * $p < 0.0001$; ** $p = 0.0079$

Heterogeneity $I^2 = 0\%$ [0.0%; 79.2%]; p-value of Q statistics = 0.9348 in heterogeneity (within designs).

##### Bayesian Network Meta-Analysis Results

| | | | | |
|---|---|---|---|---|
| Aducanumab | | | | |
| **-1.9208 (-3.6298, -0.2018)** | Bapineuzumab | | | |
| -1.4098 (-3.2073, 0.4174) | 0.5265 (-1.8008, 2.8489) | Donanemab | | |
| -1.1439 (-3.4552, 1.1798) | 0.7736 (-1.9838, 3.5052) | 0.2609 (-2.5521, 3.0664) | Lecanemab | |
| **1.5783 (0.9377, 2.2032)** | **3.4974 (1.9069, 5.0693)** | **2.9847 (1.2685, 4.6738)** | **2.7208 (0.4912, 4.9368)** | Placebo |

#### TABLE S4.6. Total events of ARIA-H in APOE-ε4 carriers

Including all incidents of cerebral microhemorrhages, macrohemorrhages and superficial siderosis.

##### Frequentist Network Meta-Analysis Results

| | | |
|---|---|---|
| Donanemab | | |
| **0.4045 † (0.0526; 0.7563)** | Lecanemab | |
| **0.9597 † (0.7381; 1.1814)** | **0.5553 † (0.282; 0.8286)** | Placebo |

Statistical significance for Drug vs. Placebo: * $p < 0.0001$

**APOE-ε4 HOMOZYGOTES**

| | | |
|---|---|---|
| Donanemab | | |
| 0.2795 (-0.2488; 0.8079) | Lecanemab | |
| **0.8962 † * (0.5381; 1.2543)** | **0.6167 † ** (0.2283; 1.0052)** | Placebo |

**APOE-ε4 HETEROZYGOTES**

| | | |
|---|---|---|
| Donanemab | | |
| **0.4991 † (0.0385; 0.9596)** | Lecanemab | |
| **0.9881 † * (0.7104; 1.2658)** | **0.489 † *** (0.1217; 0.8564)** | Placebo |

Statistical significance for Drug vs. Placebo: * $p < 0.0001$; ** $p = 0.0019$; *** $p = 0.0091$

##### Bayesian Network Meta-Analysis Results

| | | |
|---|---|---|
| Donanemab | | |
| 0.4035 † (-1.3280, 2.1454) | Lecanemab | |
| 0.9593 † (-0.2623, 2.1748) | 0.5552 † (-0.6757, 1.7771) | Placebo |

**APOE-ε4 HOMOZYGOTES**

| | | |
|---|---|---|
| Donanemab | | |
| 0.285 (-1.3817, 1.9324) | Lecanemab | |
| 0.8935 † (-0.2638, 2.0532) | 0.6110 † (-0.5784, 1.7952) | Placebo |

**APOE-ε4 HETEROZYGOTES**

| | | |
|---|---|---|
| Donanemab | | |
| 0.4973 † (-1.3142, 2.2817) | Lecanemab | |
| 0.9866 † (-0.2676, 2.2491) | 0.4906 † (-0.7837, 1.7736) | Placebo |

#### TABLE S4.7. Total events of ARIA-H in APOE-ε4 non-carriers

Including all incidents of cerebral microhemorrhages, macrohemorrhages and superficial siderosis.

##### Frequentist Network Meta-Analysis Results

| | | |
|---|---|---|
| Donanemab | | |
| -0.5207 (-1.2928; 0.2513) | Lecanemab | |
| **0.5192 † * (0.087; 0.9514)** | **1.04 † ** (0.4003; 1.6797)** | Placebo |

Statistical significance for Drug vs. Placebo: * $p = 0.0185$; ** $p = 0.0014$

##### Bayesian Network Meta-Analysis Results

| | | |
|---|---|---|
| Donanemab | | |
| -0.5248 (-2.4876, 1.4291) | Lecanemab | |
| 0.5191 † (-0.8382, 1.8825) | 1.0367 † (-0.3819, 2.4616) | Placebo |

#### TABLE S5. Results of influence analyses and GOSH plot diagnostics

Abbreviations: DBSCAN (Density-Based Spatial Clustering of Applications with Noise); GMM (Gaussian Mixture Model); GOSH (Graphic Display of Study Heterogeneity)

| **Efficacy Outcomes** | |
|---|---|
| Method | Influential observations / Studies |
| Leave-one-out meta-analysis | Sims et al (Low-medium tau) (ADAS-Cog, CDR-SB)<br>Clarity AD (Amyloid burden on PET)<br>Salloway et al 2 (ADAS-Cog)<br>Salloway et al 3 (MMSE and CDR-SB)<br>EXPEDITION 3 (MMSE) |
| Baujat and Influence Diagnostics | Sims et al (Low-medium tau) (ADAS-Cog, CDR-SB)<br>Clarity AD (Amyloid burden on PET) |
| k-means clustering | Sims et al (Low-medium tau), EMERGE2 (ADAS-Cog)<br>Sims et al (Low-medium tau) (CDR-SB)<br>Salloway et al. 3 (MMSE, CDR-SB)<br>Clarity AD (Amyloid burden on PET) |
| DBSCAN | Salloway et al. 2 (ADAS-Cog),<br>Sims et al (Low-medium tau), EMERGE2 (ADAS-Cog, CDR-SB)<br>Salloway et al. 3 (MMSE, CDR-SB)<br>Clarity AD (CDR-SB, Amyloid burden on PET) |
| GMM | Salloway et al. 2 (ADAS-Cog),<br>Sims et al (Low-medium tau), EMERGE2 (ADAS-Cog, CDR-SB)<br>Salloway et al. 3 (MMSE, CDR-SB)<br>Clarity AD (CDR-SB, Amyloid burden on PET) |
| **Safety Outcomes** | |
| Leave-one-out meta-analysis. | ENGAGE1 (Fall, Total ARIA-E, Back Pain)<br>Sims et al (Low/medium tau), EXPEDITION 1 and 2 (T. Discontinuation)<br>EXPEDITION 1 and 2 (Total ARIA-H, ARIA-H (Microhemorrhages))<br>EXPEDITION 1 (Total ARIA-E)<br>EXPEDITION 3 (Serious adverse events)<br>Vandenberghe et al 3 (Headaches, Urinary Infections) |
| Baujat and Influence Diagnostics | ENGAGE 1 (Fall, Back Pain, Nausea)<br>Sims et al (pooled) (ARIA-E in APOE-ε4 carriers)<br>Sims et al Low/medium tau (T. Discontinuation), Total ARIA-H<br>All Aducanumab studies (ARIA-E in APOE-ε4 non-carriers)<br>Vandenberghe et al 2 (Diarrhea)<br>Vandenberghe et al 3 (Headaches)<br>EXPEDITION 1 and 2 (Total ARIA-E, Total ARIA-H, ARIA-H (Microhemorrhages), Syncope)<br>EXPEDITION 3 (Serious adverse events, Total ARIA-E, Fatigue) |
| k-means clustering | Sims et al. Low/medium tau (T. Discontinuation)<br>Vandenberghe et al. 1 (Total ARIA-E and ARIA-H (Microhemorrhages))<br>Vandenberghe et al 2 (Urinary infections)<br>Vandenberghe et al 3 (Headaches, Dizziness, Urinary infections)<br>ENGAGE1 (Fall, Back Pain, Nausea)<br>ENGAGE2 (Fall)<br>EXPEDITION 1 and 2 (Total ARIA-H, ARIA-H (Microhemorrhages))<br>EXPEDITION 3 (Diarrhea, Fatigue, Syncope, ARIA-H Microhemorrhages)<br>Salloway et al 2 Study 301 (ARIA-E in APOE-ε4 non-carriers)<br>Salloway et al 3 Study 302 (ARIA-E in APOE-ε4 carriers)<br>Clarity AD (Amyloid PET, Serious adverse events, ARIA-H (Microhemorrhages)) |

| | |
|---|---|
| DBSCAN | Sims et al. Low/medium and High tau (T. Discontinuation, Fatigue)<br>Sims et al High tau (Nausea)<br>ENGAGE1 (Back Pain, Fall)<br>ENGAGE1, Sims et al (high tau) (Nausea)<br>ENGAGE1, EMERGE 1, Vandenberghe et al 2 (Dizziness)<br>EMERGE2, ENGAGE2 (ARIA-H (Microhemorrhages))<br>ENGAGE2 (Fall)<br>EMERGE1, EXPEDITION 3 (Serious adverse events)<br>Vandenberghe et al 1 (Total ARIA-E, Diarrhea)<br>EXPEDITION 1 and 2 (Diarrhea, Total ARIA-H, ARIA-H (Microhemorrhages))<br>EXPEDITION 3 (Syncope, Fatigue, ARIA-H (Microhemorrhages))<br>Salloway et al 3 (2014) Study 302 (ARIA-E in APOE-ε4 carriers)<br>Salloway et al 2 Study 301 (ARIA-E in APOE-ε4 non-carriers)<br>Clarity AD (Amyloid PET, Serious adverse events, Dizziness)<br>Vandenberghe et al 2 (Arthralgia, Urinary infections)<br>Vandenberghe et al 3 (Arthralgia, Headaches, Dizziness, ARIA-H (Microhemorrhages), Urinary infections) |
| GMM | Sims et al. Low/medium and High tau (T. Discontinuation)<br>Sims et al High tau (Nausea)<br>ENGAGE1, Sims et al (high tau) (Nausea)<br>ENGAGE1 (Back Pain, Fall)<br>ENGAGE1, Clarity AD, Salloway et al 2 Study 301 (ARIA-E in APOE-ε4 non-carriers)<br>ENGAGE2 (Fall)<br>EXPEDITION 1 and 2, EMERGE1 (Total ARIA-H)<br>EXPEDITION 1 and 2 (T. Discontinuation, Fall, Total ARIA-H, ARIA-H (Microhemorrhages))<br>EXPEDITION 3 (Syncope, Fatigue)<br>Salloway et al 3 (2014) Study 302 (Total ARIA-E, ARIA-E in APOE-ε4 carriers, Back Pain))<br>Vandenberghe et al 1 (Headaches, Total ARIA-E)<br>Vandenberghe et al 2, EMERGE2 (Diarrhea, Urinary infections)<br>Vandenberghe et al 2 (Arthralgia)<br>Vandenberghe et al 3 (Arthralgia, Headaches, Dizziness, Urinary infections)<br>Clarity AD (Amyloid PET, Serious adverse events) |

#### TABLE S6. The Impact of COVID-19

| Author (Year): Trial Name | Drug | Number (%) of patients affected by COVID-19 disease (Drug vs. Placebo) |
|---|---|---|
| van Dyck et al (2023) Clarity AD | Lecanemab | 64 (7.1) vs. 60 (6.7) |
| Sims et al (2023) TRAILBLAZER-ALZ 2 Low/medium-tau population | Donanemab | 94 (16.1) vs. 106 (17.9) |
| Sims et al (2023) TRAILBLAZER-ALZ 2 High-tau population | Donanemab | 42 (15.7) vs. 48 (17.1) |

**Clarity AD Study (Lecanemab)**: COVID-19 pandemic caused missing doses during the trial, postponed assessments, and intercurrent illnesses. In accordance with previous agreement with the FDA, the sample size was increased by 200 to retain 90% power, for a total sample size of approximately 1766 randomized subjects to account for participants who missed three or more consecutive doses during the initial 6-month peak period of COVID-19 disease. Sensitivity analyses of the CDR-SB score that evaluated the effect of COVID-19 (missed doses) were generally consistent with the primary analysis.

**TRAILBLAZER-ALZ 2 Study (Donanemab)** COVID-19 was the most commonly reported adverse event across treatment groups. The pandemic also caused staffing difficulties and delays in start-up activities (4-6 weeks), and delays in enrollment and study visits. Treatment emergent COVID-19 adverse events, including the discontinuation and missed visits due to COVID-19 were analyzed. Since the study started after onset of the COVID-19 pandemic, elements intended to mitigate the impact of COVID-19 were built into the protocol or subsequent amendments. Adjustments in study procedures included site visits conducted over telephone to collect preexisting conditions and adverse events, concomitant medications as well as cognitive/functional assessments.

#### TABLE S7. SENSITIVITY ANALYSIS: Efficacy Outcomes

League tables representing the results of network meta-analysis (NMA) (random-effect model). The values in the tables below represent relative effects (Standardized Mean Difference) for efficacy outcomes. Bolded numbers indicate statistical significance (p < 0.05) for Frequentist NMA, or strong evidence in Bayesian NMA. The numbers in parentheses represent confidence intervals (CIs) for Frequentist NMA, or credible intervals (CrIs) in Bayesian framework. Dagger symbol (†) denotes when the interpretation differs significantly between frequentist and Bayesian NMA results, indicating the uncertainty and lack of strong evidence for a non-zero effect.

##### TABLE S7.1. The Alzheimer's Disease Assessment Scale–Cognitive Subscale (ADAS Cog)

Sensitivity analysis after excluding two influential studies: high-dose Aducanumab EMERGE study and Donanemab study in low/medium *tau* load.

For each comparison, the number below 0 suggests that the drug in the column is superior to the drug in the row, and darker green colors correspond to greater relative effect.

###### Frequentist Network Meta-Analysis Results

| | | | | | |
|---|---|---|---|---|---|
| Aducanumab | | | | | |
| -0.0694<br>(-0.1778; 0.0390) | Bapineuzumab | | | | |
| -0.0275<br>(-0.2464; 0.1913) | 0.0419<br>(-0.1658; 0.2495) | Donanemab | | | |
| 0.0249<br>(-0.1062; 0.1560) | 0.0943<br>(-0.0171; 0.2057) | 0.0524<br>(-0.1679; 0.2728) | Lecanemab | | |
| -0.0908 *<br>(-0.1817; 0.0001) | -0.0214<br>(-0.0805; 0.0376) | -0.0633<br>(-0.2623; 0.1358) | **-0.1157** † **<br>**(-0.2101; -0.0213)** | Placebo | |
| -0.0237<br>(-0.1330; 0.0856) | 0.0457<br>(-0.0390; 0.1304) | 0.0038<br>(-0.2043; 0.2120) | -0.0486<br>(-0.1609; 0.0637) | **0.0671** † ***<br>**(0.0064; 0.1278)** | Solanezumab |

Statistical significance for Drug vs. Placebo: * p = 0.0502; ** p = 0.0163; *** p = 0.0303
$I^2$ = 0% [0.0%; 62.4%]; p-value of Q statistics = 0.9223 in heterogeneity (within designs).

###### Bayesian Network Meta-Analysis Results

| | | | | | |
|---|---|---|---|---|---|
| Aducanumab | | | | | |
| -0.0697<br>(-0.1892, 0.0508) | Bapineuzumab | | | | |
| -0.0296<br>(-0.2580, 0.2008) | 0.0397<br>(-0.1801, 0.2617) | Donanemab | | | |
| 0.0247<br>(-0.1309, 0.1791) | 0.0938<br>(-0.0410, 0.2304) | 0.0555<br>(-0.1854, 0.2916) | Lecanemab | | |
| -0.0912<br>(-0.1920, 0.0076) | -0.0213<br>(-0.0887, 0.0443) | -0.0613<br>(-0.2719, 0.1477) | -0.1158 †<br>(-0.2334, 0.0024) | Placebo | |
| -0.0240<br>(-0.1478, 0.1016) | 0.0463<br>(-0.0533, 0.1456) | 0.0065<br>(-0.2166, 0.2274) | -0.0480<br>(-0.1869, 0.0909) | 0.0675 †<br>(-0.0057, 0.1414) | Solanezumab |

#### TABLE S7.2. The Clinical Dementia Rating Scale Sum of Boxes (CDR-SB)

Sensitivity analysis after excluding two influential studies: high-dose Aducanumab EMERGE study and Donanemab study in low/medium *tau* load.

For each comparison, the number below 0 suggests that the drug in the column is superior to the drug in the row, and darker green colors correspond to greater relative effect.

##### Frequentist Network Meta-Analysis Results

| | | | | | |
|---|---|---|---|---|---|
| Aducanumab | | | | | |
| -0.0750 (-0.1852; 0.0353) | Bapineuzumab | | | | |
| 0.1642 (-0.0604; 0.3889) | **0.2392 (0.0264; 0.4520)** | Donanemab | | | |
| 0.0663 (-0.0712; 0.2037) | **0.1412 (0.0243; 0.2582)** | -0.0980 (-0.3260; 0.1301) | Lecanemab | | |
| -0.0698 (-0.1630; 0.0234) | 0.0051 (-0.0538; 0.0641) | **-0.2340 * (-0.4385; -0.0296)** | **-0.1361 † ** (-0.2371; -0.0350)** | Placebo | |
| -0.0159 (-0.1306; 0.0987) | 0.0590 (-0.0301; 0.1481) | -0.1802 (-0.3953; 0.0349) | -0.0822 (-0.2034; 0.0390) | 0.0539 (-0.0130; 0.1207) | Solanezumab |

Statistical significance for Drug vs. Placebo: * p = 0.0249; ** p = 0.0083

$I^2$ = 6.8% [0.0%; 64.9%]; p-value of Q statistics = 0.3788 in heterogeneity (within designs).

##### Bayesian Network Meta-Analysis Results

| | | | | | |
|---|---|---|---|---|---|
| Aducanumab | | | | | |
| -0.0731 (-0.2043, 0.0583) | Bapineuzumab | | | | |
| 0.166 (-0.0841; 0.4175) | **0.2378 (0.0003; 0.4766)** | Donanemab | | | |
| 0.067 (-0.1141; 0.24596) | 0.14 (-0.0221; 0.2981) | -0.0985 (-0.3683; 0.1699) | Lecanemab | | |
| -0.0692 (-0.1815; 0.0403) | 0.00403 (-0.0704: 0.0749) | **-0.2343 (-0.4633; -0.0090)** | -0.1359 † (-0.28; 0.0073) | Placebo | |
| -0.018 (-0.1639; 0.1198) | 0.0551 (-0.0654; 0.1658) | -0.183 (-0.4291; 0.0572) | -0.0822 (-0.2034; 0.0390) | 0.0539 (-0.0130; 0.1207) | Solanezumab |

#### TABLE S7.3. The Mini Mental Examination (MMSE)

Sensitivity analysis after excluding two influential studies: high-dose Aducanumab EMERGE study and Donanemab study in low/medium *tau* load.

For each comparison, the number above 0 suggests that the drug in the column is superior to the drug in the row, and darker green colors correspond to greater relative effect.

##### Frequentist Network Meta-Analysis Results

| | | | | |
|---|---|---|---|---|
| Aducanumab | | | | |
| -0.0090 (-0.1281; 0.1101) | Bapineuzumab | | | |
| -0.0901 (-0.3107; 0.1305) | -0.0811 (-0.2966; 0.1344) | Donanemab | | |
| 0.0072 (-0.0833; 0.0978) | 0.0162 (-0.0611; 0.0936) | 0.0973 (-0.1038; 0.2985) | Placebo | |
| -0.0804 (-0.1909; 0.0302) | -0.0714 (-0.1714; 0.0287) | 0.0097 (-0.2012; 0.2206) | **-0.0876 * (-0.1511; -0.0242)** | Solanezumab |

Statistical significance for Drug vs. Placebo: * p = 0.0068

$I^2$ = 0% [0.0%; 70.8%]; p-value of Q statistics = 0.8483 in heterogeneity (within designs).

##### Bayesian Network Meta-Analysis Results

| | | | | |
|---|---|---|---|---|
| Aducanumab | | | | |
| -0.0111 (-0.1478, 0.1260) | Bapineuzumab | | | |
| -0.0900 (-0.3311, 0.1485) | -0.0785 (-0.3144, 0.1549) | Donanemab | | |
| 0.0070 (-0.0946, 0.1097) | 0.0177 (-0.0724, 0.1089) | 0.0968 (-0.1187, 0.3131) | Placebo | |
| -0.0797 (-0.2086, 0.0516) | -0.0682 (-0.1889, 0.0529) | 0.0106 (-0.2198, 0.2409) | **-0.0864 (-0.1664, -0.0061)** | Solanezumab |

#### TABLE S7.4. Amyloid burden on PET

Sensitivity analysis after excluding two influential studies: high-dose Aducanumab EMERGE study and Donanemab study in low/medium *tau* load.

For each comparison, the number below 0 suggests that the drug in the column is superior to the drug in the row, and darker green colors correspond to greater relative effect.

##### Frequentist Network Meta-Analysis Results

| | | | |
|---|---|---|---|
| Aducanumab | | | |
| -0.3714 (-2.1893; 1.4464) | Donanemab | | |
| **3.5730 † (1.6985; 5.4475)** | **3.9444 † (1.6843; 6.2045)** | Lecanemab | |
| **-4.2267 (-5.1514; -3.3020)** | **-3.8553 † (-5.4204; -2.2902)** | **-7.7997 (-9.4303; -6.1692)** | Placebo |

Statistical significance for Drug vs. Placebo: * p = 0.0001
$I^2$ = 91.3% [77.7%; 96.6%]; p-value of Q statistics < 0.0001 in heterogeneity (within designs). Design-specific decomposition of within-designs.

##### Bayesian Network Meta-Analysis Results

| | | | |
|---|---|---|---|
| Aducanumab | | | |
| -0.3657 (-6.8581; 6.0830) | Donanemab | | |
| 3.5588 † (-3.1157; 10.0763) | 3.9218 † (-4.1589; 11.9339) | Lecanemab | |
| **-4.2210 (-7.5093; -0.9741)** | **-3.8638 † (-9.3948; 1.7020)** | **-7.7814 (-13.4679, -1.9530)** | Placebo |

#### TABLE S7.5. CSF Aβ1-42

Sensitivity analysis after excluding high-dose Aducanumab EMERGE study.

For each comparison, the number above 0 suggests that the drug in the column is superior to the drug in the row, and darker green colors correspond to greater relative effect.

##### Frequentist Network Meta-Analysis Results

| | | | |
|---|---|---|---|
| Aducanumab | | | |
| -0.0627 (-0.6273; 0.5018) | Lecanemab | | |
| **1.0429 * (0.6377; 1.4481)** | **1.1056 * (0.7126; 1.4986)** | Placebo | |
| **1.0030 † (0.4302; 1.5757)** | **1.0657 † (0.5015; 1.6300)** | -0.0399 (-0.4447; 0.3649) | Solanezumab |

Statistical significance for Drug vs. Placebo: * p < 0.0001
$I^2$ = 15% [0.0%; 87.0%]; p-value of Q statistics = 0.3170 in heterogeneity (within designs).

##### Bayesian Network Meta-Analysis Results

| | | | |
|---|---|---|---|
| Aducanumab | | | |
| -0.0684 (-1.3471; 1.1902) | Lecanemab | | |
| **1.0358 (0.3389; 1.7066)** | **1.1050 (0.0439; 2.1693)** | Placebo | |
| 1.0010 † (-0.0429; 2.0671) | 1.0693 † (-0.2384; 2.4425) | -0.0343 (-0.8221; 0.7990) | Solanezumab |

#### TABLE S7.6. CSF p-tau

Sensitivity analysis after excluding high-dose Aducanumab EMERGE study.

For each comparison, the number below 0 suggests that the drug in the column is superior to the drug in the row, and darker green colors correspond to greater relative effect.

##### Frequentist Network Meta-Analysis Results

| | | | |
|---|---|---|---|
| Aducanumab | | | |
| -0.3468 (-0.7469; 0.0534) | Bapineuzumab | | |
| -0.0485 (-0.5014; 0.4044) | 0.298 (-0.0396; 0.6361) | Lecanemab | |
| **-0.5403 * (-0.8946; -0.1860)** | **-0.1935 † ** (-0.3795; -0.0076)** | **-0.4918 † *** (-0.8063;-0.1773)** | Placebo |

Statistical significance for Drug vs. Placebo: * p = 0.0028; ** p = 0.0414; *** p = 0.0006
$I^2$ = 0% [0.0%; 79.2%]; p-value of Q statistics = 0.4863 in heterogeneity (within designs).

##### Bayesian Network Meta-Analysis Results

| | | | |
|---|---|---|---|
| Aducanumab | | | |
| -0.3380 (-0.8972, 0.2477) | Bapineuzumab | | |
| -0.0343 (-0.7755, 0.7355) | 0.3011 (-0.3864, 1.0089) | Lecanemab | |
| **-0.5278 (-0.9772, -0.0540)** | **-0.1900 † (-0.5392, 0.1701)** | -0.4919 (-1.0985, 0.1068) | Placebo |

#### TABLE S7.7. The Alzheimer's Disease Cooperative Study - Activities of Daily Living Scale for use in Mild Cognitive Impairment (ADCS-ADL-MCI)

For each comparison, the number above 0 suggests that the drug in the column is superior to the drug in the row, and darker green colors correspond to greater relative effect.

Sensitivity analysis after excluding high-dose Aducanumab EMERGE study.

##### Frequentist Network Meta-Analysis Results

| | | |
|---|---|---|
| Aducanumab | | |
| -0.0590 (-0.1934; 0.0754) | Lecanemab | |
| **0.1153 † * (0.0243; 0.2063)** | **0.1743 † ** (0.0755; 0.2731)** | Placebo |

Statistical significance for Drug vs. Placebo: * p = 0.0131; ** p = 0.0005
$I^2$ = 0% [0.0%; 89.6%]; p-value of Q statistics = 0.9908 in heterogeneity (within designs).

##### Bayesian Network Meta-Analysis Results

| | | |
|---|---|---|
| Aducanumab | | |
| -0.0578 (-0.2873; 0.1715) | Lecanemab | |
| 0.1157 † (-0.0138; 0.2446) | 0.174 † (-0.0176; 0.3635) | Placebo |

#### TABLE S8. SENSITIVITY ANALYSIS: Safety Outcomes

League tables representing the results of network meta-analysis (NMA) (random-effect model). The values in the tables below represent Relative Risks (natural logarithm) for safety outcomes. Bolded numbers indicate statistical significance ($p < 0.05$) for Frequentist NMA, or strong evidence in Bayesian NMA. The numbers in parentheses represent confidence intervals (CIs) for Frequentist NMA, or credible intervals (CrIs) in Bayesian framework. Dagger symbol (†) denotes when the interpretation differs significantly between frequentist and Bayesian NMA results, indicating the uncertainty and lack of strong evidence for a non-zero effect.

For each comparison, the number below 0 suggests that the drug in the column is safer than the drug in the row, and darker red colors correspond to relatively higher risk.

##### TABLE S8.1. Headaches

Sensitivity analysis after excluding two low-dose Bapineuzumab studies (Vandenberghe et al., 2016) that contributed to high heterogeneity in the main analysis.

###### Frequentist Network Meta-Analysis Results

| | | | | | |
|---|---|---|---|---|---|
| Aducanumab | | | | | |
| 0.159 (-0.0793; 0.3972) | Bapineuzumab | | | | |
| -0.0944 (-0.3861; 0.1971) | -0.2535 (-0.5818; 0.0748) | Donanemab | | | |
| -0.0579 (-0.3734; 0.2575) | -0.2169 (-0.5665; 0.1327) | 0.0365 (-0.3514; 0.4245) | Lecanemab | | |
| **0.2556 * (0.1252; 0.3861)** | 0.0967 (-0.1027; 0.2961) | **0.3501 ** (0.0893; 0.611)** | **0.3136 † *** (0.0264; 0.6008)** | Placebo | |
| **0.2868 (0.0403; 0.5333)** | 0.1278 (-0.1612; 0.4168) | **0.3812 (0.0469; 0.7156)** | 0.3447 (-0.0106; 0.7) | 0.0311 (-0.1781; 0.2403) | Solanezumab |

Statistical significance for Drug vs. Placebo: * p = 0.0001; ** p = 0.0085; *** p = 0.0323
Heterogeneity $I^2 = 0\%$ [0.0%; 64.8%]; p-value of Q statistics = 0.9858 in heterogeneity (within designs).

###### Bayesian Network Meta-Analysis Results

| | | | | | |
|---|---|---|---|---|---|
| Aducanumab | | | | | |
| 0.1577 (-0.1129; 0.426) | Bapineuzumab | | | | |
| -0.0976 (-0.4234; 0.2287) | -0.2528 (-0.62; 0.106) | Donanemab | | | |
| -0.0581 (-0.4197; 0.307) | -0.2166 (-0.6092; 0.179) | 0.0388 (-0.3936: 0.4744) | Lecanemab | | |
| **0.254 (0.1012; 0.4079)** | 0.0961 (-0.12; 0.3154) | **0.3503 (0.065; 0.6372)** | 0.314 † (-0.016; 0.6388) | Placebo | |
| **0.287 (0.003; 0.5711)** | 0.1303 (-0.1923; 0.4476) | **0.3834 (0.0089; 0.7575)** | 0.3456 (-0.0642; 0.7528) | 0.0319 (-0.2046; 0.2709) | Solanezumab |

#### TABLE S8.2. Falls

Sensitivity analysis after excluding ENGAGE studies of Aducanumab.

##### Frequentist Network Meta-Analysis Results

| | | | | | |
|---|---|---|---|---|---|
| Aducanumab | | | | | |
| 0.0532 (-0.2134; 0.3196) | Bapineuzumab | | | | |
| -0.0425 (-0.3687; 0.2837) | -0.0956 (-0.3854; 0.1942) | Donanemab | | | |
| -0.0603 (-0.4126; 0.2921) | -0.1134 (-0.4323; 0.2055) | -0.0178 (-0.388; 0.3525) | Lecanemab | | |
| 0.0169 (-0.1993; 0.233) | -0.0362 (-0.1921; 0.1196) | 0.0594 (-0.1849; 0.3037) | 0.0771 (-0.2011; 0.3554) | Placebo | |
| -0.0016 (-0.2967; 0.2935) | -0.0548 (-0.309; 0.1995) | 0.0409 (-0.2754; 0.3571) | 0.0587 (-0.2011; 0.4018) | -0.0185 (-0.2193; 0.1824) | Solanezumab |

Heterogeneity $I^2$ = 0% [0.0%; 67.6%]; p-value of Q statistics = 0.9837 in heterogeneity (within designs).

##### Bayesian Network Meta-Analysis Results

| | | | | | |
|---|---|---|---|---|---|
| Aducanumab | | | | | |
| 0.0538 (-0.2343; 0.3402) | Bapineuzumab | | | | |
| -0.0406 (-0.3944; 0.3169) | -0.094 (-0.4057; 0.2209) | Donanemab | | | |
| -0.0625 (-0.4465; 0.3286) | -0.1157 (-0.4649; 0.2343) | -0.0214 (-0.4285; 0.3903) | Lecanemab | | |
| 0.016 (-0.2202; 0.2517) | -0.0379 (-0.2053; 0.1286) | 0.0567 (-0.2063; 0.3228) | 0.0771 (-0.2306; 0.3845) | Placebo | |
| -0.0055 (-0.3441; 0.3383) | -0.0587 (-0.3501; 0.2422) | 0.0381 (-0.3206; 0.3930) | 0.0566 (-0.33; 0.450) | -0.02 (-0.2618; 0.2273) | Solanezumab |

#### REFERENCES

Beis G & Papasotiriou I. (2023). Is Network Meta-analysis a Revolutionary Statistical Tool for Improving the Reliability of Clinical Trial Results? A Brief Overview and Emerging Issues Arising. *In Vivo* **37,** 972-984.

Cipriani A, Barbui C, Salanti G, Rendell J, Brown R, Stockton S, Purgato M, Spineli LM, Goodwin GM & Geddes JR. (2011). Comparative efficacy and acceptability of antimanic drugs in acute mania: a multiple-treatments meta-analysis. *Lancet* **378,** 1306-1315.

Cipriani G, Ulivi M, Danti S, Lucetti C & Nuti A. (2016). Sexual disinhibition and dementia. *Psychogeriatrics* **16,** 145-153.

Harrer M, Cuijpers P, Furukawa TA & Ebert DD. (2021). *Doing meta-analysis with R: A hands-on guide*. Chapman and Hall/CRC.

Rücker G & Schwarzer G. (2015). Ranking treatments in frequentist network meta-analysis works without resampling methods. *BMC Med Res Methodol* **15,** 58.

Salanti G. (2012). Indirect and mixed-treatment comparison, network, or multiple-treatments meta-analysis: many names, many benefits, many concerns for the next generation evidence synthesis tool. *Research synthesis methods* **3,** 80-97.

Shannon CE. (1948). A mathematical theory of communication. *The Bell system technical journal* **27,** 379-423.

Veroniki AA, Straus SE, Rücker G & Tricco AC. (2018). Is providing uncertainty intervals in treatment ranking helpful in a network meta-analysis? *J Clin Epidemiol* **100,** 122-129.

Wu Y-C, Shih M-C & Tu Y-K. (2021). Using normalized entropy to measure uncertainty of rankings for network meta-analyses. *Medical Decision Making* **41,** 706-713.
