## Supplementary material for "Donanemab outperformed Aducanumab and Lecanemab on cognitive, but not on biomarker and safety outcomes: systematic review, frequentist and Bayesian network meta-analyses": 08.Supplementary Figures.pdf

**Figure S1. Power Analysis**

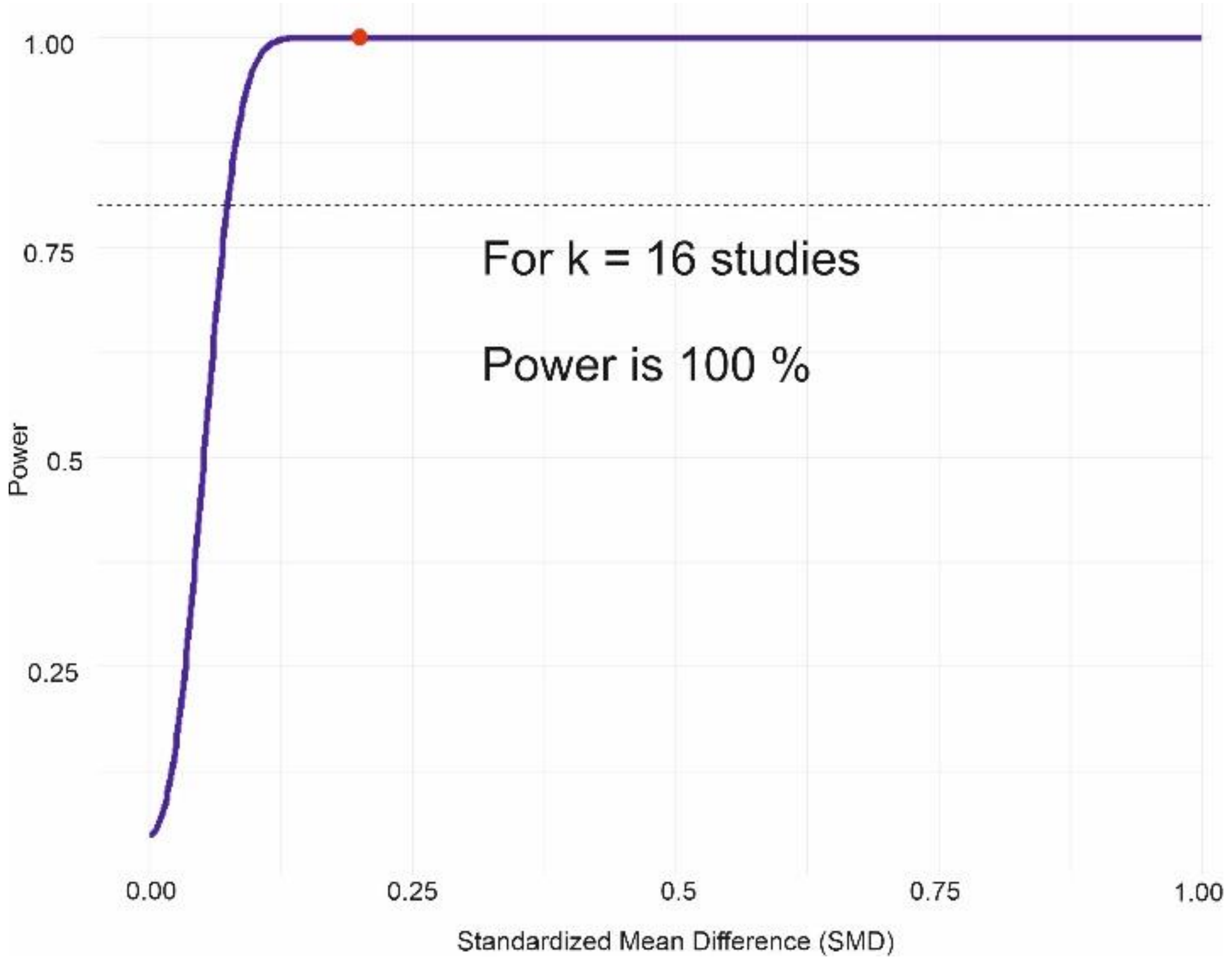

**Figure S2a. Risk of Bias: Summary Plot**

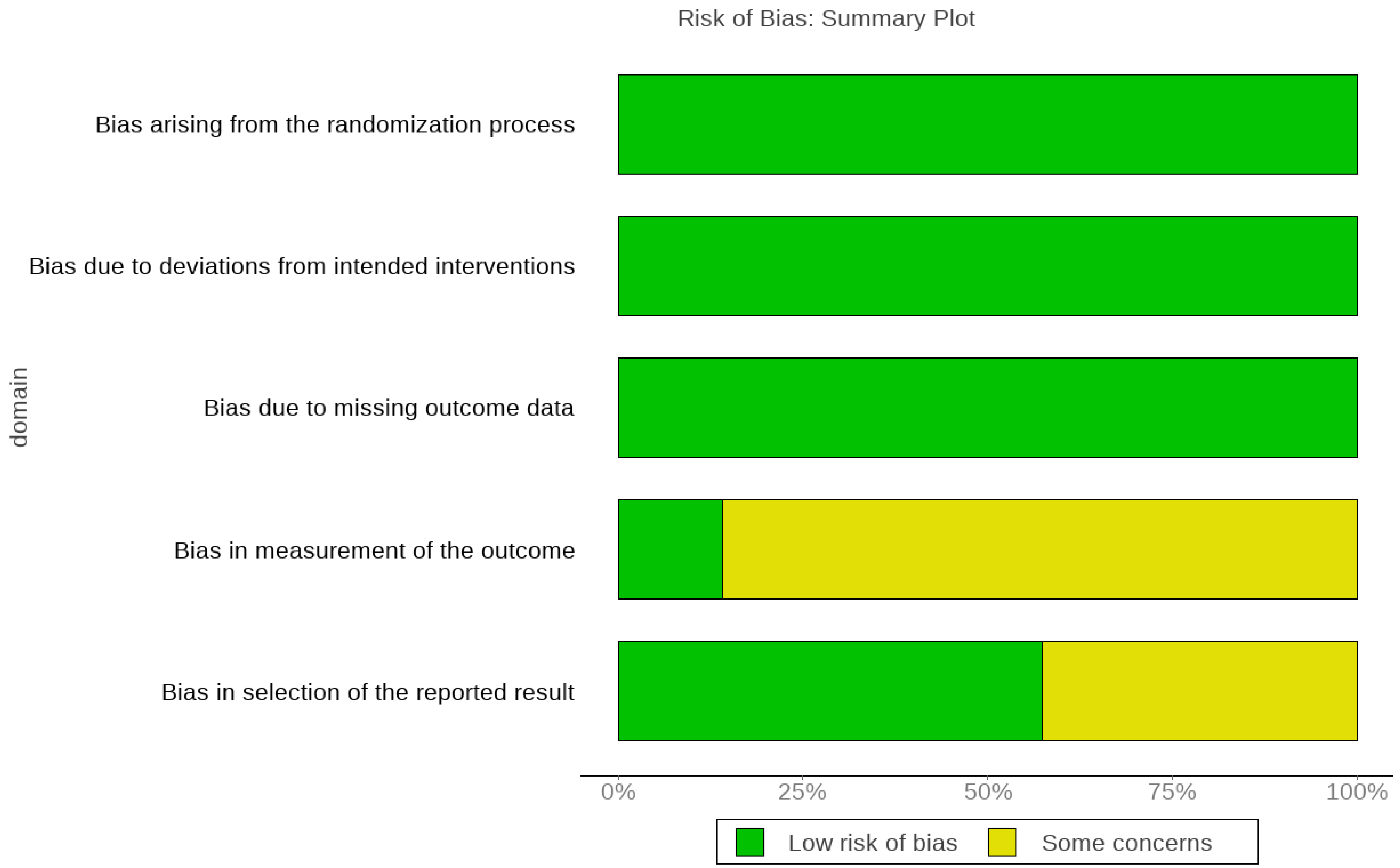

**Figure S2b. Risk of Bias: Traffic Light Plot**

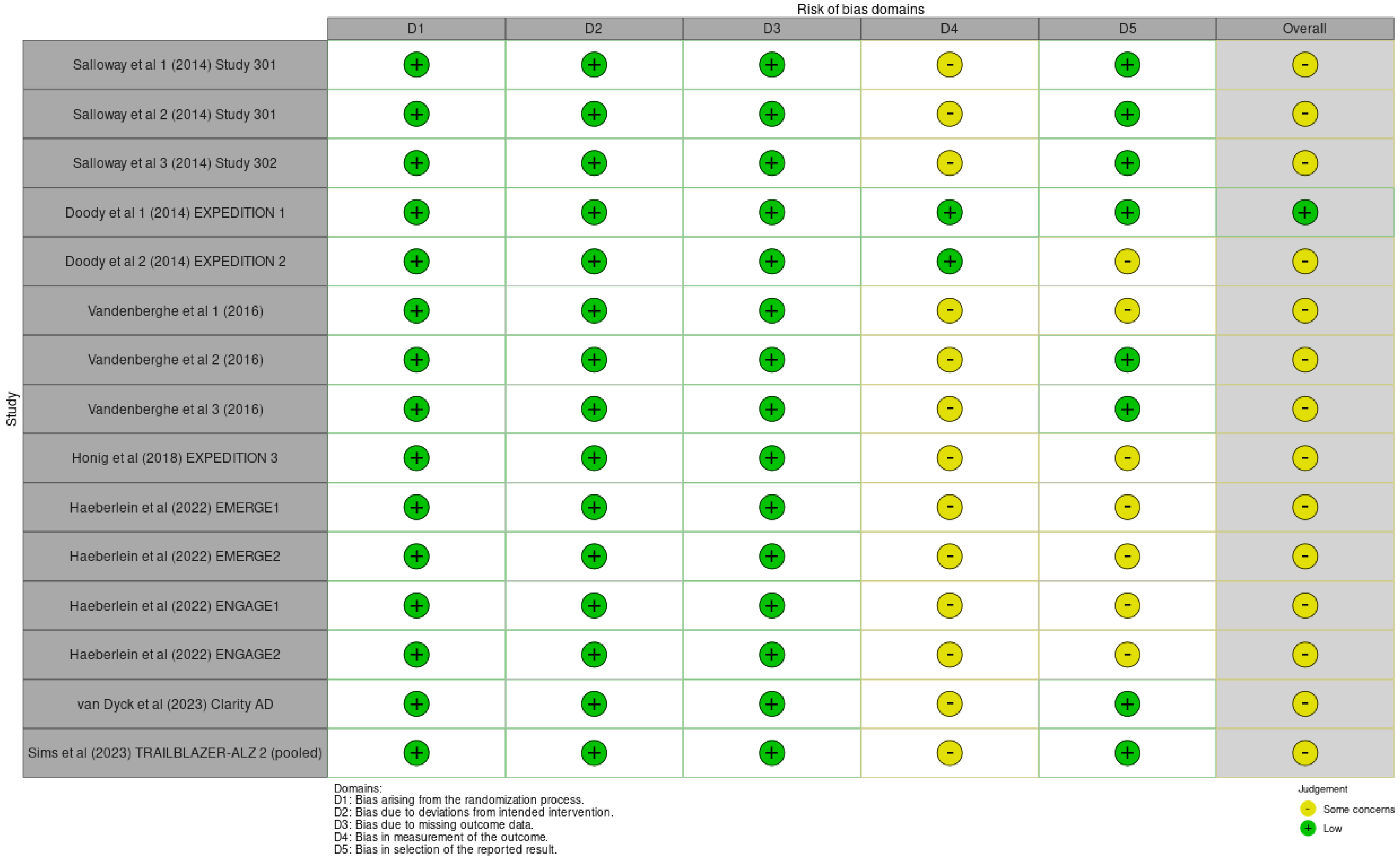

**Figure S3a. Funnel Plots for ADAS-Cog (Frequentist NMA, random-effect model)**

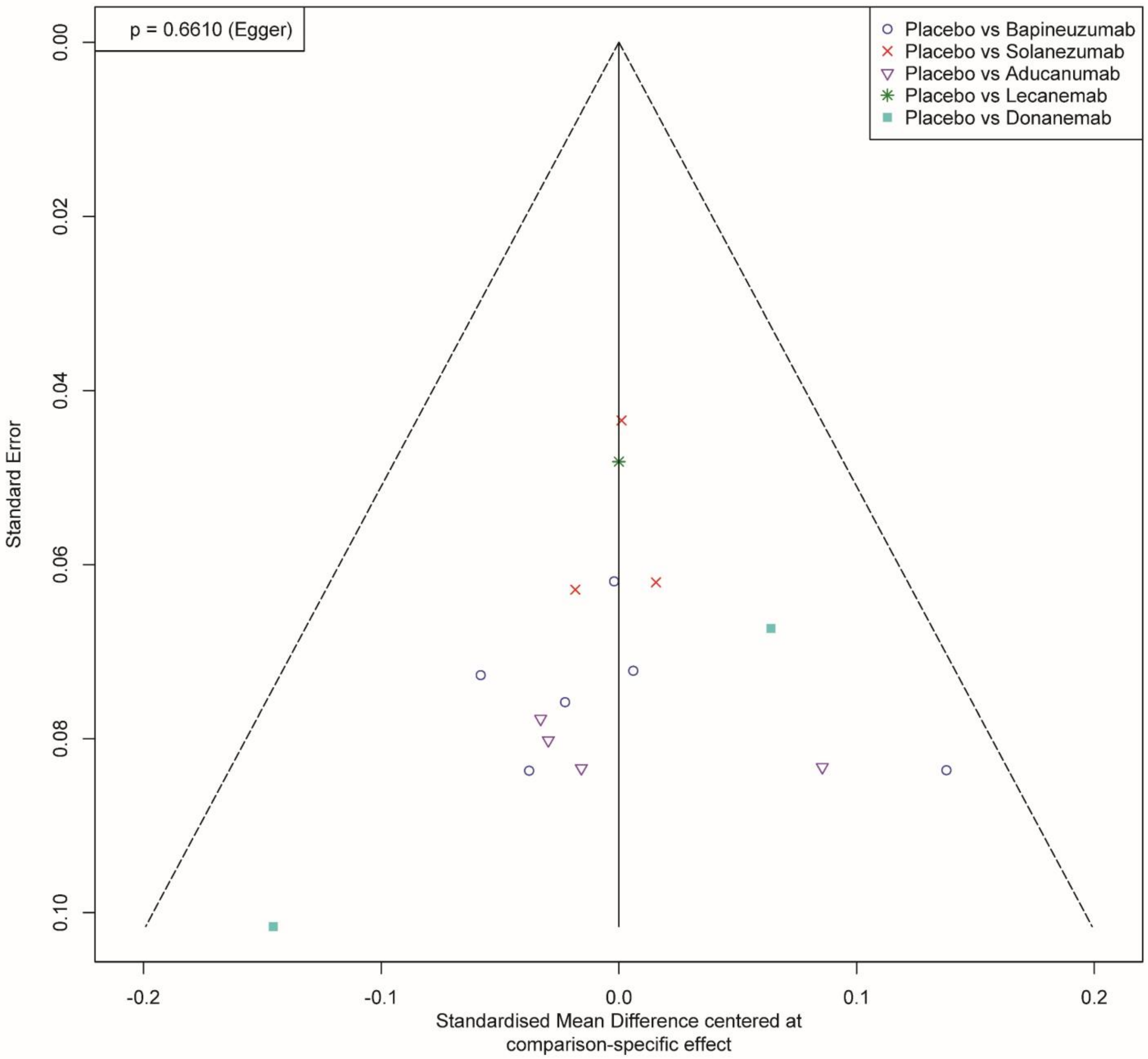

**Figure S3b. Funnel Plots for CDR-SB (Frequentist NMA, random-effect model)**

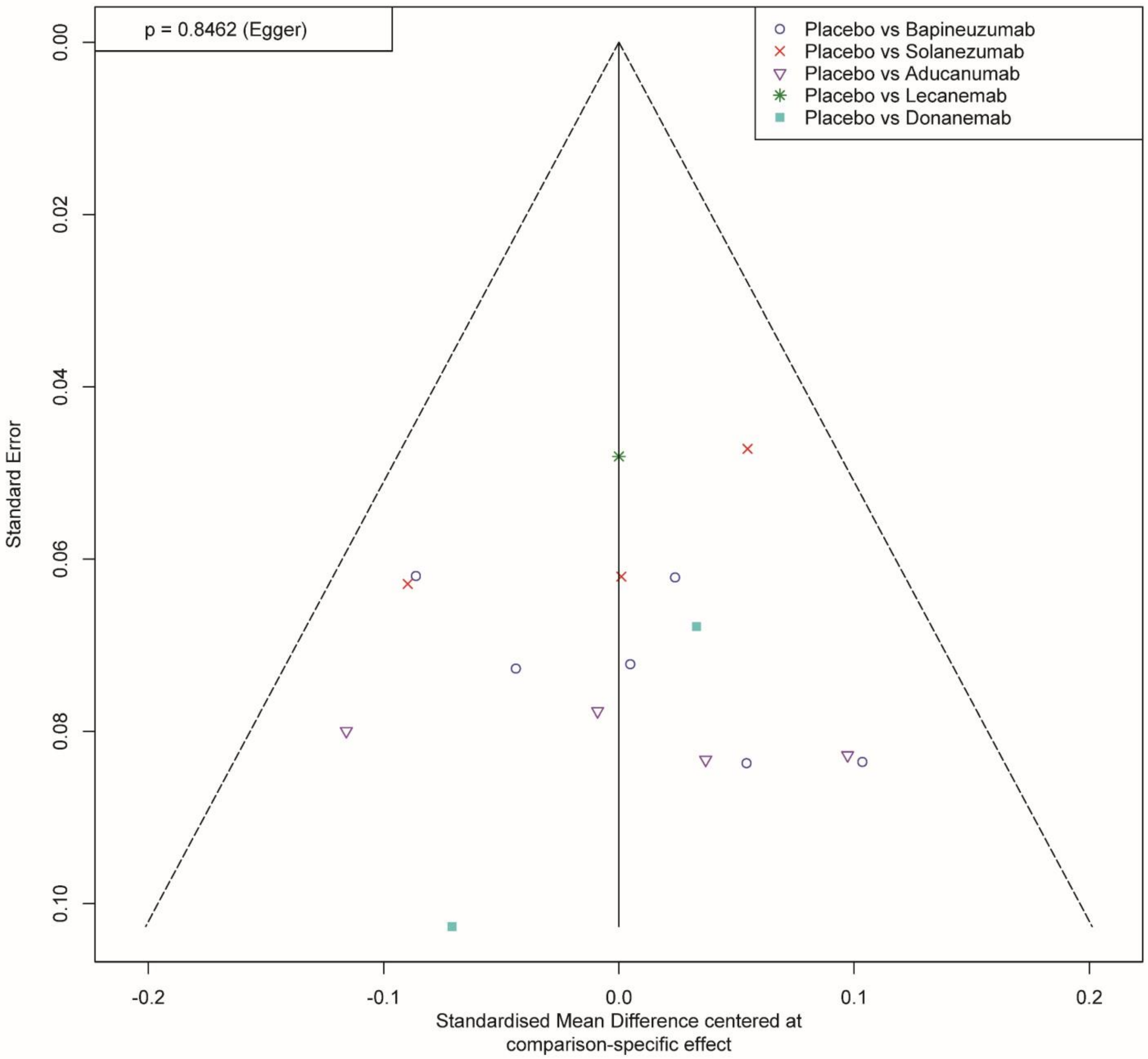

**Figure S3c. Funnel Plots for MMSE (Frequentist NMA, random-effect model)**

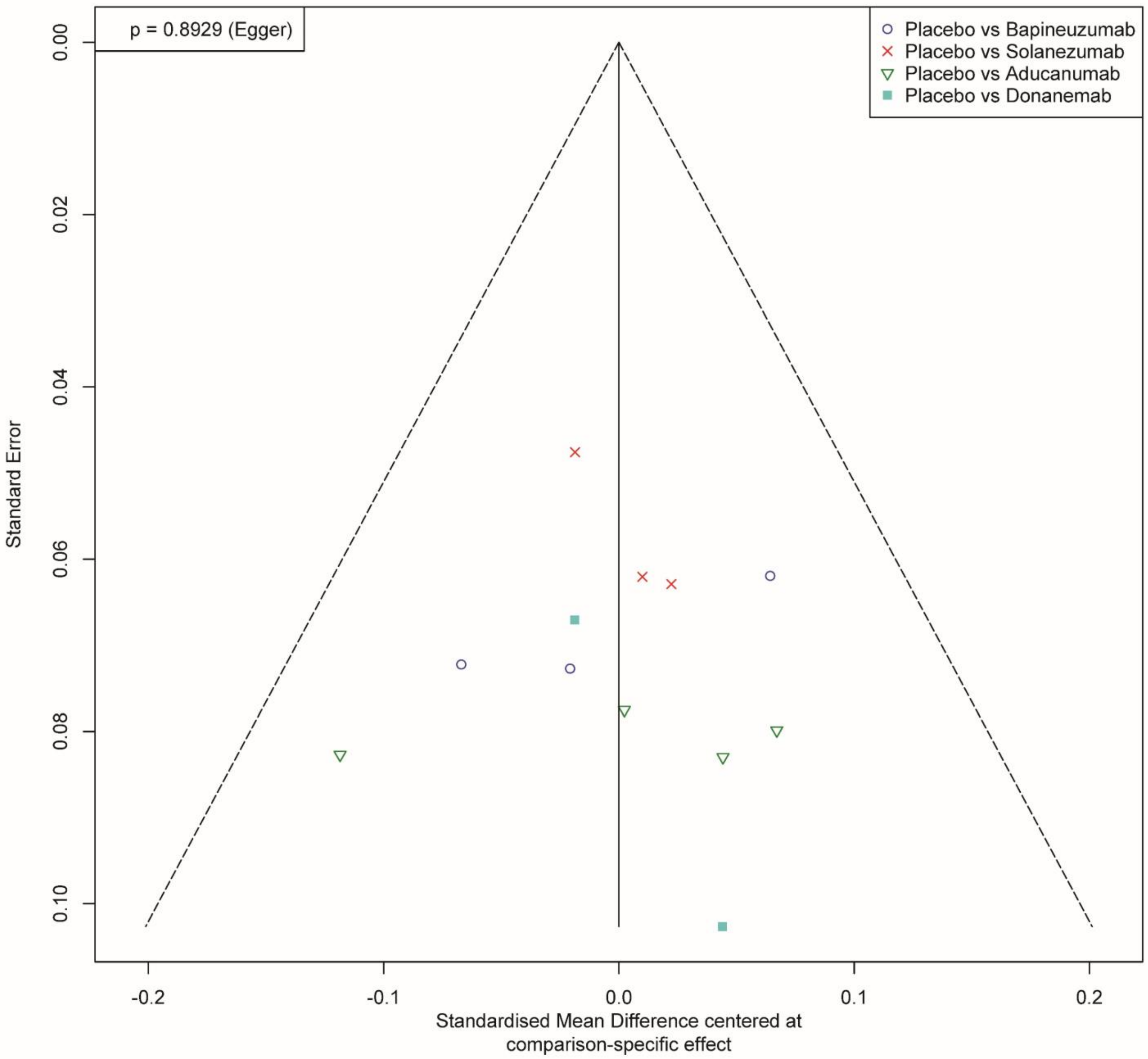

**Figure S4. GOSH Plot Diagnostics: ADAS-Cog (Gaussian Mixture Model, GMM)**

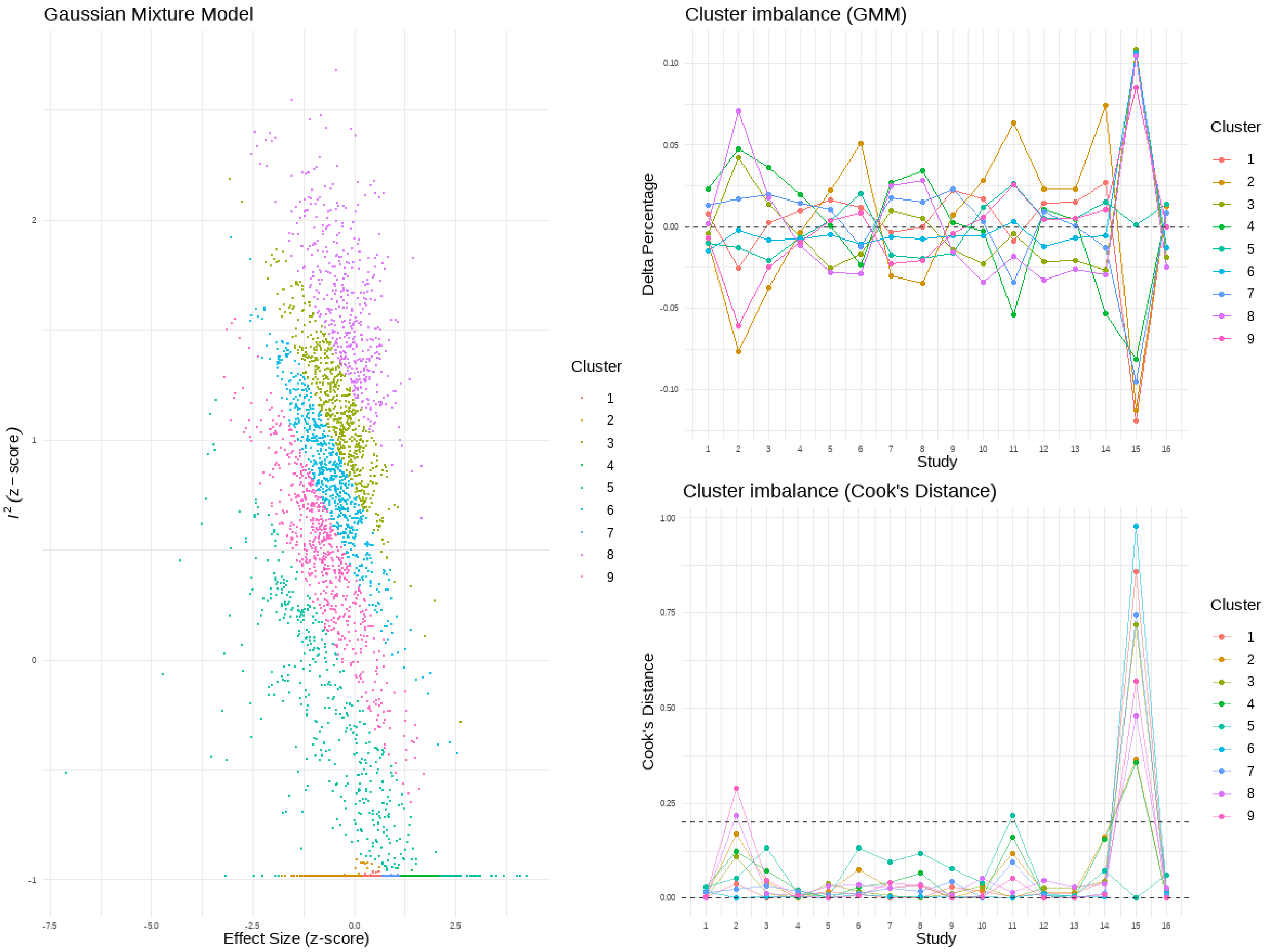

### Figure S5. GOSH Plot Diagnostics: CDR-SB (Gaussian Mixture Model, GMM)

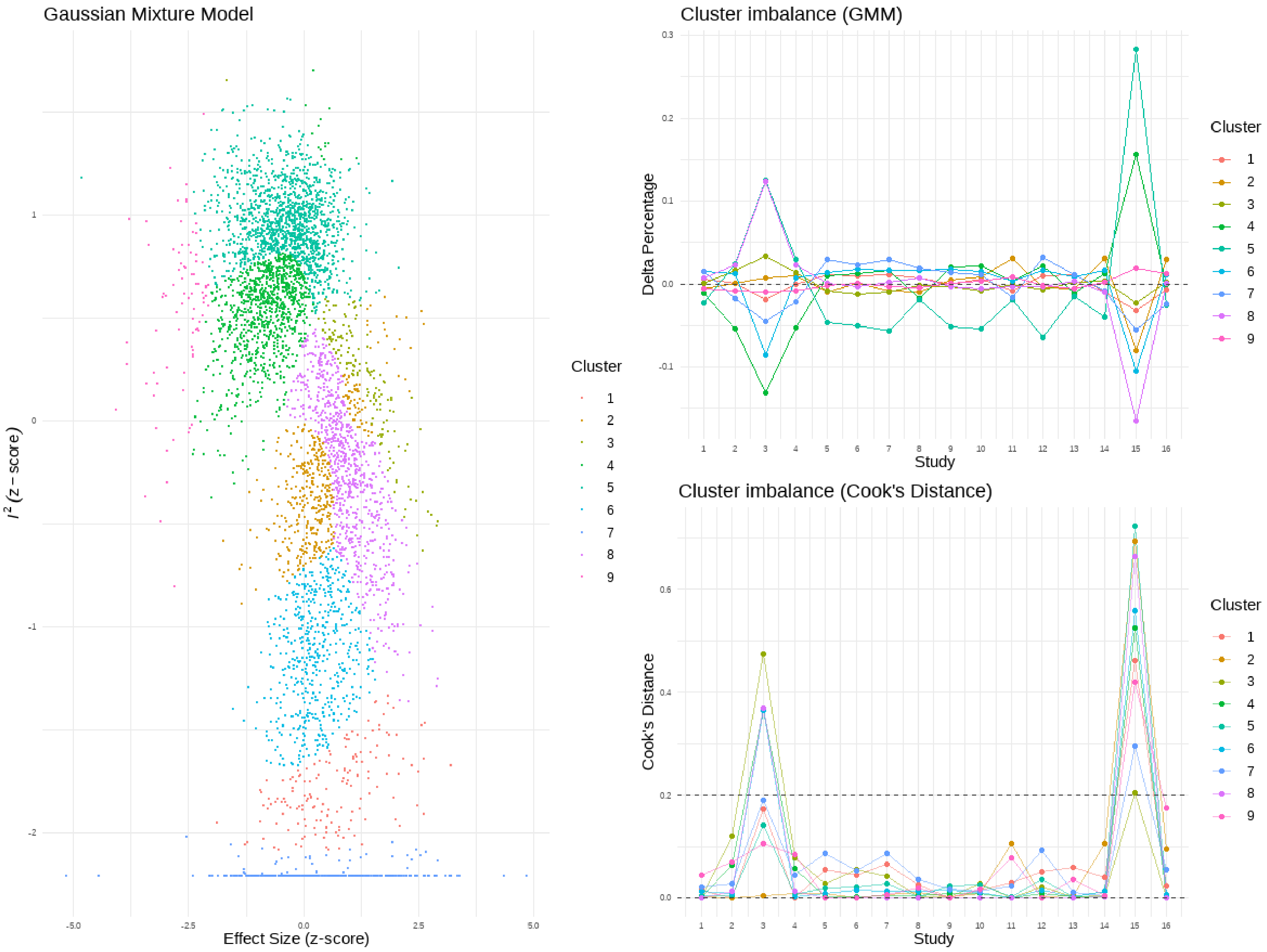

**Figure S6. GOSH Plot Diagnostics: MMSE (Gaussian Mixture Model, GMM)**

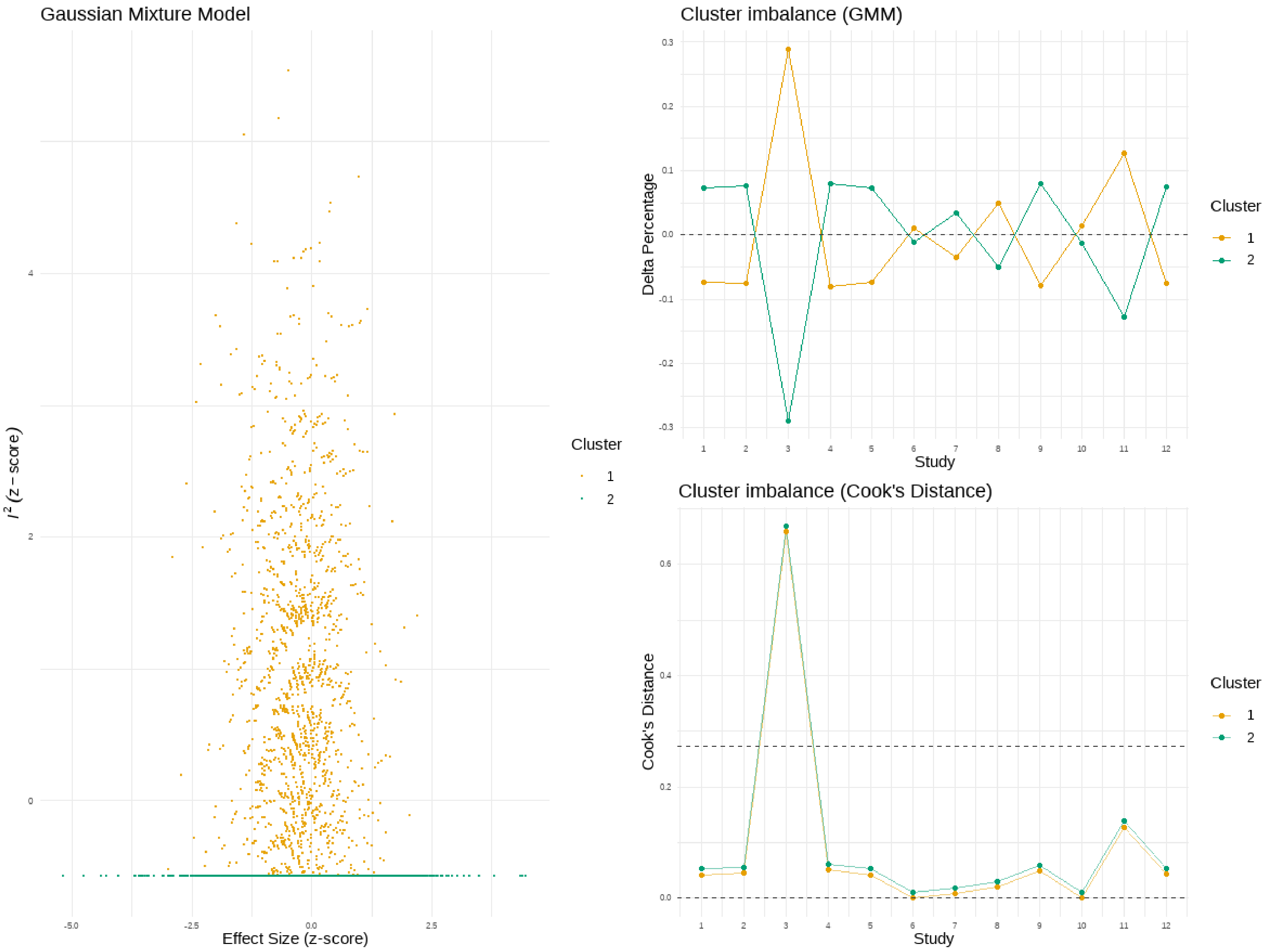

### Figure S7. GOSH Plot Diagnostics: Amyloid Burden on PET (Gaussian Mixture Model, GMM)

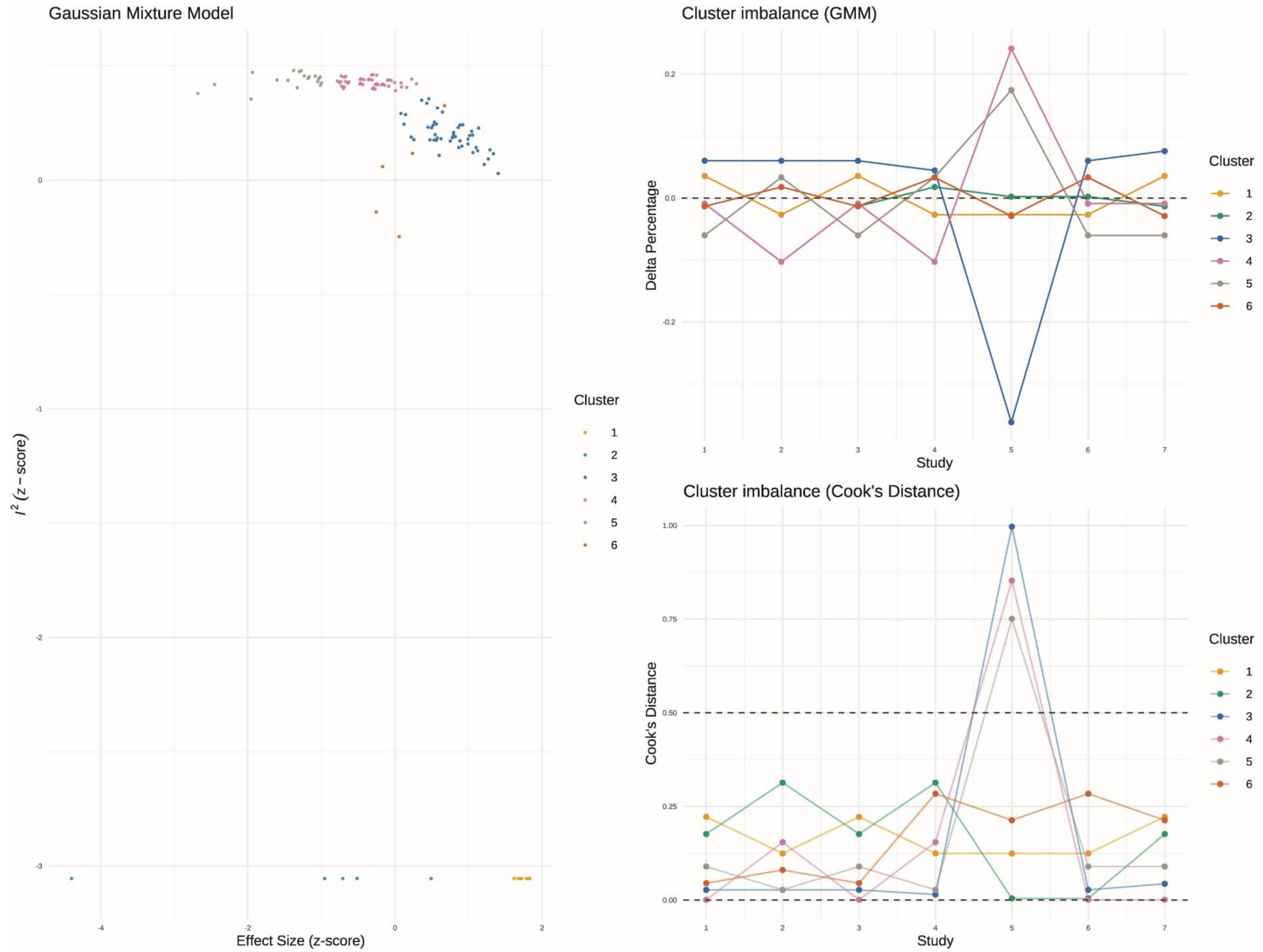

**Figure S8. GOSH Plot Diagnostics: Treatment Discontinuations due to Adverse Events (Gaussian Mixture Model, GMM)**

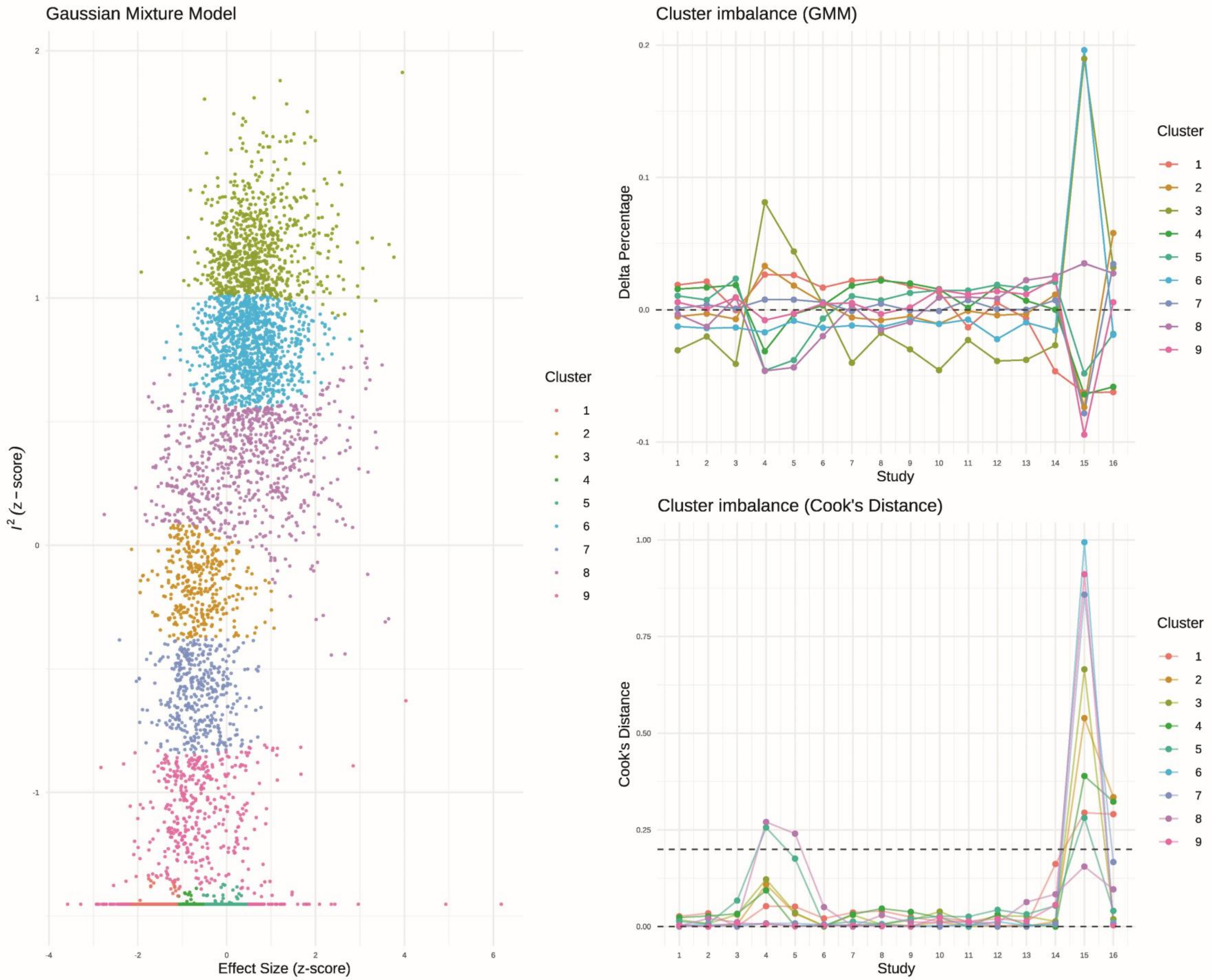

**Figure S9. GOSH Plot Diagnostics: Serious Side Effects (Gaussian Mixture Model, GMM)**

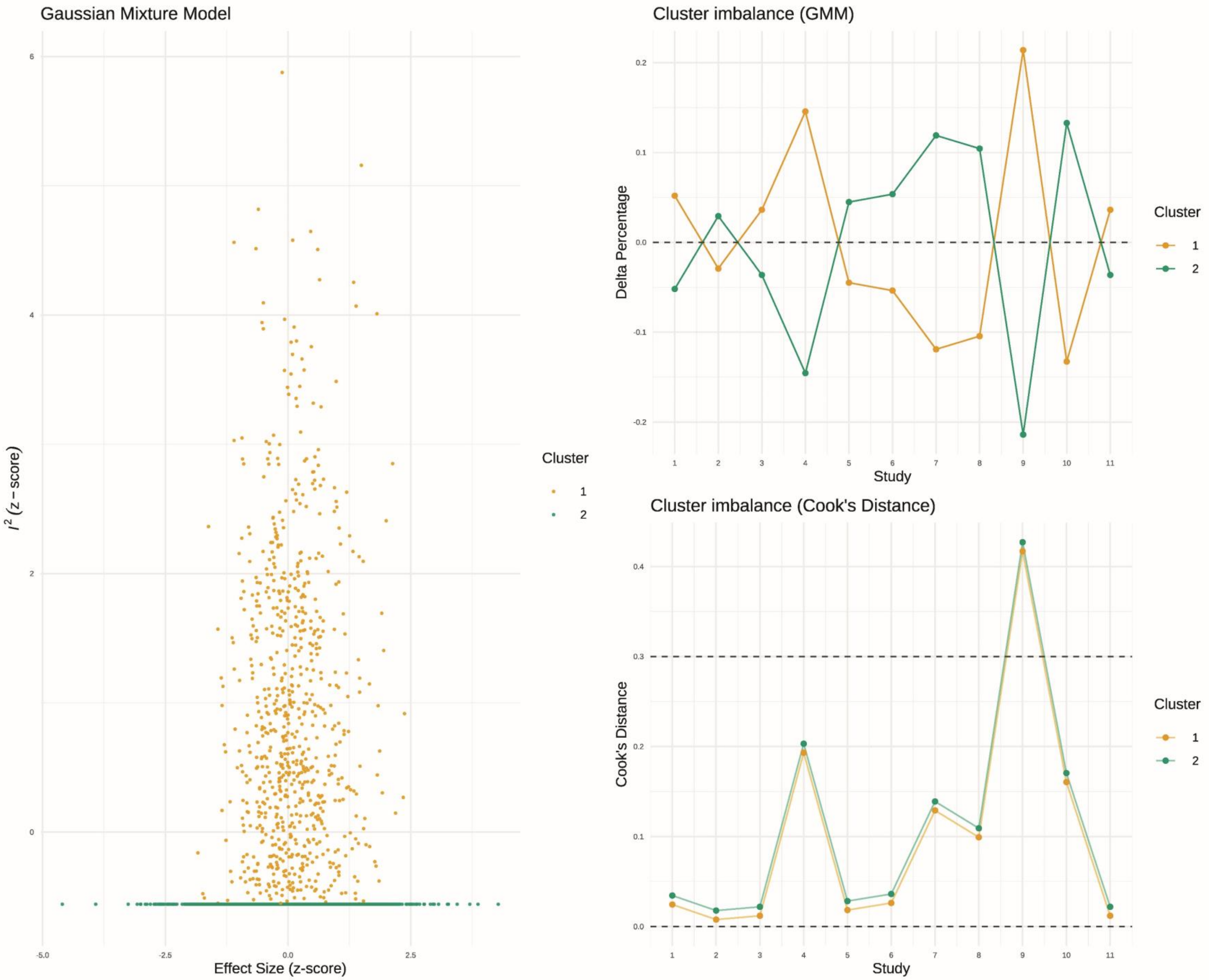

### Figure S10. GOSH Plot Diagnostics: Total events of ARIA-E (Gaussian Mixture Model, GMM)

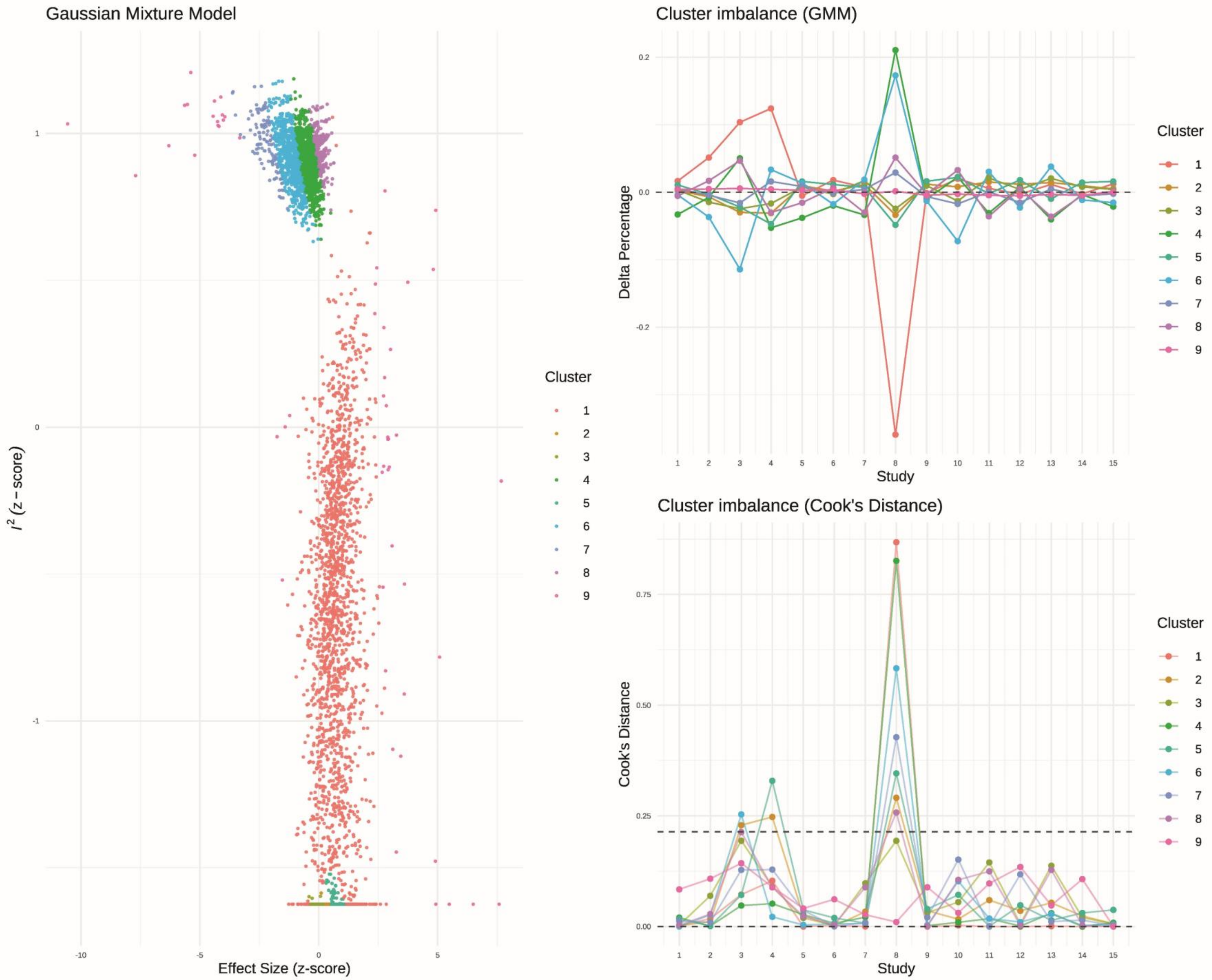

**Figure S11. GOSH Plot Diagnostics: Headaches (Gaussian Mixture Model, GMM)**

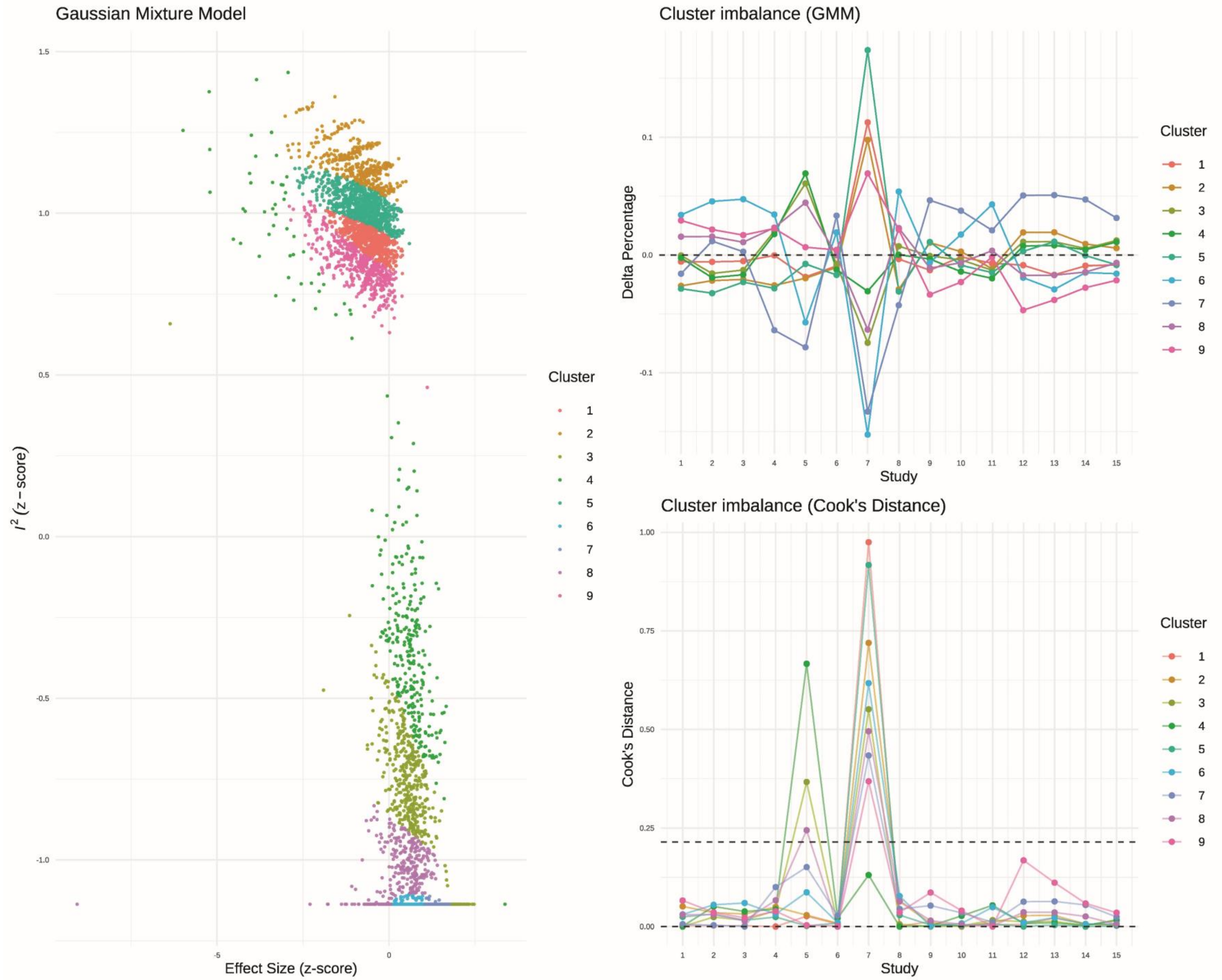

**Figure S12. GOSH Plot Diagnostics: Syncope (Gaussian Mixture Model, GMM)**

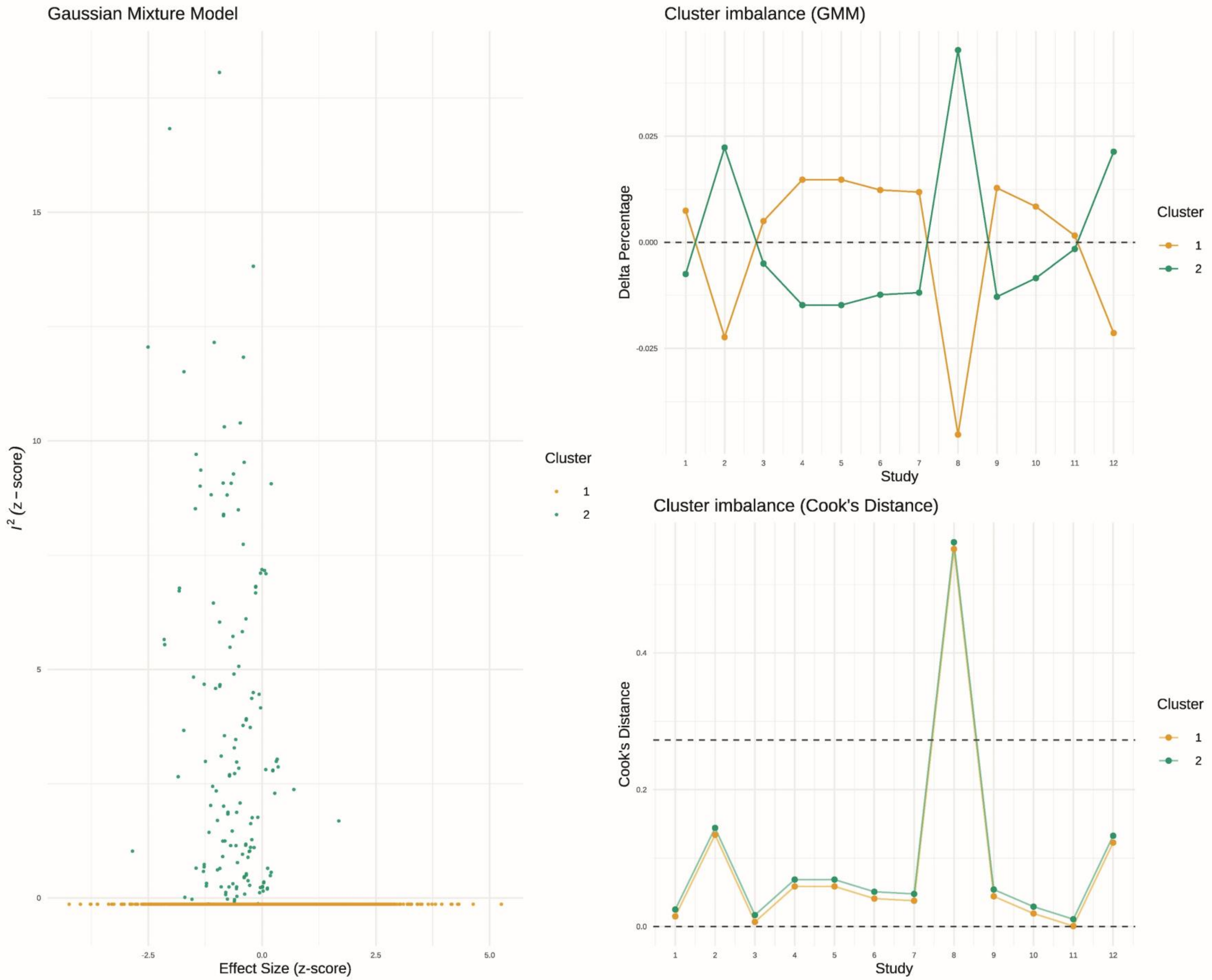

**Figure S13. GOSH Plot Diagnostics: Dizziness (Gaussian Mixture Model, GMM)**

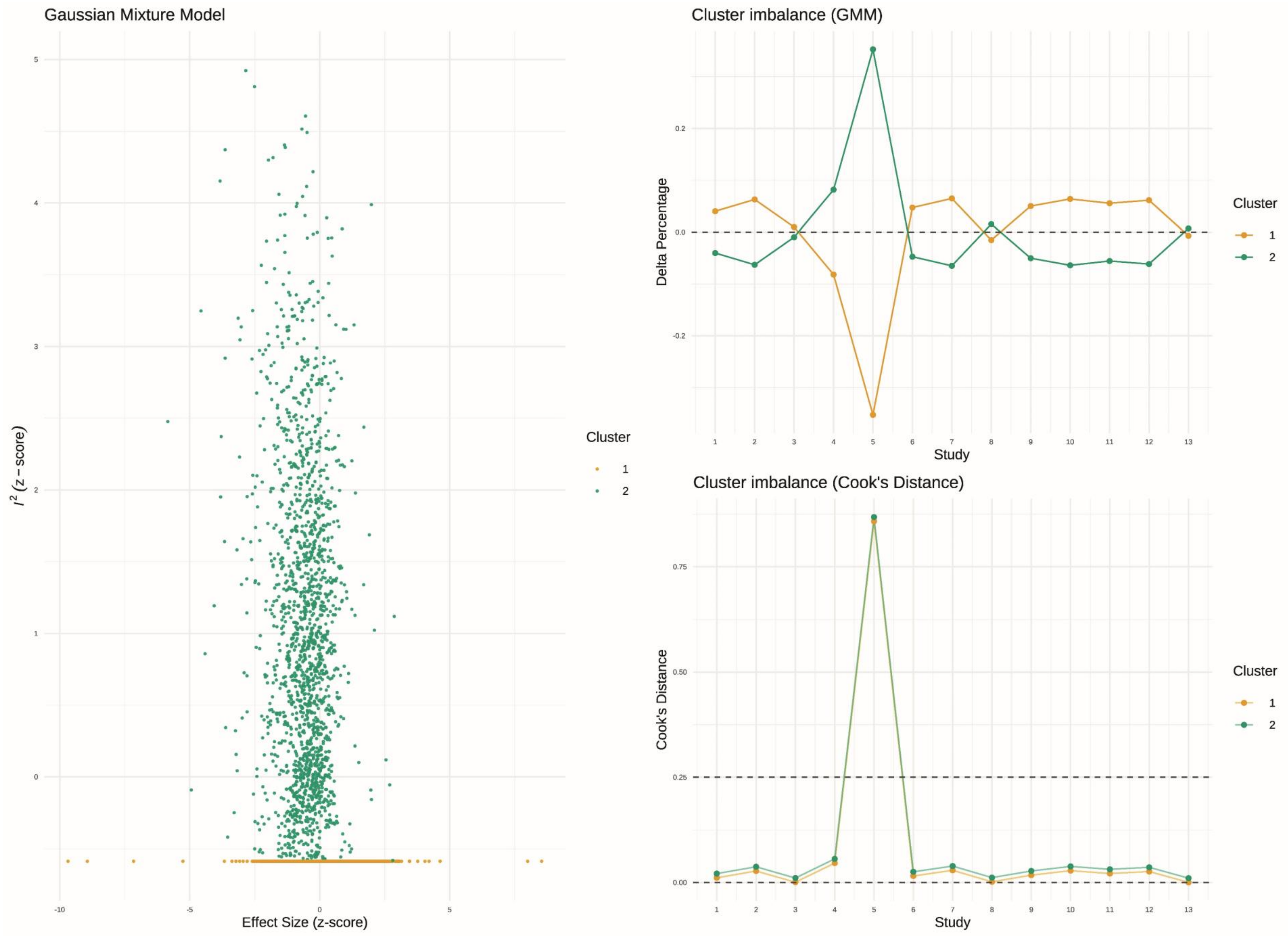

**Figure S14. GOSH Plot Diagnostics: Fatigue (Gaussian Mixture Model, GMM)**

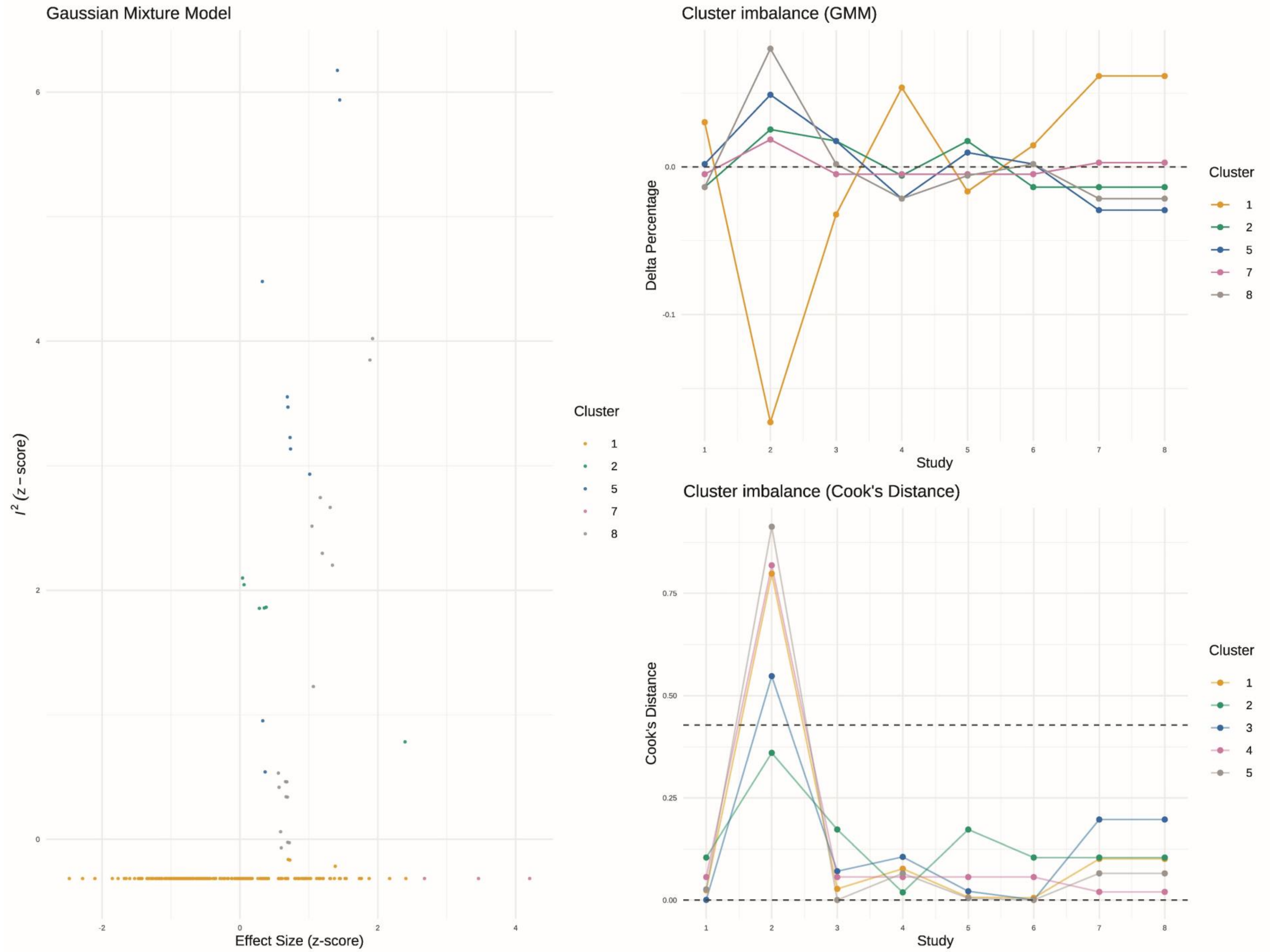

**Figure S15. GOSH Plot Diagnostics: Nausea (Gaussian Mixture Model, GMM)**

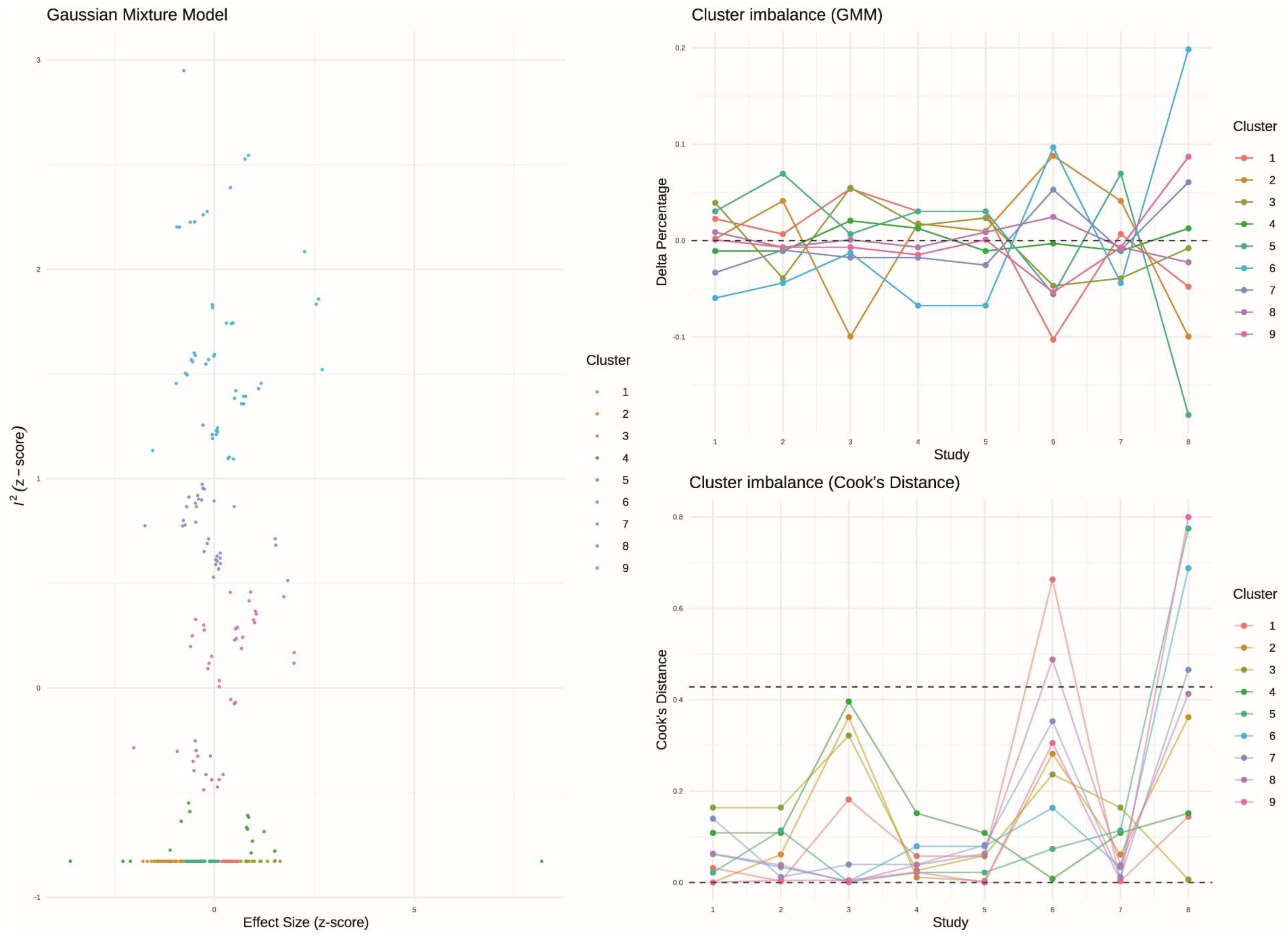

**Figure S16. GOSH Plot Diagnostics: Total Events of ARIA-E in APOE-e4 carriers (Gaussian Mixture Model, GMM)**

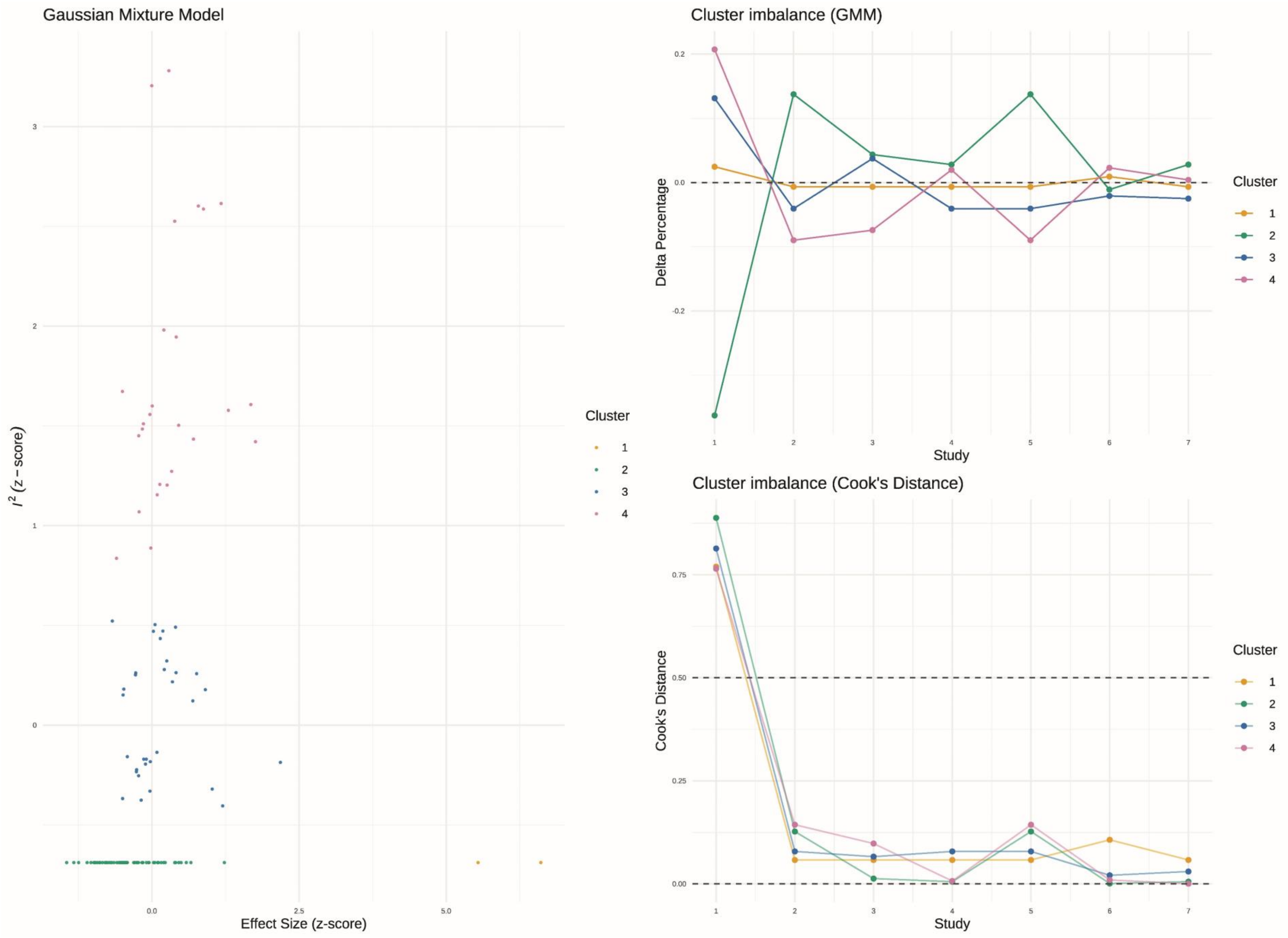

**Figure S17. GOSH Plot Diagnostics: Total Events of ARIA-E in APOE-e4 non-carriers (Gaussian Mixture Model, GMM)**

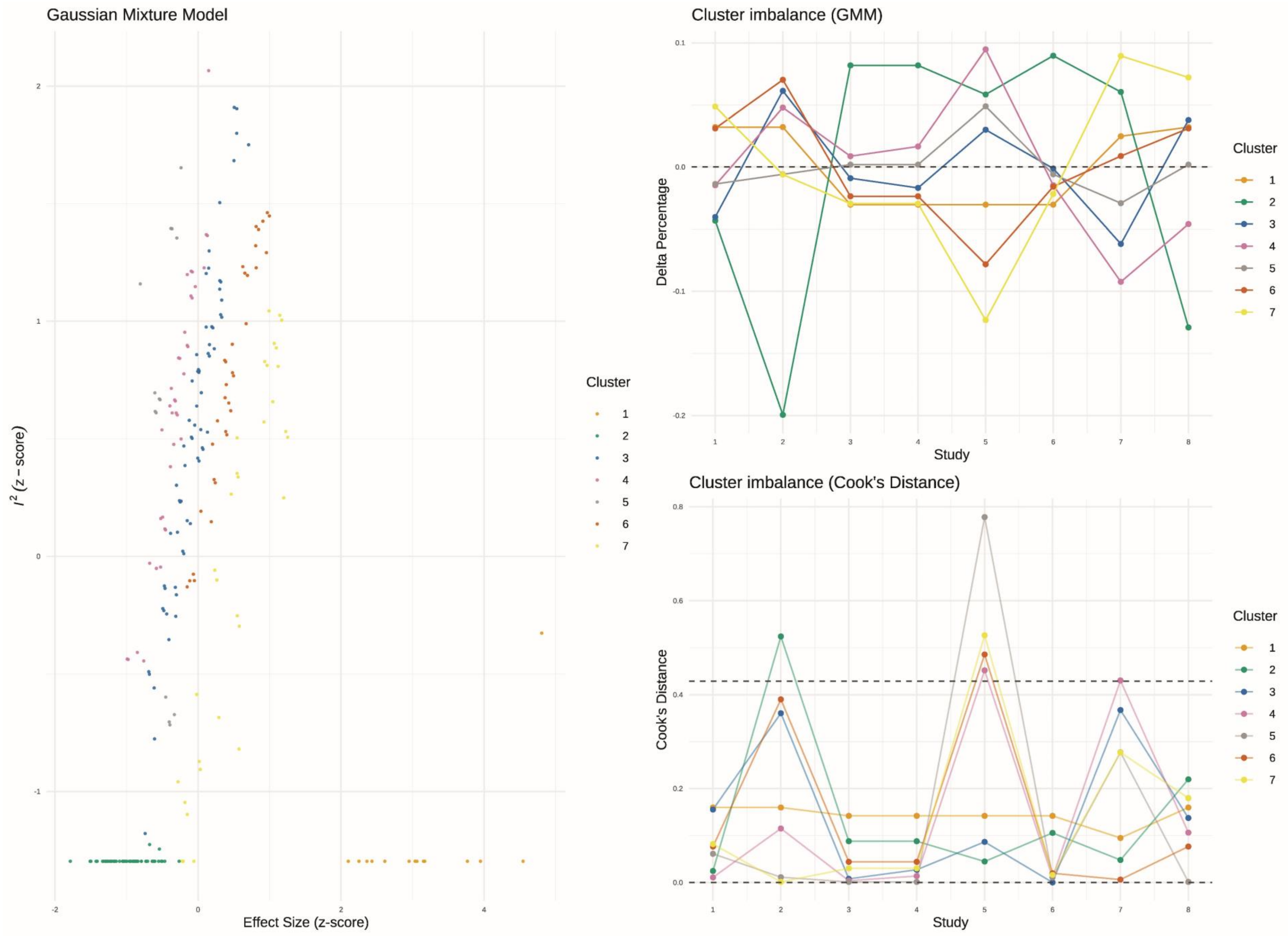

**Figure S18. GOSH Plot Diagnostics: Total Events of ARIA-H (Gaussian Mixture Model, GMM)**

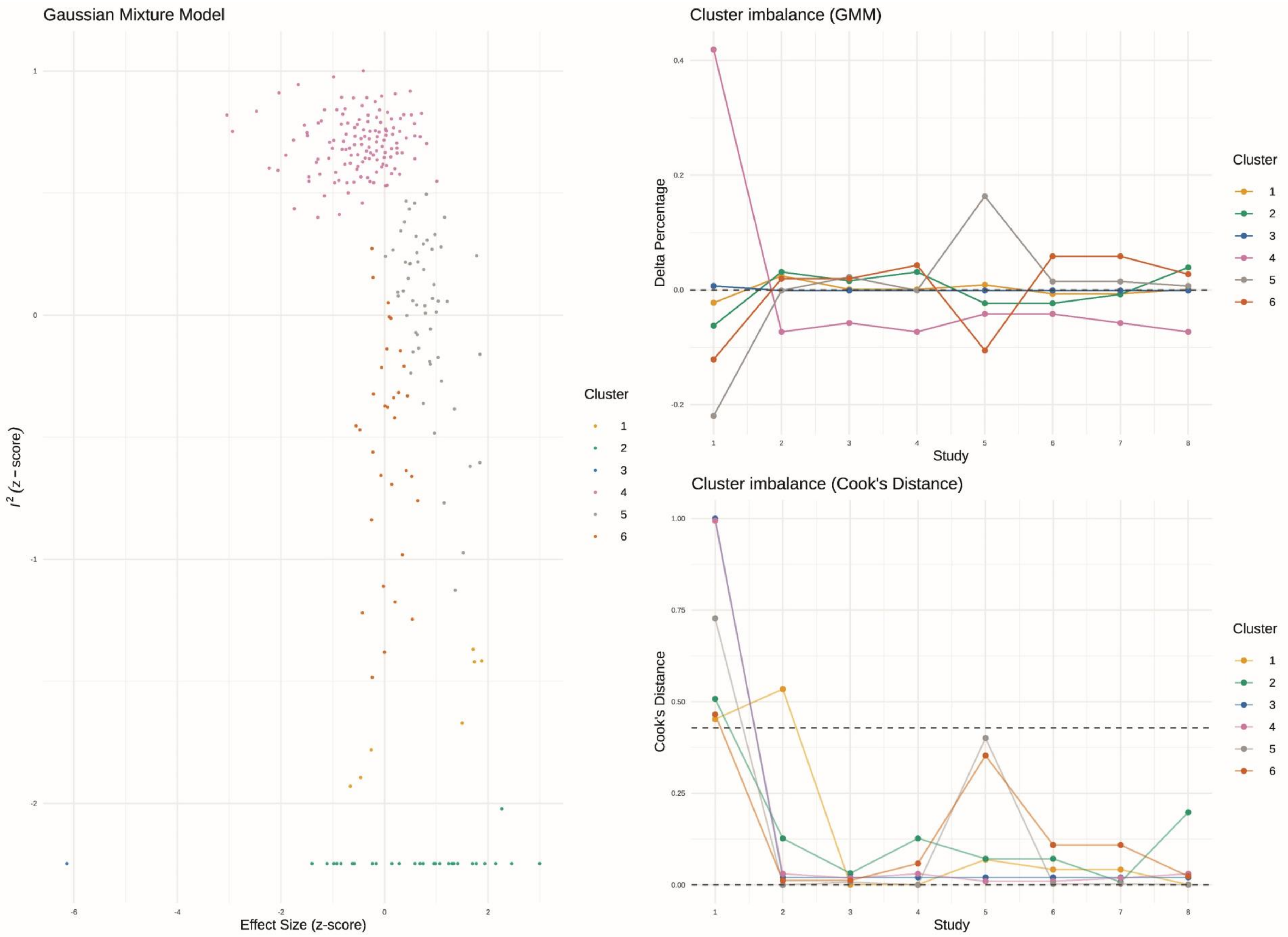

**Figure S19. GOSH Plot Diagnostics: ARIA-H (Microhemorrhages reported) (Gaussian Mixture Model, GMM)**

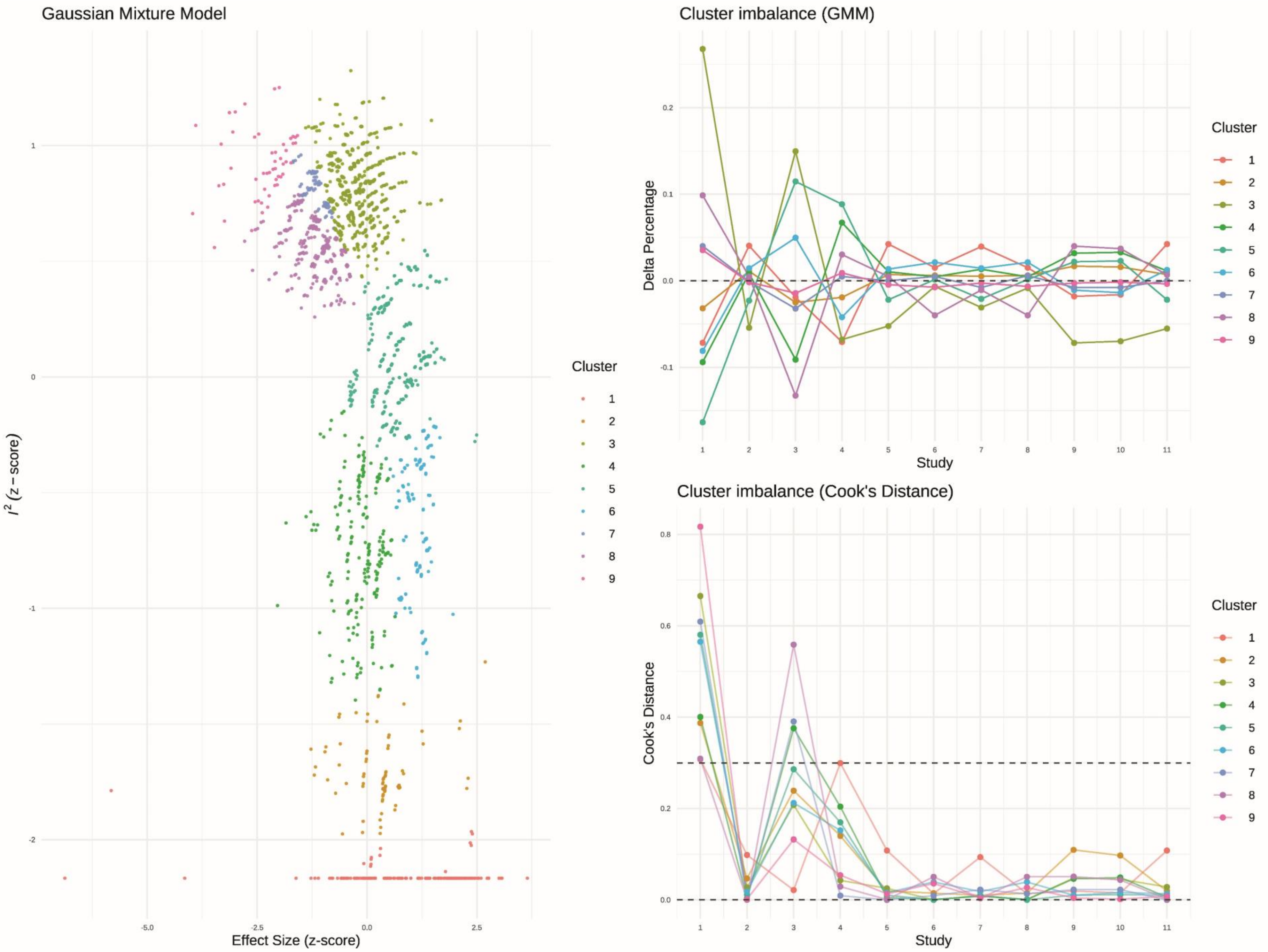

**Figure S20. GOSH Plot Diagnostics: Arthralgia (Gaussian Mixture Model, GMM)**

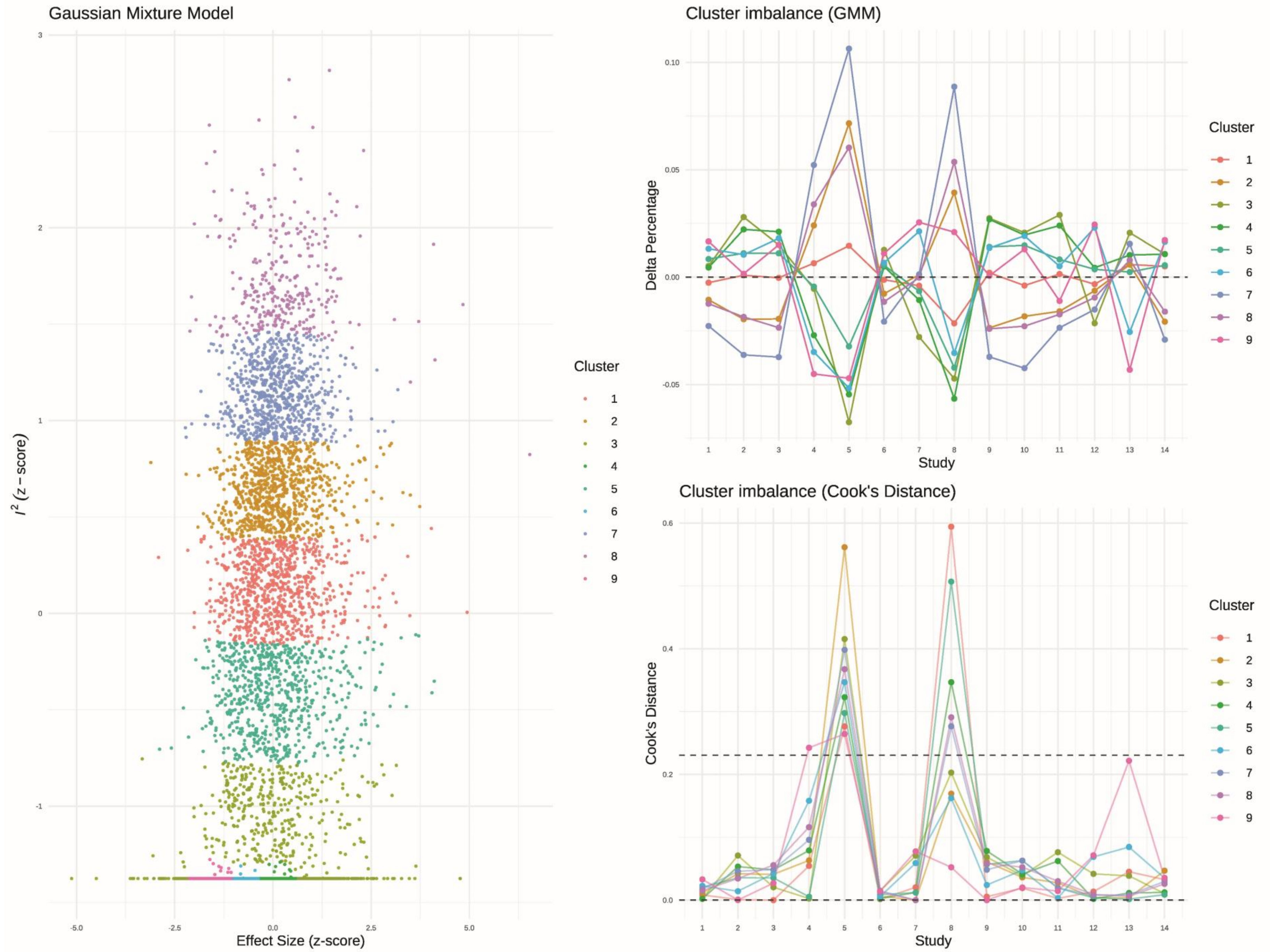

**Figure S21. GOSH Plot Diagnostics: Back Pain (Gaussian Mixture Model, GMM)**

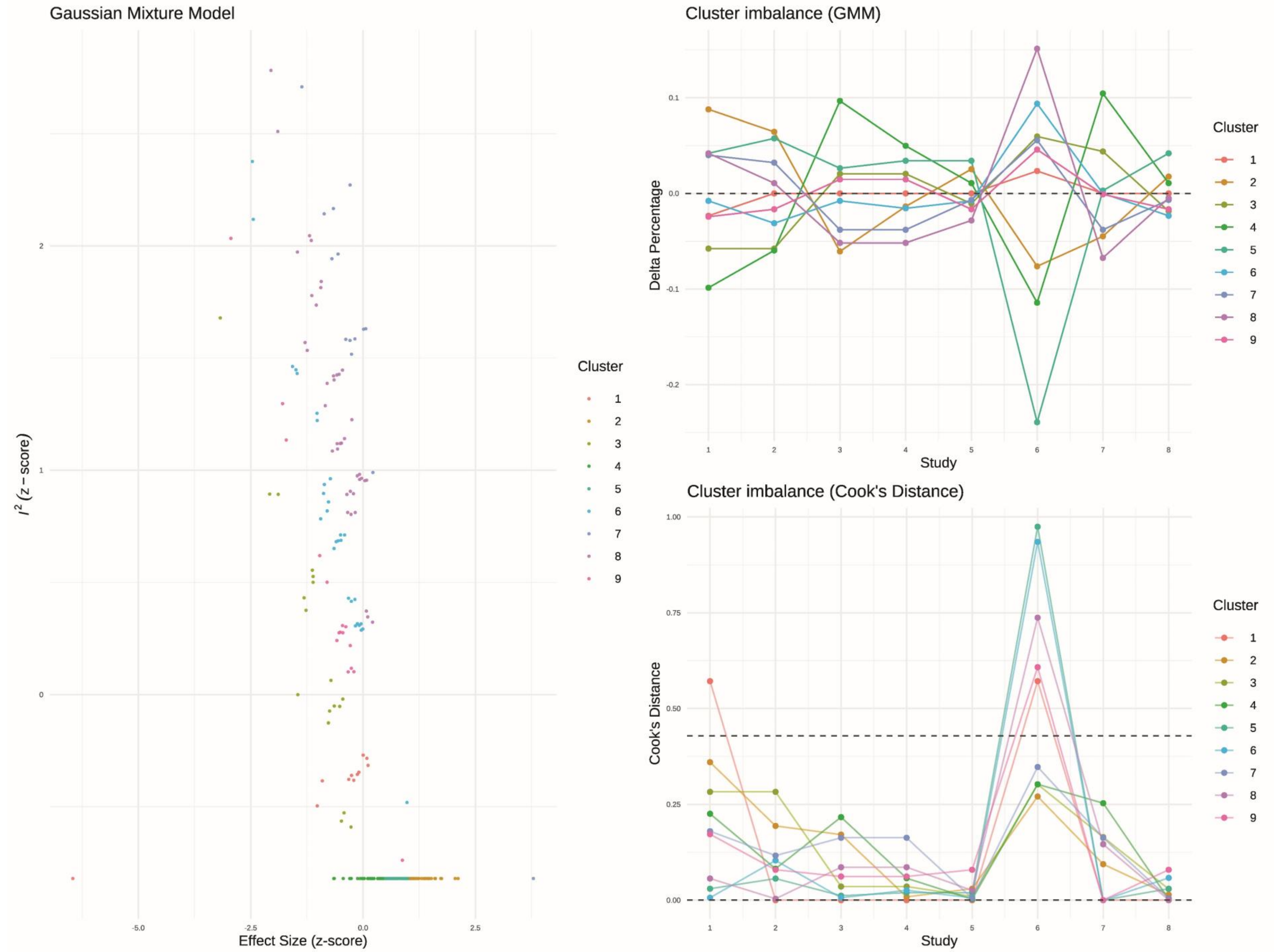

**Figure S22. GOSH Plot Diagnostics: Diarrhea (Gaussian Mixture Model, GMM)**

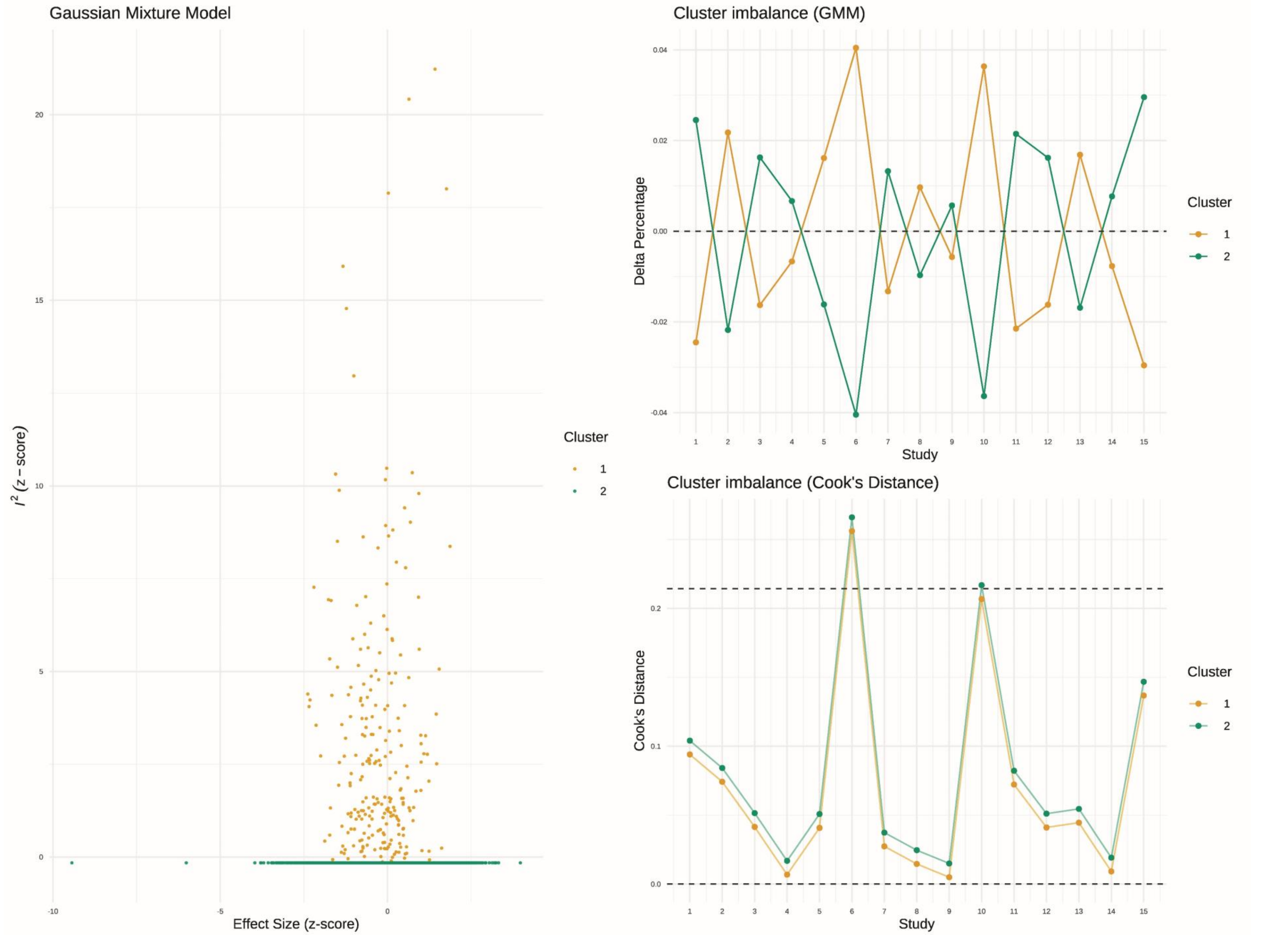

**Figure S23. GOSH Plot Diagnostics: Fall (Gaussian Mixture Model, GMM)**

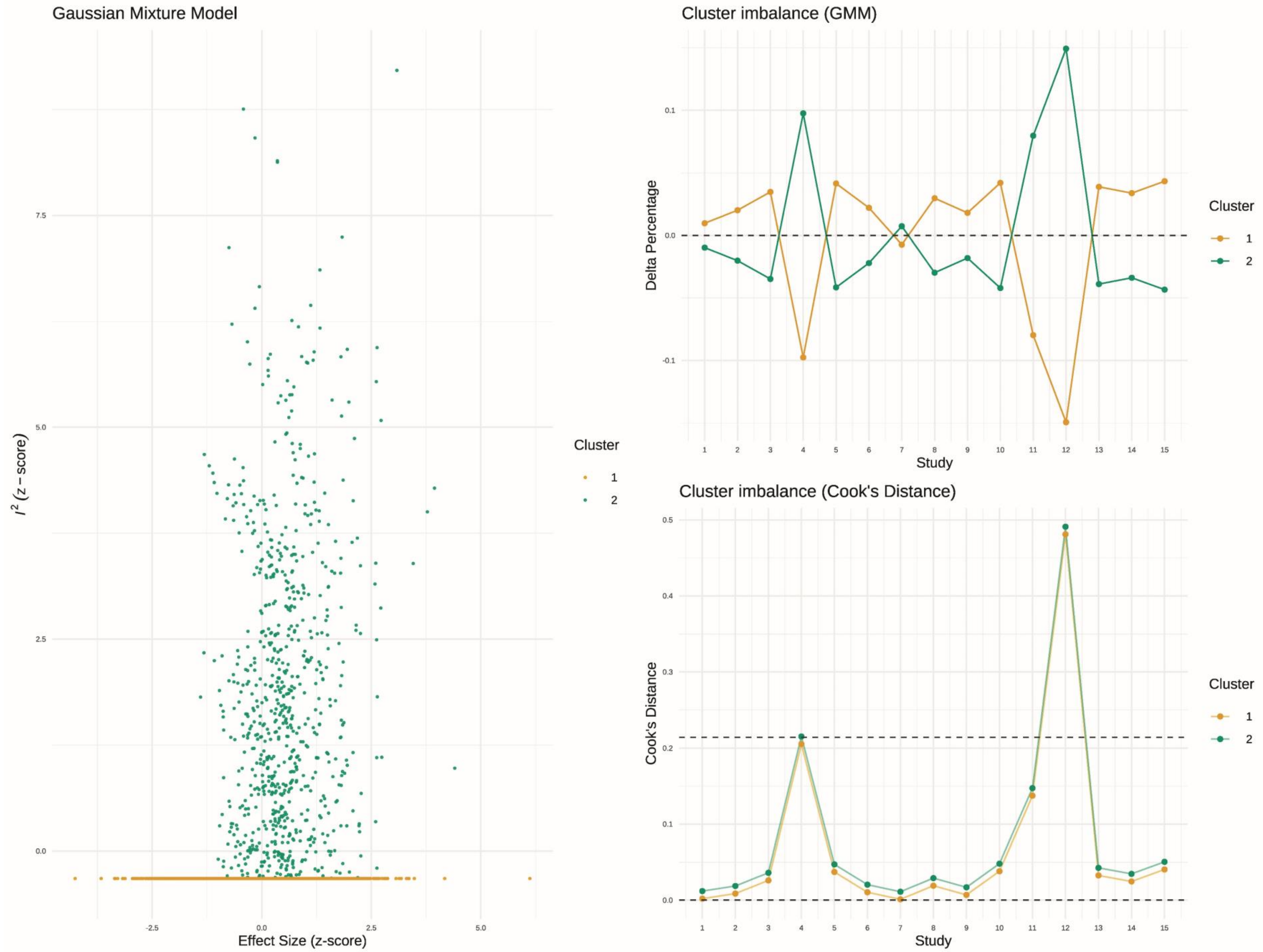

**Figure S24. GOSH Plot Diagnostics: Urinary Infections (Gaussian Mixture Model, GMM)**

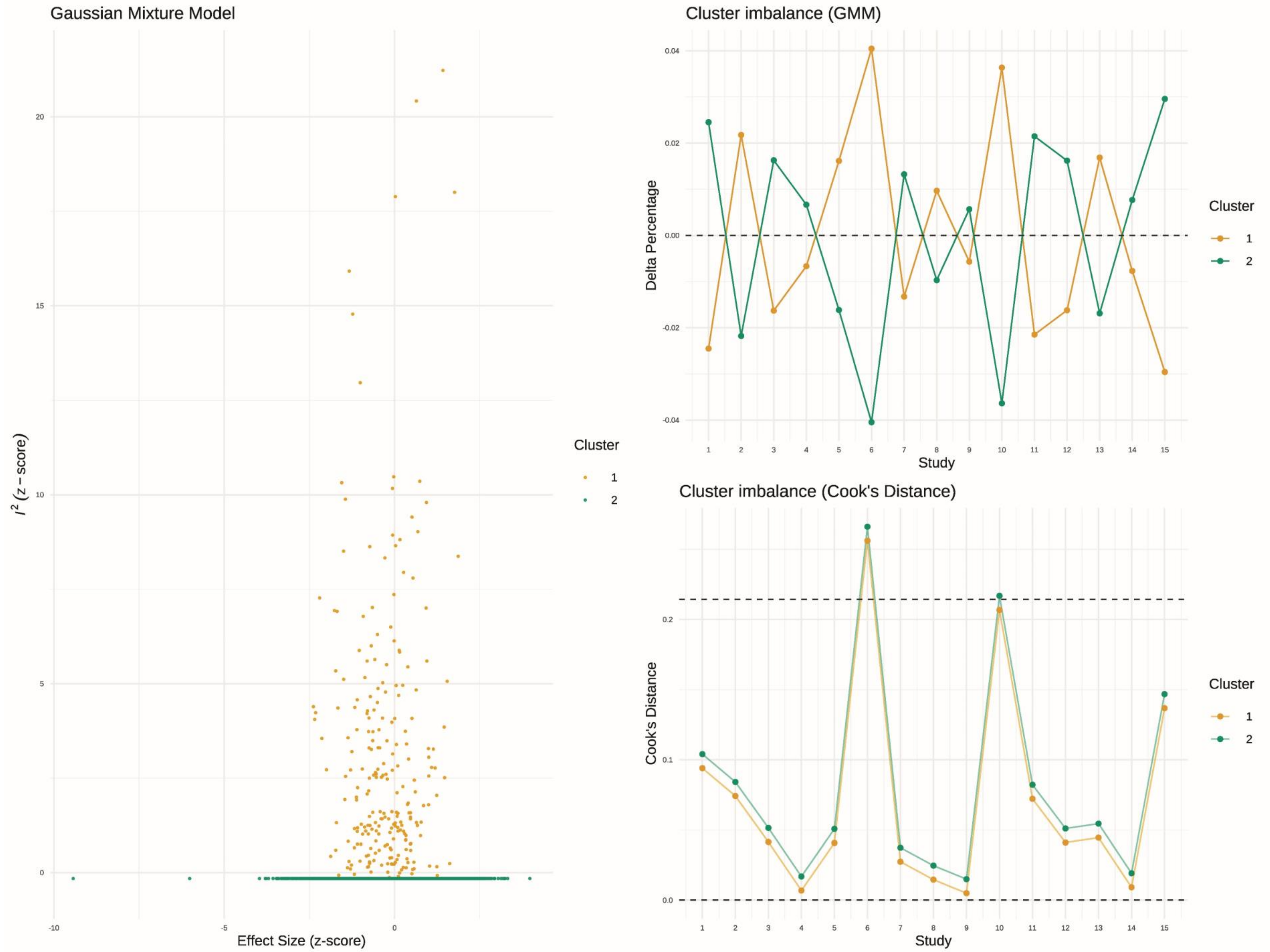
